# Genealogy based trait association with LOCATER boosts power at loci with allelic heterogeneity

**DOI:** 10.1101/2024.11.04.24316696

**Authors:** Xinxin Wang, Ryan Christ, Erica Young, Chul Joo Kang, Indraniel Das, Edward A. Belter, Markku Laakso, Louis J.M. Aslett, David Steinsaltz, Nathan O. Stitziel, Ira M. Hall

**Author notes:** Institute for Informatics, Data Science & Biostatistics, Washington University in St. Louis, Saint Louis, MO, 63110, USA. Collaborative and Integrative Genomics Lab at the McDonnell Genome Institute at Washington University School of Medicine, Saint Louis, MO, 63108, USA.

## Abstract

A key methodological challenge for genome-wide association studies is how to leverage haplotype diversity and allelic heterogeneity to improve trait association power, especially in noncoding regions where it is difficult to predict variant impacts and define functional units for variant aggregation. Genealogy-based association methods have the potential to bridge this gap by testing combinations of common and rare haplotypes based purely on their ancestral relationships. In parallel work, we have developed an efficient local ancestry inference engine and a novel statistical method (LOCATER) for combining signals present on different branches of a locus specific haplotype tree. Here, we developed a genome-wide LOCATER analysis pipeline and applied it to a genome sequencing study of 6,795 Finnish individuals with 101 cardiometabolic traits and 18.9 million autosomal variants. We identify 351 significant trait associations at 47 distinct genomic loci and find that LOCATER boosts single marker test (SMT) association signal at 5 loci by combining independent signals from distinct alleles. LOCATER successfully recovers known quantitative trait loci not found by SMT, including *LIPG*, recovers known allelic heterogeneity at the *APOE/C1/C4/C2* gene cluster, and suggests one novel association. We find that confounders have a more pronounced effect on genealogy-based methods than SMT, and we propose a new randomization approach and a general method for genomic control to eliminate their effects. This study demonstrates that genealogy-based methods such as LOCATER excel when multiple causal variants are present and suggests that their application to larger and more diverse cohorts will be fruitful.

## INTRODUCTION

Genome-wide association studies have been extremely successful at identifying variants and genes associated with common human diseases and other complex traits. The vast majority of studies have used one or both of two statistical methods for trait association: common variant association using single marker tests (SMT) and rare variant association using gene-based aggregation tests (Lee et al. 2014; Povysil et al. 2019). However, neither method is well suited for identifying rare variant associations in noncoding regions where most causal variants are known to reside (Maurano et al. 2012), or for testing the combined association of multiple independent common and/or rare variant signals (potentially with opposing effects) in cases of allelic heterogeneity (i.e., when multiple causal variants are present at a locus). Region-based methods have been adapted to noncoding regions using sliding window approaches, with some success (Li et al. 2019), but this approach is limited by two major challenges. First, it is difficult to decide which intervals and sets of variants to test in noncoding regions where knowledge of variant function is limited. Second, including nonfunctional variants in these tests can greatly reduce power.

In theory, genealogy-based methods that seek to associate local ancestral clades with traits have the potential to overcome these limitations through their ability to combine independent and potentially opposing signals present in different regions of the local ancestral tree, without the need to define functional regions or variant sets. Despite notable early progress (Zöllner and Pritchard 2005; Minichiello and Durbin 2006), these methods have proven difficult to implement in practice due to the computational challenges of genome-wide population-scale haplotype inference and the statistical challenges of tree-based association testing. Recent advances in haplotype inference have eased the computational burden of building local genealogies (Rasmussen et al. 2014; Zhang et al. 2023; Speidel et al. 2019; Kelleher et al. 2019; Aslett and Christ 2024), making genome-wide trait association studies feasible (albeit still computationally expensive). Building on this, several new methods have recently been developed to test genome-wide local genealogies for trait association.

The first, ARG-Needle (Zhang et al. 2023), builds local genealogies using a scalable ARG-based algorithm, samples a set of inferred clades that may harbor an unobserved variant, and adds the genotypes corresponding to those clades to the list of genotypes tested in genome-wide SMT. Although ARG-Needle is an extremely powerful method for reference free imputation, this “inferred variant” SMT approach is not designed to combine independent genetic signals at loci with allelic heterogeneity.

A second method (Link et al. 2023) uses the Li and Stephens (LS) model HMM implemented in Relate (Speidel et al. 2019) to generate local expected genetic relatedness matrices (eGRMs) which are then tested for association with the phenotype. Link et al. showed via simulations that this approach can boost power when multiple independent causal variants are present at a locus, and their analysis of two chromosomes in a Native Hawaiian cohort of 5,384 individuals found evidence for a robust (albeit not genome-wide significant) BMI association signal with allelic heterogeneity. A more recent method (Gunnarsson et al. 2024; Zhu et al. 2024) uses a highly scalable local ancestry inference engine and a statistical testing approach similar to Link et al. (Link et al. 2023), and has been applied to gene-based trait association in the UK Biobank. However, since both of the above methods are based purely on a quadratic form test statistic, they struggle against the enormous multiple-testing burden incurred by attempting to test all of the clades in a local tree, genome-wide.

Our method, LOCATER (Christ et al. 2025), employs a novel three-step procedure to test for the association of local genealogical relationships with traits, including a traditional single marker test (SMT), the newly developed Stable Distillation (SD) association test (Christ et al. 2022), and a quadratic form (QForm) association test similar to the method used in Link et al (Link et al. 2023). SD is a specialized association test that is much more powerful than quadratic form testing at assessing the combined effects of many small clades marked by ultra-rare variants. When used in conjunction with an optimized implementation of the LS model we developed to infer local ancestry, kalis (Aslett and Christ 2024), our simulation studies have shown that LOCATER outperforms SMT in cases of allelic heterogeneity, that this advantage is more pronounced as the number of causal variants increases and their allele frequency decreases, and that the SD sub-test is the primary driver of power gains (Christ et al. 2025).

Recent evidence suggests that allelic heterogeneity is widespread in humans. For example, a recent massive parallel reporter assay (MPRA) study estimated that 10%∼20% of expression quantitative trait loci (eQTLs) have multiple causal variants in humans (Abell et al. 2022), and a previous study showed that by inference, the proportion of all loci with allelic heterogeneity is 4 to 23% in eQTLs, 35% in GWASs of high-density lipoprotein (HDL), and 23% in GWASs of schizophrenia (Hormozdiari et al. 2017). These and related observations (GTEx Consortium 2020) suggest that methods such as LOCATER that leverage allelic heterogeneity to improve power have the potential to discover novel trait associated alleles and genes not found by other methods.

Here, we use LOCATER to screen for trait-associated loci in the METSIM cohort. METSIM is a population sampled cohort of Finnish men with whole-genome sequencing data and a large number of cardiometabolic traits. Prior genome-wide association studies in METSIM have mapped many loci associated with cardiometabolic traits and disease, including studies based on array-based genotype data (Karjalainen et al. 2024; Davis et al. 2017), exome sequencing data (Locke et al. 2019), and whole-genome sequencing data (Chen et al. 2021; Ganel et al. 2021). Due to a recent population bottleneck and subsequent expansion, genetic diversity is somewhat reduced in Finland and there is a larger fraction of deleterious variants at intermediate allele frequencies, facilitating trait mapping at relatively modest sample sizes (Manolio et al. 2009; Locke et al. 2019). These features, coupled with extensive prior knowledge of Finnish genetics, make this an ideal cohort to test new trait mapping methods such as LOCATER.

## RESULTS

The LOCATER method has been described in detail and tested on simulated data in separate work (Christ et al. 2025). Briefly, LOCATER involves a 3-step process, the first of which is to run local ancestry inference at each genetic marker. LOCATER uses an optimized version of the LS model and represents (Speidel et al. 2019). The second step is to identify small clades, which we refer to as “sprigs”. Sprigs are independent predictors that were derived from the smallest possible inferred clades. LOCATER tests discretely called clade genotypes in X(ℓ) and clades encoded in the local relatedness matrix in Ω(ℓ). As in Christ et al. (Christ et al. 2025), we use X(ℓ) to only encode very small clades (sprigs): each typically has at most 10 haplotypes under it. LOCATER includes a sprig calling algorithm that clusters the genomic distances and calls sprigs with a greedy clique finding procedure. Then, at each genetic marker, LOCATER performs three association tests sequentially: (1) a standard SMT to measure the contribution of that specific marker; (2) Stable Distillation (SD) to measure the contribution of the inferred small clades based on inferred local ancestry; and (3) a quadratic-form based test (QForm) to measure the contribution of remaining ancestral relationships (Christ et al. 2025). Steps 2 and 3 use a residualized phenotype vector from the prior step. This, combined with the independence guarantees of SD (Christ et al. 2022), ensures that the resulting three p-values from SMT, SD, and QForm are mutually independent under the null hypothesis. We then combine the three p-values using an adapted version of Fisher’s method that we call Maximizing over Subsets of Summed Exponentials (MSSE) (Christ et al. 2025). Multiple optimizations for computing efficiency were implemented throughout the LOCATER pipeline, with more details in Christ et al. (Christ et al. 2025) and the Methods section.

Applying LOCATER in this study required overcoming two key methodological challenges that were not addressed in our prior simulation-based work (Christ et al. 2025): tuning our ancestry inference engine for use with real-world whole-genome sequencing (WGS) data, specifically from the METSIM cohort in Finland, and fully accounting for the effects of cryptic confounders in haplotype screening. Although the specific details of these analysis steps will need to be worked out for any new association study that uses LOCATER or similar methods on a new dataset, the approaches we describe below may provide a general solution that helps guide future implementation of these methods.

### Parameter tuning for local ancestry inference

The LOCATER pipeline used for our prior and current work uses the Speidel version (Speidel et al. 2019) of the LS model implemented in kalis (Aslett and Christ 2024), which includes recent developments to improve efficiency for genome-wide testing (Christ et al. 2025). As described in Aslett & Christ (Aslett and Christ 2024), the LS model in kalis has two parameters: the effective population size parameter (N_e_), and the mutation probability (𝜇). Since N_e_ acts by rescaling the provided recombination map in the LS model (Equation 3, Aslett and Christ 2024), we will refer to −log_10_N_e_ as the recombination penalty parameter. Analogously, we will refer to −log_10_μ as the mutation penalty parameter.

To best represent local genealogy structure and propagate association signals, tuning these parameters has been a focus of recent work on the LS model (Speidel et al. 2019; Jin and Terhorst 2023). Rather than using the data likelihood and expectation-maximization (EM) (Baum et al. 1970) to select these two parameters for our METSIM analysis, Relate (Speidel et al. 2019) proposes using a more relevant objective aimed at maximizing the performance of the LS model for their specific purpose: capturing local variants in their inferred ancestral trees. Here, we propose an objective aimed at optimizing the discovery power of LOCATER and other methods aiming to leverage allelic heterogeneity. At a given locus, LOCATER works on matrices representing inferred local genealogy derived from the modified LS model, and LOCATER boosts the power of SMT by applying Stable Distillation (SD) and a quadratic form test (QForm) to the local genealogy representations (Christ et al. 2025). Thus, we selected HMM parameters that optimize the propagation of nearby signals using the following objective function and sampling scheme.

We randomly sampled many genomic regions from the dataset and assigned a causal variant and a target variant that are 0.05 cM away (see Methods for details). We then simulated a phenotype vector with a strong effect driven by the causal variant and ran LOCATER at the target variant to measure how well it could capture the signal driven by the nearby causal variant. We calculated the relative signal as our metric, defined as

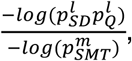

where 𝑙 is the index of the target variant and 𝑚 is the index of the causal variant. 𝑝*_SMT_* refers to p-values returned by SMT, 𝑝*_SD_* and 𝑝_Q,_ refer to p-values returned by the SD and QForm sub-tests in LOCATER, respectively. This objective focused on association signal propagation could be used to train parameters for any association method targeting loci with allelic heterogeneity.

The trimmed mean surface for this relative signal showed that multiple parameters have comparable efficiency and formed a plateau (Figure S1) (See Methods, see Table S3 for HMM parameters evaluated). Our result aligns with a similar finding in Speidel et al. (Speidel et al. 2019) that high mutation penalties combined with low recombination penalties are not well suited for haplotype inference. After ruling out parameters that require a much longer time to run, we randomly picked one parameter (recombination penalty of 6 and a mutation penalty of 8) on the plateau of the surface, averaging across allele frequency bins. We also generated surfaces for different allele frequency bins and confirmed that the shape of the surfaces remains consistent across allele frequency bins for both the QForm and SD association methods.

### Methodological improvements to the LOCATER pipeline to account for cryptic confounders

In our initial trait association experiments we observed deviations in the empirical p-value distributions from LOCATER, with cases of both mild inflation and mild deflation across the 101 traits analyzed in this study, despite using standard procedures to correct for population structure using principal component analysis (PCA). In contrast, we did not observe inflation when running SMT on the exact same data, nor did we observe inflation when running LOCATER on simulated phenotype data. This suggests that genealogy-based trait association methods are especially sensitive to the effects of cryptic confounders. We believe that this is due to the fact that there is a much greater degree of correlation between nearby genetic markers for tree-based tests than for SMT, and that this causes a much larger fraction of markers to be affected by confounders. Notably, this implies that these confounders are also affecting the SMT results, just in a less readily detectable way.

To more precisely calibrate LOCATER results, we took inspiration from the phenotype rank-matching procedure used in LOCATER. By default, LOCATER normalizes phenotypes by mapping ranks to simulated Gaussian random variables rather than to fixed quantiles (Christ et al. 2025). This approach avoids the subtle dependence induced when mapping to fixed quantiles. Under the assumption that the original residuals are exchangeable, matching to simulated Gaussian random variables indeed yields independent Gaussian phenotypes, which is assumed by the SD procedure underlying LOCATER.

Building on this procedure, we repeated the rank-matching process for the same phenotype several times, each version of the phenotype vector is a subtly different perturbation of the original phenotype.

We found that when we ran LOCATER across these different perturbations, the amount of inflation observed via Q-Q plots differed moderately across perturbations. We believe this variation reflects the fact that, by chance, different perturbations can have stronger or weaker correlations with confounding factors such as population structure. Thus, the rank-matching procedure allows us to modulate the correlation between the phenotype and confounding variables such that we can identify a rank-matched version that is the least correlated with confounders. We therefore rank matched each phenotype 100 times and chose the version that showed the least alignment with confounders for our modified genomic control (GC) (see Methods). For each version of each phenotype, we conducted an association test for ∼30,000 evenly spaced variants and selected the version for which the p-value distribution is the closest to a uniform distribution for both SD and QForm (see Methods). We found that SMT p-values of all versions are consistently well calibrated, and SD and QForm p-values from different versions will have substantially different deviations from the theoretical null (Figures S2, S3 and S4, column “Rank matched chosen”, “Rank matched 2” and “Rank matched 3”). We also found that the rank-matched phenotype chosen for the association study indeed has the least deviation and eliminated the inflated body of the QForm p-value distribution. Our study is the first to report genome-wide Q-Q plots for a genealogy-based association method and demonstrate calibration of that method after rigorously adjusting for unobserved confounding factors.

We then sought to apply a more general genomic control to the p-values. In short, we fit a line to the Q-Q plot of LOCATER −log_10_p of the selected rank-matched phenotype. We applied modified GC to p-values using the slope and intercept of the fitted line (see Methods). This is similar to traditional genomic control (Devlin and Roeder 1999), which only fits a slope to the Q-Q plot. By incorporating a non-zero intercept, we can achieve a much more accurate fit to the tail of our null distribution of -log_10_p. In order to gain a better intuition for the role of this intercept, it is helpful to think in terms of a simple application like single-marker testing where each p-value corresponds to a Z-score. In this setting, fitting a slope via classic genomic control to the Q-Q plot is equivalent to fitting a scale parameter to the distribution of null Z-scores while keeping the location of that null distribution fixed at zero. Introducing a non-zero intercept allows one to fit a null distribution to the Z-scores where the location may also be adjusted. A similar procedure is described in Chapter 6 of Efron (Efron 2010) and is standard in large-scale testing problems where the null assumptions are not satisfied (e.g., when there is confounding). In Figures S2, S3 and S4, we applied this modified GC procedure to p-value distributions from LOCATER-specific tests, and all distributions align with the expected distribution much better (Figures S2, S3 and S4, row “SD modified GC” and “QForm modified GC”).

### GWAS of 101 traits

We then performed an association analysis of 6,795 METSIM individuals with 101 quantitative metabolic phenotypes (see Table 1 for highlighted associations) using LOCATER. We also analyzed these traits with SMT as our benchmark. All traits were residualized based on trait-specific covariates beforehand, exactly as described in our prior exome based study of this same cohort (Locke et al. 2019). After filtering out indels and variants lacking ancestral allele annotation (see Methods), we tested 18,949,137 autosomal variants. We used the top 10 Finnish-specific principal components as background covariates for the association test. As mentioned above, during the pre-screening stage we selected the phenotype vector perturbation that yielded the most calibrated Q-Q plot (see Methods). Post-screening we applied modified GC to the resulting p-values based on the slope and intercept (as described above, see Methods) in the Q-Q plot to ensure that both SD and QForm sub-tests in LOCATER were well-calibrated.

**Table 1:** Summary of highlighted associations. Chromosome positions are based on GRCh38. See Table S1 for trait descriptors, see Table S2 for full results. The genome-wide significance threshold is 7.17 x 10^-9^. To allow for a straightforward comparison, the LOCATER p-value was standardized to match the effective number of independent tests for single marker association. * denotes the associations that SMT lead marker is the same as LOCATER lead marker.

| Variant ID | Mapped Gene | Trait | LOCATER P (modified GCed) | LOCATER lead marker MAF | Variant ID of SMT lead marker | SMT P | SMT lead marker MAF | Known hit in GWAS catalog | SMT Lead marker LD w/ GWAS catalog marker |
| --- | --- | --- | --- | --- | --- | --- | --- | --- | --- |
| Chr7-73440219-C-T | *FZD9*, *BAZ1B* | MUFA | 1.57E-09 | 0.096 | Chr7-73440219-C-T* | 2.52E-08 | 0.096 | Yes | 0.701 |
| Chr7-73482065-A-C | *BAZ1B* | Triglycerides in VLDL | 1.64E-11 | 0.12 | Chr7-73467477-C-T | 1.11E-09 | 0.12 | Yes | 0.852 |
| Chr7-73643687-A-G | *VPS37D*, *MLXIPL* | Concentration of large VLDL particles | 2.82E-09 | 0.12 | Chr7-73641131-A-C | 3.40E-08 | 0.12 | Yes - related trait | 0.933 |
| Chr11-61843278-G-A | *FADS2* | HDL2-C | 6.58E-09 | 0.43 | Chr11-61798436-T-C | 1.65E-08 | 0.45 | Yes - related trait | 0.871 |
| Chr11-105327888-G-C | *CARD18* | Triglycerides in medium VLDL | 2.31E-09 | 0.43 | Chr11-105331384-A-T | 1.05E-05 | 0.43 | No | NA |
| Chr18-49653146-G-A | *ACAA2*, *SMUG1P1* | Triglycerides in medium HDL | 1.68E-09 | 0.15 | Chr18-49642278-G-A | 4.71E-07 | 0.16 | Yes | 0.864 |
| Chr18-49817040-T-A | *SNHG22* | ApoA1 | 6.51E-09 | 0.0041 | Chr18-49817040-T-A* | 2.15E-08 | 0.0041 | Yes | 1 |
| Chr19-44922203-A-G | *APOC1*, *APOC1P1* | Remnant-C | 7.53E-11 | 0.29 | Chr19-44922203-A-G* | 1.40E-09 | 0.29 | Yes | 0.0783 |
| Chr20-45906012-G-A | *PLTP* | Triglycerides in small HDL | 1.80E-11 | 0.23 | Chr20-45923216-T-C | 3.10E-10 | 0.18 | Yes | 1 |
| Chr21-16318536-C-T | *MIR99AHG* | HDL3-C | 3.87E-09 | 0.090 | Chr21-16318536-C-T* | 1.77E-08 | 0.090 | No | NA |

In terms of computational performance, we ran LOCATER using a total of 4,780 jobs, each of which was assigned 12 cores and 60 GB of memory in an academic high-throughput computing system. On average, each job took ∼32 minutes. This shows the feasibility of running LOCATER at scale on real-world datasets. However, we note that LOCATER runtimes are sensitive to the number of genomic loci that show promising p-values during the initial screening procedure, and will vary on a dataset-by-dataset basis.

Considering that this genome sequencing dataset contains an abundance of rare variants, applying the canonical genome-wide significance threshold (5 x 10^-8^), which assumes one million independent tests, is not appropriate. We used permuted phenotypes to estimate the effective number of independent tests (Gao et al. 2008; Hoh et al. 2001) (see Methods). Based on the distribution of minimum p-values, we estimated that the effective number of independent tests for SMT (𝑇*_SMT_*) is 6,977,438, and for LOCATER (𝑇*_LOCATER_*) is 3,551,616 (α= 0.05). Based on 𝑇*_SMT_*, the genome-wide significance threshold for SMT should be 7.17 x 10^-9^. Note that LOCATER and SMT used the exact same set of variants, but that the p-values reported by LOCATER are more dependent across loci due to shared local genealogies. For a clearer comparison with SMT in visualizations (e.g., Fig. 2A), we further standardized the LOCATER p-value with

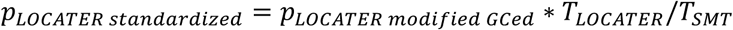

to make the LOCATER genome-wide Bonferroni threshold match that of SMT (7.17 x 10^-9^). This does not change the interpretation of LOCATER results since both the results and the significance cutoff were rescaled to the same extent. To enable a direct comparison between SMT and LOCATOR, all LOCATER results reported below are standardized.

After the screening, we identified loci of interest as genomic regions with one or more variants with significant p-values by either LOCATER or SMT in any of the 101 traits. To compare LOCATER and SMT signals at these loci, we defined a common shared association interval by merging the set of significantly associated variants identified by either method, where the merging includes a 600 kb flanking region for each variant. For convenience, we used a similar process to merge association intervals across traits to obtain a nonredundant set of genomic regions, although in this case, we note that independent signals for different traits may often be lumped together (see Table S2 for the full set). Altogether, we identified 47 genomic regions with 351 associations across all traits.

When comparing the most significant signal for LOCATER and SMT at every identified association (identified as min(𝑝_-./0*12 45678698:;<8_) < threshold or min(𝑝_()*_) < threshold), we found that LOCATER and SMT identified many associations together (327 out of 351, Figure 1D). Many of these are in canonical regions known to be associated with cardiovascular diseases, such as *PCSK9*, *APOB*, *LPL*, *LIPC*, *CETP*, and the *APOE/C1/C4/C2* gene cluster. A small number of associations are significant only in LOCATER (7 out of 351), and SMT found 17 associations that LOCATER did not (Figure 1B).

**Figure 1.**
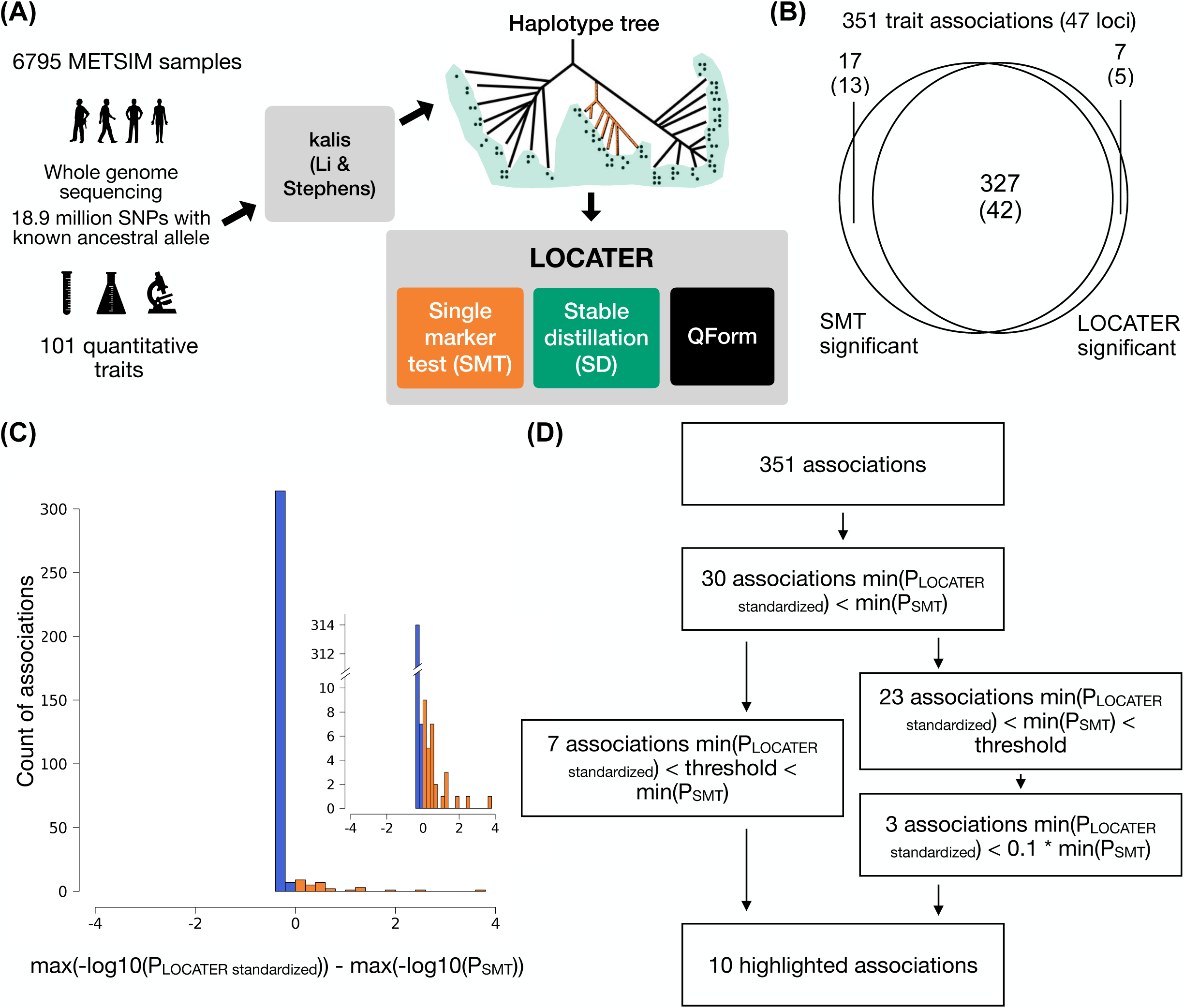
Schematic of our genealogy-based screening procedure, LOCATER, and summary of screening results. (A) Diagram of the experimental design and association testing method. Kalis is an implementation of the Li & Stephens model for local ancestry inference. QForm: quadratic form testing. (B) Venn diagram showing the number of associations and number of loci (in parentheses) with significant SMT and/or significant LOCATER results. Note that a given locus may have distinct associations represented in different parts of the Venn diagram. (C) Distribution of max(-log10(P_LOCATER standardized_)) - max(-log10(P_SMT_)) for all 351 associations. All associations that are genome-wide significant by either SMT or LOCATER are included. (D) Overview of significant associations and highlighted associations. The genome-wide significance threshold is 7.17 x 10^-9^ for SMT and standardized LOCATER.

**Figure 2.**
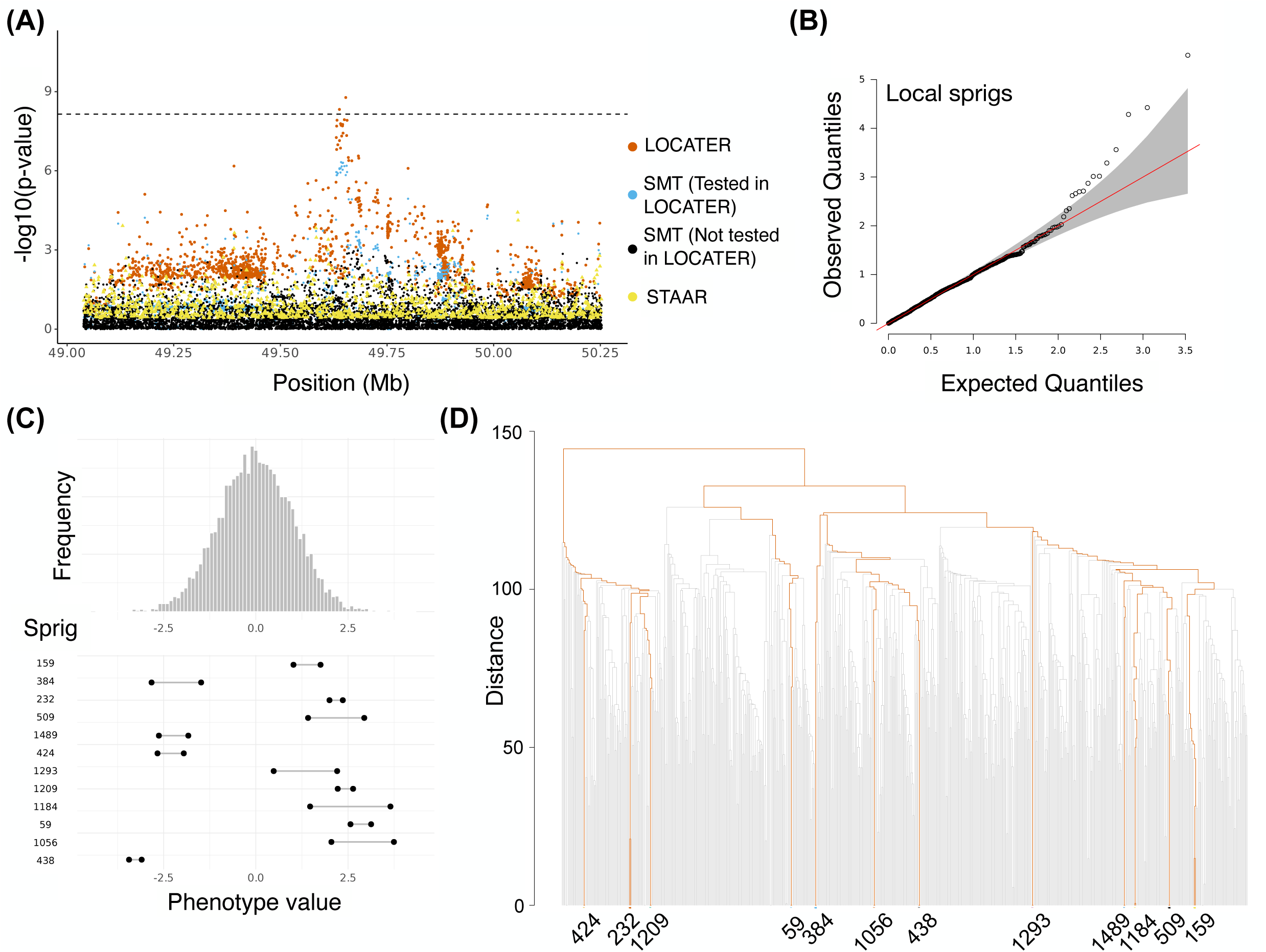
Association of triglycerides in medium HDL at LIPG locus. **(A)** Local Manhattan plot of the association signal for “triglycerides in medium HDL” on chr18:49038347-50253146, including results for single marker test (SMT; blue and black), LOCATER (orange) and STAAR (yellow). Note that LOCATER results are only shown for variants with an SMT p-value less than 1×10^-3^, since for computational efficiency only these variants were tested by LOCATER (see Methods). SMT results from variants tested by LOCATER are shown in blue, and those from variants not tested by LOCATER are shown in black. The black dashed line corresponds to the genome-wide significance threshold for SMT, standardized LOCATER, and standardized STAAR. **(B)** Q-Q inflation plot of −log10(p-values) from all “sprigs” at the lead marker chr18:49653146, where “sprigs” are defined as the smallest possible inferred clades. The gray area corresponds to the 95% confidence interval, and the red line denotes x=y. **(C)** Histogram of phenotype values after projecting out the genotype vector of the LOCATER lead marker (chr18:49653146), thus removing signal that can be accounted for by the SMT sub-test. Connected dots show the phenotype value of individuals assigned to significant sprigs. **(D)** Dendrogram generated from the haplotype-level local distance matrix at the lead marker chr18:49653146. The UPGMA method was used for hierarchical clustering. Orange branches highlight the path of all haplotypes in significant sprigs shown previously in **(C)**. Labels at the bottom show the sprig assignment. For plotting clarity, 95% of haplotypes under insignificant sprigs were pruned.

Notably, in the cases where SMT is more significant than LOCATER (321 out of 351 associations), it is typically by a very small margin, whereas in the cases where LOCATER is more significant (30 out of 351), it is typically by a relatively substantial margin (Figure 1C). Our interpretation of this result is that LOCATER has less power than SMT at trait associations resulting from a single causal variant because of the statistical penalty incurred by attempting to incorporate nearby signals, but that LOCATER greatly outperforms SMT at loci with multiple causal variants. In total, 5 of the 47 significant loci (10.6%) show a signal boost from LOCATER, indicating that allelic heterogeneity is fairly common, even in a relatively small sample of the Finnish population that is known to be depleted of genetic diversity relative to most other human populations due to historical bottlenecks.

For the 30 associations with a more significant signal from LOCATER, we inferred the number of independent causal variants based on the Stable Distillation (SD) and quadratic form (QForm) sub-test signals from LOCATER (Table S5). These 30 associations reside in 13 distinct genomic regions. After clumping associations that reside in the same genomic region (with 600 kb flanking regions) and involve traits that are highly correlated to each other (r^2^>0.8) in our dataset, there were 21 nonredundant associations. Of these, the LOCATER signal boost came from SD in 15 cases, and from QForm in 6 cases.

Of the 15 nonredundant associations that were boosted by SD, 7 of them were boosted by only one sprig, while the other 8 of them had 2-12 significant sprigs contributing to the signal. All significant sprigs represent distinct haplotype groups in the local ancestry trees. As a result, the total number of inferred causal variants for these 15 associations ranges from 2 to 13. We also report the variants that are completely linked with significant sprigs in Table S4. For the 6 associations boosted by QForm, we used iterative conditional analysis to infer the number of causal variants. One association became insignificant after accounting for the lead marker, and signals in 5 other associations were diminished after multiple (2-5) rounds of conditional analysis, suggesting that the total number of inferred causal variants for these six associations ranges from 1-5.

Rare variant association methods such as STAAR (Li et al. 2020) also seek to assess the combined effects of multiple causal variants at a locus, albeit using a very different approach than LOCATER. To test whether STAAR could potentially detect the same signals as LOCATER, we ran STAAR on the 30 associations mentioned above for which LOCATER had a more significant p-value than SMT. Notably, STAAR did not detect any of these 30 associations at genome-wide significance. A caveat to this analysis is that there are many different ways to run STAAR based on window size, variant inclusion, variant annotation and weighting criteria, and so we cannot discount the possibility that STAAR might be able to detect some of these signals. For example, if STAAR is provided with variant impact scores from CADD (Rentzsch et al. 2019), it is able to identify 4 significant associations at the LIPC locus, albeit with far less significant p-values than SMT and LOCATER (>30 orders of magnitude difference). These results suggest that the trait association signals detected by LOCATER are not easily captured by current rare variant association methods such as STAAR. This is perhaps not surprising given that such methods are designed and primarily used for gene-based association, where the target interval is well defined and variant annotations are much more informative.

Below, we discuss some of the trait associations detected by LOCATER. We highlight five known association signals that LOCATER detected but SMT did not, three cases where both LOCATER and SMT detected the association but LOCATER provided a substantial boost in signal strength, and two potentially novel association signals detected solely by LOCATER.

### LOCATER recovers known associations at the *LIPG* locus

LOCATER recovered several known quantitative trait loci that would otherwise have been missed by SMT in our present study (see Figure 1B), an example being the *LIPG* locus. *LIPG* encodes a well-known member of the triglyceride lipase family of proteins and is primarily involved in the metabolism of HDL (Jaye et al. 1999; Jin et al. 2003; Ma et al. 2003; Strauss et al. 2002). LOCATER recovered genome-wide significant associations for triglycerides in medium HDL (𝑝=1.68 x 10^-9^) and apolipoprotein A-I (𝑝=6.51 x 10^-9^), the major protein component of HDL particles. These two trait associations are likely to be independent given that the lead markers are in low LD (r^2^ of 7.29 x 10^-4^) and the two traits are not significantly correlated (Pearson correlation of 0.132), which is consistent with prior work (Davis et al. 2017). Neither of these associations was captured by SMT at genome-wide significance. The smallest SMT p-value for triglycerides in medium HDL within a 1.2 Mb window of *LIPG* was 4.71×10^-7^, and that for apolipoprotein A-I was 2.15×10^-8^ (Table 1). The lead variant for apolipoprotein A-I was also found in a prior study of Finns (𝑝=2 x 10^-10^) using many of the same METSIM samples analyzed here (Davis et al. 2017), and the lead variant for triglycerides in medium HDL was found in a large study of 233 metabolic traits in 33 cohorts (Karjalainen et al. 2024).

We first discuss the *LIPG* association with triglycerides in medium HDL, where LOCATER detected a significant signal but SMT did not (Figure 2A). LOCATER’s improved power over SMT in this case comes from the Stable Distillation (SD) sub-test (Figure S5C, D), which indicates contributions from multiple ultra-rare causal variants. We confirmed that the p-value distribution after modified GC aligns very well with the expected distribution, and the QQ-plot-based modified GC required to control confounding was minimal (Figure S5A, B).

The LOCATER SD sub-test has the advantage that p-values from all predictors (sprigs, defined as the smallest possible inferred clades) are independent under the null hypothesis. A Q-Q plot of all -log_10_ sprig p-values calculated at the lead marker shows that the top 12 sprigs significantly deviated from the expected distribution and thus contributed to the SD signal (Figure 2B). To highlight the coalescent path of significant haplotypes, we plotted a dendrogram of haplotypes based on hierarchical clustering of the similarity matrix at the lead marker, which showed that different significant sprigs were present within distinct clades in the local coalescent tree, and that haplotypes in the same sprig were very similar. Haplotypes in the same sprig have a very recent coalescence, and haplotypes from different sprigs have a much more distant coalescence (Figure 2D). This suggests that the SD sub-test captured signals from multiple distinct haplotype groups rather than multiple signals driven by a single variant.

Notably, SD is able to combine signals from individuals at both extremes of the phenotype distribution, which correspond to alleles with opposing effects. As expected, all samples within significant sprigs have phenotype values that are far from the mean (Figure 2C). Individuals within each sprig resided on the same side of the distribution, but different sprigs could reside on different sides.

We next sought to visualize variants influencing the SD signal at this locus using residual analysis. The first phase involved conducting SMT based on a phenotype that projected out the lead marker genotype vector, removing its effect on the signal. We defined the p-values resulting from this experiment as 𝑝*_S_*. The second phase involved performing SMT based on the residualized phenotype orthogonal against the lead marker SMT and SD signal, yielding a second set of p-values defined as 𝑝_+_. The difference between 𝑝*_S_* and 𝑝*_D_* shows the contribution of genomic variants to the SD signal. We plotted -log_10_𝑝*_s_* and -log_10_𝑝*_D_* on a Manhattan plot (Figure S5E) and a scatter plot (Figure S5F), highlighting variants where 𝑝*_S_* < 1 x 10^-3^ and 𝑝_+_ > 10 * 𝑝*_S_*. This experiment shows that SD captured signals from many variants scattered in an extensive genomic region (> 10 Mb), supporting the success of our ancestry inference model tuning procedure.

By plotting principal components (PCs), we confirmed that SD-significant individuals do not form a tight cluster in any specific area of the plot, so this association signal was not obviously confounded by population structure (Figure S5G).

We now turn to the second *LIPG* association, apolipoprotein A-I. LIPG is known to regulate serum apolipoprotein A-I (Jaye et al. 1999; Ishida et al. 2003). A previous study in METSIM (Davis et al. 2017) showed that our lead marker is associated with five HDL subclass traits and apolipoprotein A-I (Figure 3A), and in this study, LOCATER recovered the signal in apolipoprotein A-I, but SMT missed it.

**Figure 3.**
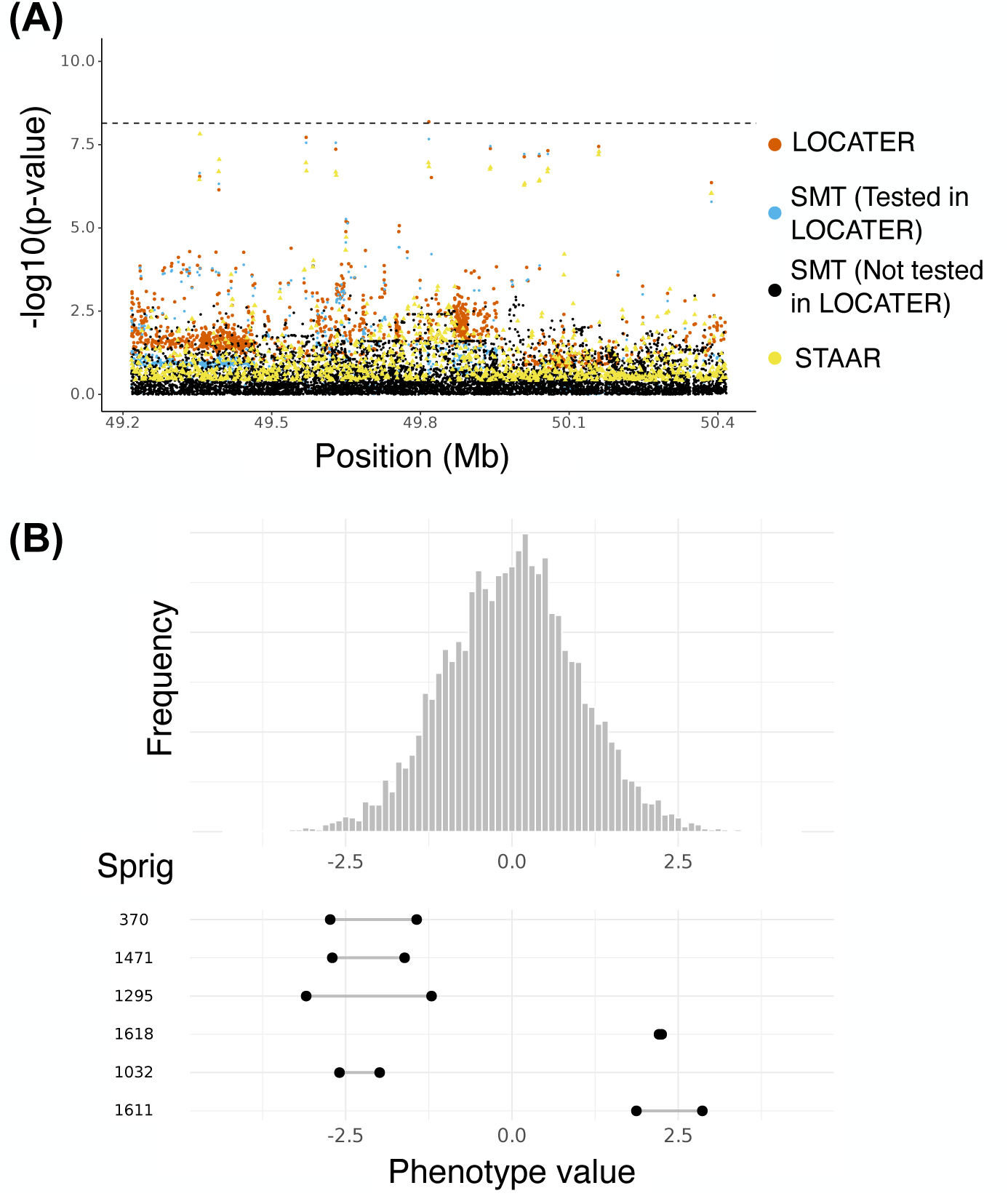
Association of apolipoprotein A1 at LIPG locus. **(A)** Local Manhattan plot of the association signal for apolipoprotein A1 on chr18:49217040-50417040, shown using the exact same data types and color scheme as Figure 2A. **(B)** Histogram of phenotype values after projecting out the genotype vector of the LOCATER lead marker (chr18:49817040), thus removing signal that can be accounted for by the SMT sub-test. Connected dots show the phenotype value of individuals assigned to significant sprigs.

LOCATER gained its advantage over SMT from SD (Figure S6A). The Q-Q inflation plot of sprig -log_10_ p-values showed that 6 sprigs contributed to the signal (Figure S6B). Similar to the first *LIPG* association result discussed above, haplotypes from the same sprig have a very recent coalescence and those from different sprigs have a more distant coalescence (Figure S6C), and samples within outlying sprigs had phenotypes that were far away from the median and on different sides of the distribution (Figure 3B).

In summary, LOCATER was able to discover two independent trait associations at *LIPG* based on the presence of allelic heterogeneity, where both trait associations were missed by SMT. In addition to these two examples at *LIPG*, three additional trait associations at other known loci were also detected by LOCATER but not by SMT. The association with HDL2 cholesterol on Chr11 (Figure S13) and the association of monounsaturated fatty acids (MUFA) on Chr7 (Figure S9) were also identified based on SD signals and also showed consistent phenotype values across relevant individuals. Both associations were boosted by only one sprig, and phenotype values of individuals assigned to significant sprigs are all far from the mean of the corresponding phenotype distribution. There was also an association with ’large VLDL particle concentration’ on chromosome 7 (Figure S11) that was boosted by QForm; however, this association is somewhat less confident than others given that only one variant crossed the significance threshold and that a substantial post-hoc adjustment for confounding was required to calibrate the corresponding QQ-plot.

### A potentially new association on Chr11

LOCATER found an association of ’triglycerides in medium VLDL’ on chromosome 11, while SMT did not (Figure 4A), and the GWAS catalog (Sollis et al. 2023) did not report any known association with correlated traits. LOCATER was much more significant than SMT due to the SD sub-test, implying a contribution from ultra-rare haplotypes (Figure S7C). The SD signal was distributed evenly across the entire ∼1 Mb locus (Figure 4B). Among all sprigs called at the lead marker position, the top five sprigs significantly deviated from the expected distribution (Figure 4C). The phenotype distribution of individuals in significant sprigs showed that they are outliers, and that trait outliers are found at both extremes of the distribution (Figure 4D). The dendrogram from hierarchical clustering of the local similarity matrix, with highlighted coalescent paths for significant haplotypes, showed that the signal was not coming from a larger clade but rather from multiple distinct small groups of haplotypes (Figure 4E). These results suggest that LOCATER is combining association signals from 5 rare haplotype groups with distinct genealogical histories, while these signals were not detectable by standard SMT.

**Figure 4.**
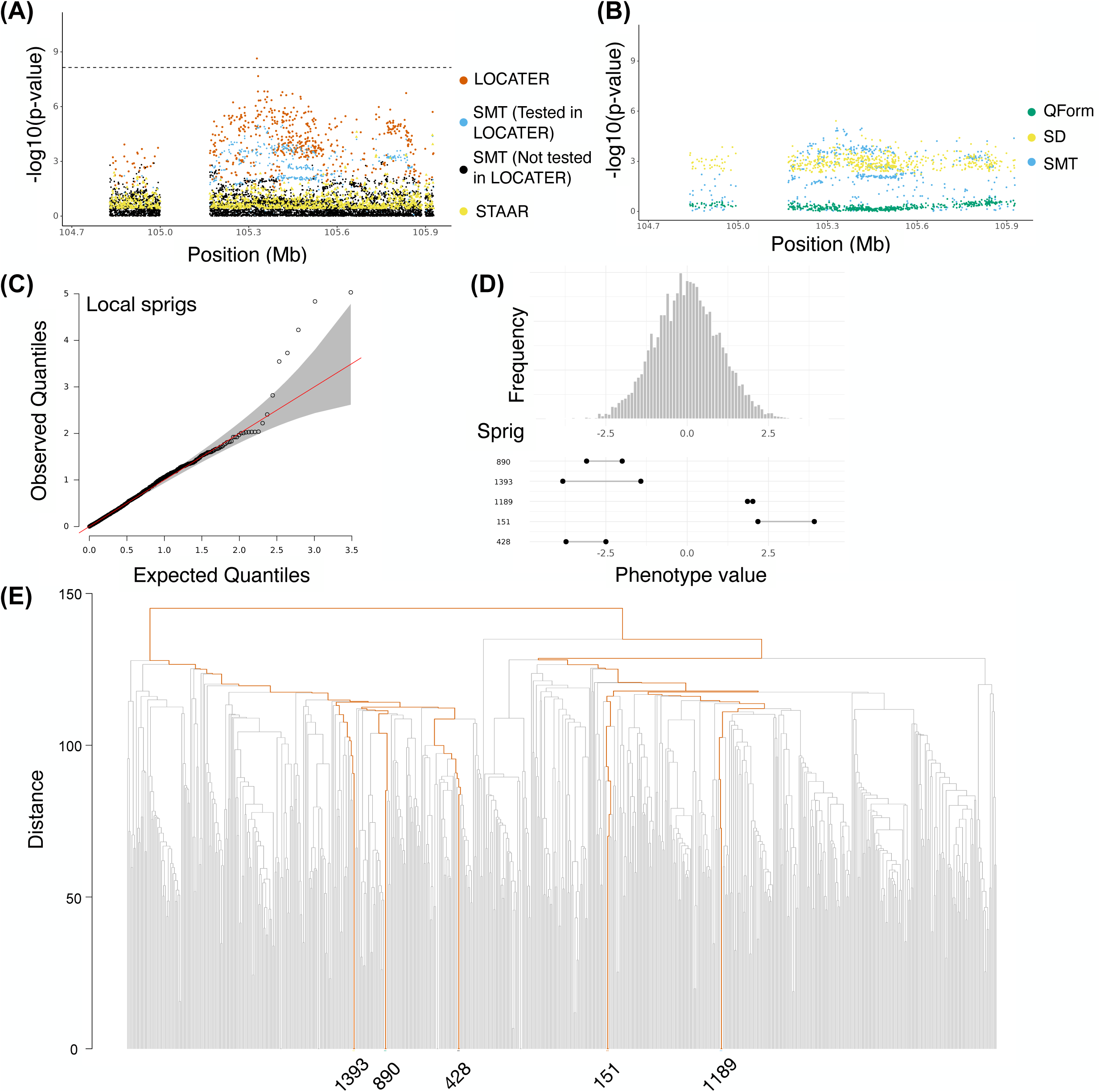
Association of triglycerides in medium VLDL on chr11. **(A)** Local Manhattan plot of the association signal for “triglycerides in medium VLDL” on chr11:104727888-105927888, shown using the exact same data types and color scheme as Figure 2A.**(B)** Local Manhattan plot of “triglycerides in medium VLDL” at chr11:104727888-105927888, showing modified genomic controlled -log_10_(P) for the 3 LOCATER sub-tests. **(C)** Q-Q inflation plot of −log10(p-values) from all “sprigs” at the lead marker chr11:105327888, where “sprigs” are defined as the smallest possible inferred clades. The gray area corresponds to the 95% confidence interval, and the red line denotes x=y. **(D)** Histogram of phenotype values after projecting out the genotype vector of the LOCATER lead marker (chr11:105327888), thus removing signal that can be accounted for by the SMT sub-test. Connected dots show the phenotype value of individuals assigned to significant sprigs. **(E)** Dendrogram generated from the haplotype-level local distance matrix at the lead marker chr11:105327888. The UPGMA method was used for hierarchical clustering. Orange branches highlight the path of all haplotypes in significant sprigs shown previously in part **(D)**. Labels at the right show the sprig assignment. For plotting clarity, 95% of haplotypes under insignificant sprigs were pruned.

We performed residual analyses to investigate the contribution of genomic variants to the SD signal and noticed many variants with drastically different 𝑝_(_ and 𝑝_+_ (i.e., substantial contribution to the SD signal). These variants extend across a mega-base long genomic distance, highlighting LOCATER’s ability to merge sub-significant signals from a large genomic region.

We investigated this association and found that the lead marker resides in a predicted transcription factor (TF) binding site for KLF16 and KLF9. KLF9 is a metabolic TF acting in the liver, where its increased expression promotes gluconeogenesis and alters glucose and lipid homeostasis (Cui et al. 2019). The adjacent 1.2 Mb region includes CASP1, CASP4, CASP5, CASP12, CARD18, and CARD16. CASP1 (encoding Caspase-1) has a core role as the central effector of the canonical inflammasome, and there is evidence from mouse studies that CASP1/inflammasome signaling can raise circulating TGs by increasing hepatic VLDL/apoB secretion, and may also impair TG clearance via IL-1–mediated suppression of LPL in adipose tissues (Bartolomé et al. 2008; van Diepen et al. 2013). The other genes in this region encode inflammasome-related caspases and CARD-family regulators, some of which enhance and others suppress CASP1–dependent inflammatory signaling. The genic content of this locus supports the plausibility of its association with ’triglycerides in medium VLDL’. On the other hand, eQTLs for CASP1 and CASP4 have been identified in METSIM adipose tissue (Raulerson et al. 2019), but the lead eQTL variants are not strongly linked to the lead variant from LOCATER (r^2^<0.03) and are not associated with any traits in our study.

In addition, we detected a second potentially novel association with HDL3 cholesterol on Chr21 (Figure S14). This association was attributable to SD combining signals from a single sprig. However, in this case, only one variant is significant, and even though the lead marker is genotyped in gnomAD, it resides in a 305 bp recent segmental duplication within which variant calling is likely to be prone to artifacts, making this result less confident than others presented here.

### LOCATER boosted a classic association at the apolipoprotein gene cluster

LOCATER recovered a known association for remnant cholesterol at the well studied *APOE/C1/C4/C2* gene cluster on chromosome 19 (Richardson et al. 2022; Karjalainen et al. 2024). This same locus is associated with various other traits that are correlated with remnant cholesterol (Table S2). Although SMT also achieved genome-wide significance for remnant cholesterol, LOCATER’s advantage over SMT implies that there are additional signals from other haplotypes (Figure 5A).

**Figure 5.**
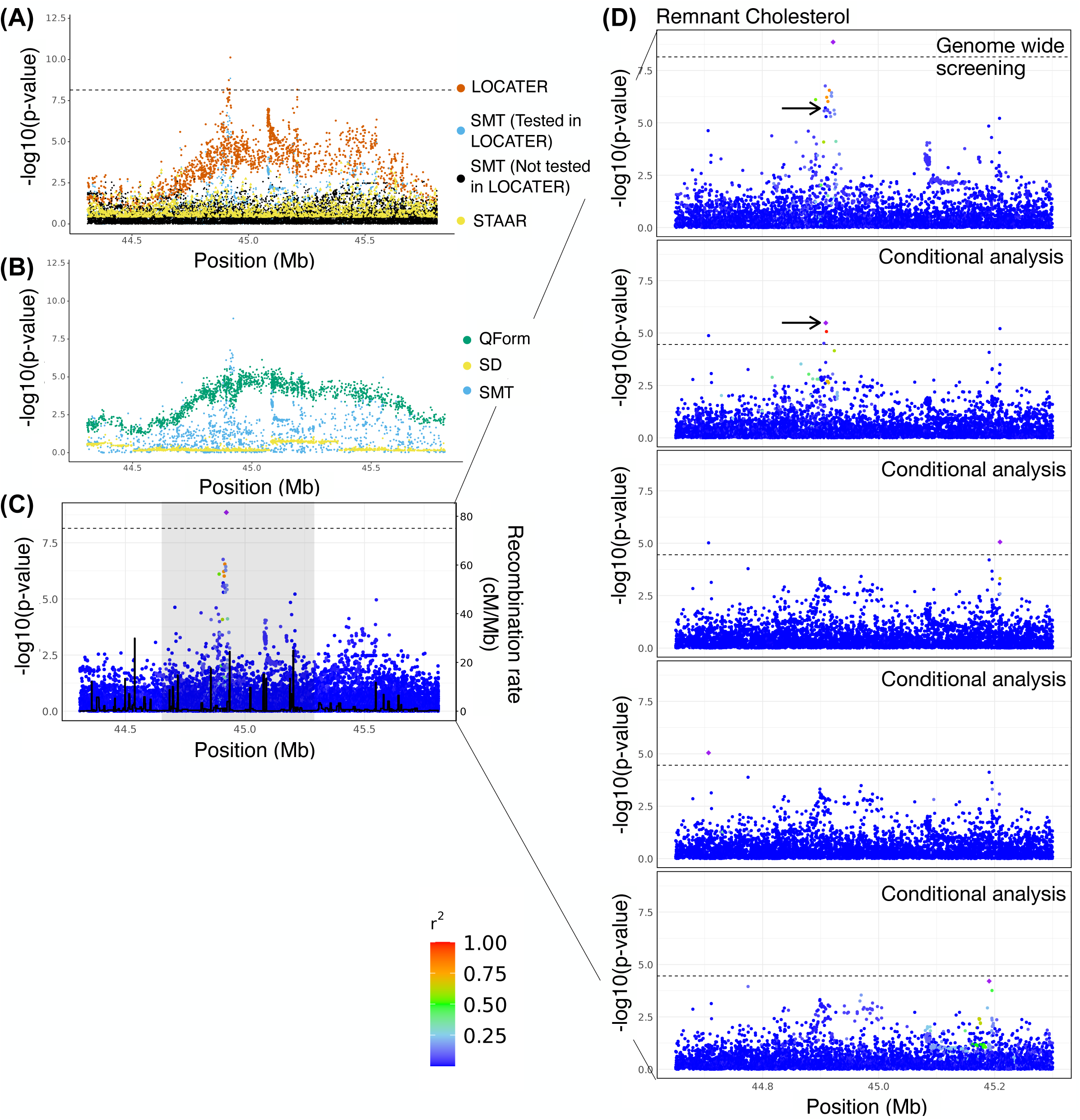
Association of remnant cholesterol at APOE cluster. **(A)** Local Manhattan plot of the association signal for remnant cholesterol on chr19:44308684-45809149, shown using the exact same data types and color scheme as Figure 2A. **(B)** Local Manhattan plot showing modified genomic controlled −log_10_(P) for the 3 LOCATER sub-tests. **(C)** LocusZoom plot of SMT results. Variants are colored based on their r^2^ with the SMT lead marker chr19:44922203 (purple diamond), where LD is calculated in the studied samples. The black line shows the recombination rate in Finns (See Methods). Gene annotations are from GENCODE v45. **(D)** Zoomed in LocusZoom plots showing the original association at top, followed by the results from stepwise conditional analysis. Results were zoomed in based on the shaded region in **(C)**. Variants are colored based on their r^2^ with the SMT lead marker of each experiment (purple diamond). Black arrows point to the most significant variant from the GWAS catalog. The black dashed line corresponds to the genome-wide significance threshold (7.17 x 10^-9^) in the top panel, and the conditional analysis threshold (3.52 x 10^-5^) for the rest.

In contrast to the other examples outlined above, LOCATER’s advantage over SMT in this case came from QForm (Figure 5B, Figure S8C), indicating that the causal haplotypes are likely to be more common (i.e., not ultra-rare). Unlike the SD subtest, QForm does not inherently provide direct insight into the number of distinct haplotypes and how they relate to each other in the genealogy; however, using multiple rounds of conditional analysis with SMT, we confirmed that there are at least four groups of causal variants. We iteratively conditioned on the genotype vector of lead markers, and observed that a significant association signal (𝑝 < 3.52 x 10^-5^) persisted through three rounds of conditional analyses (Figure 5C, D). We found that the variants used as covariates in the conditional analyses are not in LD with each other, and one of them (Chr19-44908822-C-T) is the most significant known marker associated with remnant cholesterol (Figure 5D, black arrow). These results show that LOCATER effectively combined signals from four distinct causal haplotypes, resulting in a substantial boost in power.

There are also two additional examples of known loci found by both LOCATER and SMT, where LOCATER has a more significant p-value (implying the presence of multiple causal variants): the association with ’triglycerides in small HDL’ on chromosome 20 (Figure S10) and the association of triglycerides in VLDL on chromosome 7 (Figure S12). In both of these cases the power boost was driven by the SD sub-test.

## DISCUSSION

We have used our new genealogy-based trait association method, LOCATER, to perform a genome-wide screen in a cohort of 6,795 Finnish individuals with deep cardiometabolic trait measurements and whole-genome sequencing data. In total we identified 30 associations at 13 known GWAS loci at which LOCATER was genome-wide significant and provided a clear power boost over SMT, 7 associations of which (at 5 loci) were not genome-wide significant by SMT and would have been missed. LOCATER also identified two novel association signals, one of which is fairly compelling based on the underlying haplotype structure. At each locus, dissection of the association signals and underlying haplotype structure revealed evidence for allelic heterogeneity in the form of multiple independent association signals present in distinct portions of the local ancestry tree. Moreover, in the process of optimizing LOCATER’s performance on real world genomic data, we made several key methodological improvements, including a novel approach for tuning ancestry inference parameters for trait association and a rigorous approach to account for the effects of cryptic confounders.

Genealogy based trait association has been a topic of interest for nearly two decades. Seminal early work established its potential value using theory, simulations, and single-locus analysis (Minichiello and Durbin 2006; Zöllner and Pritchard 2005), yet the practical advantages of these methods are only now becoming accessible due to recent advances in scalable tree inference (Wong et al. 2024; Kelleher et al. 2016; Zhang et al. 2023; Gunnarsson et al. 2024; Speidel et al. 2019; Deng et al. 2024), “clade association” methods capable of testing unobserved variants inferred by imputation (Zhang et al. 2023), and “global tree association” methods capable of combining association signals across multiple causal variants (Link et al. 2023; Christ et al. 2025, 2022; Zhu et al. 2024). In particular, our genome-wide analysis of 101 traits across 6,795 individuals greatly exceeds the scale and statistical power of two prior efforts (Minichiello and Durbin 2006; Link et al. 2023) that applied global tree association to human data, both of which focused on smaller cohorts, a single trait, and a subset of the genome – a single locus in the former case, and two chromosomes in the latter – and failed to detect any trait associations at genome-wide significance. More recently, Palamara and colleagues developed a new ARG inference engine and performed a gene-based association study with traits in the UK Biobank samples using a quadratic form test statistic (Zhu et al. 2024; Gunnarsson et al. 2024). This approach has impressive scalability and shows promise for improving gene-based testing; however, it remains unclear how this approach can be extended to genome-wide association testing while maintaining power, and whether it is well calibrated on real-world data with population structure. Here, we have used Stable Distillation and QForm to perform global tree association genome-wide, at virtually all non-singleton SNPs discovered by WGS, performed rigorous empirical correction for cryptic confounders, and identified hundreds of genome-wide significant trait associations. In the process, we encountered and overcame several key methodological obstacles related to parameter tuning and statistical calibration that have not previously been examined at this level of detail. These lessons are applicable to any haplotype association method, not just LOCATER, and thus this work may serve as a roadmap for future efforts.

Our work also demonstrates that genealogy based methods such as LOCATER show considerable promise for increasing the power of genetic association studies. LOCATER’s ability to combine multiple signals resulted in more significant p-values over SMT at 8.5% of genome-wide significant associations and for 10.6% of loci, and the difference in p-value exceeded an order of magnitude difference in 7 of these associations at 5 loci. The fact that such notable power gains are observed in the Finnish population – one of the least diverse human populations studied to date – suggests that our results are likely to be an underestimate of the performance improvements possible in more diverse populations that harbor more causal alleles per locus. LOCATER may be especially valuable for multi-ethnic studies, where allelic heterogeneity is greatest, and where standard association methods have shown poor performance.

Relatively few of the 30 association signals for which LOCATER provided a power boost are likely to have been captured by other methods. None of these signals were captured by STAAR using a window-based screening approach that has been employed previously for rare variant association studies (although we note that STAAR is not explicitly designed nor typically used for non-coding studies). Moreover, although two recently developed methods (Link et al. 2023; Gunnarsson et al. 2024; Zhu et al. 2024) have some similarities with LOCATER, they rely on a quadratic form test for global tree association, whereas LOCATER uses Stable Distillation (SD) in addition to QForm. We have previously shown via simulation-based studies that SD and Qform have complementary strengths and weaknesses: QForm excels when the causal variants are more common and reside further up in the tree, and SD excels at combining ultra-rare variants residing in small clades at the bottom of the tree (Christ et al. 2025; Wang 2024). LOCATER tests SD and QForm sequentially, such that QForm is at a disadvantage for detecting signals. Nonetheless, given the complementary nature of the two tests, it is interesting to note that for the 21 non-redundant trait associations (accounting for correlated traits) where LOCATER obtained more significant results than SMT alone, 15 were boosted by SD and 6 by QForm. This, along with our in-depth analysis of ten loci, suggests that the SD test unique to LOCATER is providing substantial value beyond QForm by efficiently combining association signals across many clades. We expect the relative value of SD to increase as allelic heterogeneity increases, as in more diverse populations or for traits under negative selection. Despite these promising results, global tree association remains a difficult statistical problem, and we expect that future work in this area will continue to yield significant power improvements.

Tree-based trait association also presents unique challenges relative to SMT and gene-based testing. Perhaps the most difficult aspect of this study was the detailed work required to optimize haplotype inference and to control for cryptic confounders, both of which are important practical considerations for future studies. First, it is well known that the performance of the LS model HMMs is sensitive to the mutation and recombination penalty parameters, and prior studies have used different approaches for selecting them. We developed a novel tuning approach that optimizes regional trait association power at a distance of 0.05 cM rather than local variant imputation as in Relate (Speidel et al. 2019), and we explored a broad range of potential parameters to optimize haplotype inference for this specific population and dataset. Optimizing for trait association power rather than imputation ensures that the resulting haplotype representations used carry long-range information required to combine independent signals at loci with allelic heterogeneity.

Second, we found that genealogy-based methods such as LOCATER are extremely sensitive to cryptic confounders, much more so than standard SMT, and that special measures are required to control for these effects. We studied this issue in detail and devised two new approaches to control for type I error: permuted rank matching of phenotype data to simulated Gaussian random variables to produce independent Gaussian phenotypes with minimal confounding, and a more general type of genomic control that fits both a slope and an intercept. In combination, these measures led to a well calibrated analysis in our study and are likely to be applicable to future genealogy based screens as well. Interestingly, the confounding effect of cryptic confounders was only apparent in our LOCATER analysis, whereas SMT appeared to be well calibrated after standard PCA-based measures. This result suggests that unadjusted confounders may plague SMT results to a larger extent than predicted from the body of QQ plots alone. We hypothesize that LOCATER is more sensitive to confounders than SMT because of the high correlation between proximal inferred genealogies along the genome.

For intuition, consider the simple case where there is a small sub-population within a dataset that has some environmental exposure that affects the phenotype of interest. There will be variants that tag that sub-population throughout the genome. Every local ancestral tree inferred near those confounded variants will have a clade marking that sub-population and thus have inflated test statistics. Thus, any potential confounders will affect LOCATER at many more markers than SMT, making the resulting inflation obvious in Q-Q plots.

Although there is more work to be done before these methods are mature, the work presented here – in combination with our prior simulation based results (Christ et al. 2025) and recent work from others (Link et al. 2023; Gunnarsson et al. 2024; Zhu et al. 2024) – suggests that genealogy-based trait association methods such as LOCATER are finally ready to fulfill their long-promised potential as practical tools for genome-wide association studies.

## METHODS

### The METSIM study

METSIM is a single-site study investigating cardiometabolic disorders and related traits in 10,197 men randomly selected from the population register of Kuopio, Eastern Finland, aged 45 to 73 years at initial examination from 2005 to 2010 (Laakso et al. 2017). All participants provided informed consent. The phenotype data used for the work described here were adapted from Locke et al. (Locke et al. 2019), in which the authors accounted for different factors (including trait specific background covariates) during linear regression of the raw phenotypes, and used rank-based inverse normal transformation.

### Whole-genome sequencing, data processing, and variant calling

DNA samples were extracted from blood. We constructed DNA libraries with automated Kapa Hyper PCR free, automated TruSeq PCR free, Kapa Hyper PCR free, or TruSeq PCR free kit, with target insert size varying from 260 to 475. The libraries were sequenced with Illumina HiSeq 2000, 2500, X10, or NovaSeq 6000, generating 2 x 151-bp paired-end sequencing data.

We performed alignment and data processing using the “functional equivalence” pipeline (Regier et al. 2018). Briefly, we aligned reads to the GRCh38 human reference genome using bwa-mem (v.0.7.15) and used Picard MarkDuplicates (v.2.4.1; http://broadinstitute.github.io/picard) to remove duplicate reads. We excluded samples that had estimated contamination >5% or that were likely to represent sample swaps (verifyBamID v.1.1.3). We also required a discordant rate of <5%, haploid coverage ≥19.5X, inter-chromosomal rate of <5%, and first-of-pair mismatch rate of <5%.

We performed variant calling with HaplotypeCaller in GATK (v.3.5) and concatenated the output into full single sample GVCFs (Picard MergeVcfs, v2.4.1). Since standard joint genotyping with GATK GenotypeGVCFs function could not scale to our CCDG callset, we used ReblockGVCFs (GATK v.4.2.2.0) to decrease GVCF file sizes for future joint analysis in Hail. We used the ValidateVariants function to ensure the quality of the reblocking process. We then used the VariantDatasetCombiner function in Hail (v.0.2.78) to combine GVCFs from each sample into multi-sample VariantDataset (VDS) files before running GnarlyGenotyper (unpublished version from Docker image gcr.io/broad-dsde-methods/gnarly_genotyper:hail_ukbb_300K, image hash ID: 7cc8cfa6e9af; created April 2020; received August 2021) for joint genotyping and VQSR to annotate variant quality. Finally, we converted VDS files into MatrixTable (MT) files (Hail v.0.2.97) and decomposed multi-allelic variants into bi-allelic variants.

To QC the variant callset, we excluded samples that had a low het/hom ratio (<5 MADs less than the median), low sequencing depth (number of bases with depth >10 is <20 MADs less than the median), or excessive number of singleton variants (>20 MADs more than the median), where each of these criteria was applied separately to each self-reported ancestry group. We also removed samples with fewer than 580,000 insertions or deletions in joint variant calling, genetic-phenotypic sex mismatches, withdrawal of consent, sample swaps, or inheritance inconsistencies and other sample identity issues. The maximal independent set of these samples was calculated in Hail using ‘hl.maximal_independent_set’ and individuals up to second-degree related were removed. For variant level QC, we flagged genotype calls with genotype quality (GQ) < 20, depth (DP) < 10, and heterozygous calls with allele balance (AB) ≤ 0.2 or ≥ 0.8 as low quality, and filtered out variants that had AS_VQSLOD < 0 or that had a high proportion (>95%) of missing or low quality genotypes.

### Phasing and variant annotation

We performed a more stringent variant quality control for phasing. After exporting Hail MT files to VCFs (Hail 0.2.95), we selected PASS and non-singleton variants and filtered out sites with high quality genotype call rate < 90% or Hardy-Weinberg 𝑝 < 10^-7^ (one-sided p-value for excess heterozygotes).

METSIM samples were phased with other WashU CCDG samples without a reference panel with Eagle2 (v.2.4.1). Due to memory restraints, we divided chromosomes into 20 Mb chunks with 2 Mb overlaps on both ends. Phasing was done with default options except for a bigger ‘Kpbwt’ value (200k) for better phasing accuracy. Missing genotypes were imputed during phasing.

### Ancestral allele encoding

The Speidel version of the LS model (Speidel et al. 2019) assumes the ancestral allele is known for all variants and uses that information in its ancestry inference. For this study, we used ancestral allele calls obtained via a 10-way EPO alignment of primates from Ensembl v106 (The 1000 Genomes Project Consortium et al. 2015). With ‘bcftools’, we updated the REF and ALT alleles and the genotype fields in VCF files to make the REF allele the ancestral allele. Encoding VCFs with the ancestral allele in large-scale data requires ancestral allele calls in FASTA format. Explicitly, we downloaded the human ancestral genome FASTA from 10-way EPO alignment of primates from Ensembl v106 available at https://ftp.ensembl.org/pub/release-106/fasta/ancestral_alleles/homo_sapiens_ancestor_GRCh38.tar.gz created on March 19, 2022. In the FASTA file, lowercase indicates lower quality. For simplicity, all lowercase letters were converted to uppercase. Since the conversion works best when multiallelic variants are merged, we merged them with ‘bcftools norm -m +any’ (bcftools v1.9). We then used ‘bcftools norm --check-ref s --fasta-ref {fasta_file} ‘ (bcftools v1.9) to edit the REF allele in VCF files to be the ancestral allele, which automatically updated the genotype field. The files also went through ‘bcftools +fill-tags’ (bcftools v1.16) to make sure the AC tag of the INFO column in VCF files is correct. Finally, we split multiallelic sites into biallelic variants with ‘bcftools norm -m -any’ (bcftools v1.16).

### The LOCATER pipeline

We performed ancestry-based association testing using the LOCATER pipeline, which is described in detail elsewhere, including methodological details and an in-depth evaluation of simulated data (Christ et al. 2025). The detailed description of the real data pipeline of this study is described here: https://github.com/Xinxin-Wang-0128/LOCATER_real_data_vignette. Briefly, the first step is to run local ancestry inference at each genetic marker included in the study. For this we used the newest version (v2) of our local ancestry inference engine, kalis (Aslett and Christ 2024), which is an optimized implementation of the Speidel version of the LS model (Speidel et al. 2019). Notably, unlike other LS model implementations, kalis v2 uses an optimal checkpointing algorithm which allows it to be run on arbitrarily large sequences (e.g., whole chromosomes). Here, in the interests of computational efficiency, we only performed local ancestry inference on genomic segments that contained one or more single markers with a promising p-value (P < 10^-3^ in this study). In total, this included 5.7% of the genome. This step produces an N x N matrix (where N is the number of haplotypes; 13,590 in this case) of genomic distances at each genetic marker.

The second step is to identify small clades, which we refer to as “sprigs”. LOCATER includes a sprig calling algorithm that uses a multithreaded partial sorting algorithm to cluster the genomic distances into level sets separately for each haplotype, followed by a greedy clique finding procedure to rapidly call sprigs.

Then, at each genetic marker, LOCATER performs three types of association tests: (1) a standard SMT to measure the contribution of that specific marker; (2) Stable Distillation (SD) to measure the contribution of the inferred small clades; and (3) a quadratic-form based test to measure the contribution of remaining ancestral relationships present at deeper portions of the tree (Christ et al. 2025). Steps 2 and 3 use a residualized phenotype vector from the prior step. This, combined with the independence guarantees of SD, ensures that the resulting three p-values at a given site are mutually independent under the null hypothesis. We then combine the three p-values using an adapted version of Fisher’s method that we call Maximizing over Subsets of Summed Exponentials (MSSE) (Christ et al. 2025).

### Rank matching and selection

The SD procedure used in LOCATER requires that all phenotypes have a Gaussian distribution. As we show in Figures S2, S3 and S4, since the quadratic form testing procedure is downstream of SD, violating this assumption can affect the null distribution of both SD and QForm. We ensured the Gaussianity of our phenotype vector by rank-matching our phenotypes to a vector of simulated independent Gaussian random variables.

During preliminary analyses, we repeated this rank-matching process for the same phenotype several times, yielding a set of vectors, each a subtly different perturbation of the original phenotype. When we ran LOCATER across these different perturbations, the amount of inflation observed via the Q-Q plot of genome-wide LOCATER p-values differed moderately across perturbations (see examples in Figures S2, S3, and S4). We believe this variation reflects the fact that, by chance, different perturbations can have stronger or weaker correlations with confounding processes such as population structure. Note that genome-wide SMT p-values are consistently well-calibrated for all perturbations; thus, no adjustments were needed for SMT. Based on this observation, we simulated 100 perturbations of each phenotype and selected the perturbation that minimized the deviation from the expected tail distribution under the null hypothesis. Explicitly, for all perturbations of each phenotype, we ran LOCATER across evenly spaced variants (∼30,000 variants) along the genome, and after plotting the Q-Q inflation plot for -log_10_𝑝 of SD and QForm in LOCATER, we fitted a least-squares line over the 𝑥 ∈ [2,2.5] domain of the resulting Q-Q inflation plot. This corresponds to fitting the tail of the null distribution based on all p-values in [0.01,0.0032]. We then used the parameters of each least square line (the slope and intercept) to select the perturbation that was closest to the expected null distribution. More explicitly, let the slopes of SD and QForm Q-Q plots be 𝑚*_SD_* and 𝑚,*_Q_* while intercepts are 𝑏*_SD_* and 𝑏*_Q_*, respectively. First, we selected all perturbations satisfying the following boolean expression 𝑚*_SD_* ∈ [0.8, 1.1] AND 𝑏*_SD_* ∈ [-0.1, 0.1] AND (( 𝑚,*_Q_*∈ [0.7, 1.2] AND 𝑏*_Q_* ∈ [-0.1, 0.1] ) OR (𝑚,*_Q_*∈ [0.6, 0.8] AND 𝑏*_Q_* ∈ [0, 0.4])). If multiple perturbations met this requirement, the perturbation with the largest 𝑚𝑖𝑛(𝑚*_SD_*, 𝑚*_Q_*) was chosen. If no perturbation met this requirement, which happened for 15 of our 101 phenotypes, the standard rank-based inverse normalized phenotype was used. After the screening, we adjusted the p-value for each sub-test based on the estimated tail parameters, 𝑚 and 𝑏, via

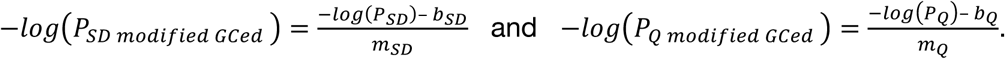

Since SMT is consistently well-calibrated, no adjustments were applied to SMT p-values. To combine the resulting SMT, SD, and QForm p-values returned by LOCATER, we used the MSSE method in Christ et al. (Christ et al. 2025).

### Tuning ancestry inference for trait association

The LS model at the heart of the ancestry inference we used in this study, kalis, is a hidden Markov model (HMM) with two parameters that can be interpreted as tolerance for recombination and mutation, respectively. As we will delineate below, rather than using expectation-maximization or other more standard tuning objectives to select our recombination and mutation parameters, we chose parameters to optimize the propagation of proximal association signals along the genome in order to maximize LOCATER’s power.

In our tuning procedure, we randomly sampled core genomic regions with at least 15,000 variants, each with flanking regions of 5,000 variants on both sides. These flanking regions served as “burn-in” regions to ensure accurate ancestry inference along the full length of the core region. We then selected a variant in the middle of the core region as our target variant and used kalis to perform ancestry inference at that site.

We then selected a causal variant 0.05 (± 0.005) cM away. In LOCATER, as in all GWAS studies, any association signals driven by variants that are colinear with the background covariates are assumed to be attributable to confounding processes. Thus, we only chose causal variants that were not colinear with the background covariates (multiple r^2^ ≤ 0.02). We then simulated a pseudo-phenotype vector with a strong effect driven by the causal variant. In order to ensure that the strength of this causal variant’s signal (in terms of the observed -log_10_ p-value) would be roughly consistent across simulations, we increased the strength of the causal variant’s effect as a function of the number of clades inferred at the target locus: more null clades yield a higher multiple testing burden for the causal variant to overcome.

Given our pseudo-phenotype vector, we ran LOCATER at our target variant *l*, yielding two p-values 𝑝*_SD_^l^* and 𝑝*_Q_^l^*. We assessed how these two p-values captured the signal at the nearby causal variant using

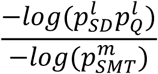

as our relative signal metric. Here 𝑝^=^ is the SMT p-value at the causal variant.

We calculated this relative signal metric for 14 parameter settings (see Table S3) across 144 distinct core regions sampled from our METSIM dataset. Altogether, this procedure allowed us to select LS HMM parameters to maximize LOCATER’s power in this METSIM study.

### Estimating the effective number of independent tests

Our dataset has a huge number of rare variants, and the canonical 5 x 10^-8^ threshold for p-value was based on 1 million independent tests. This effective number of tests was identified in the array genotyping era, and a couple of studies (Karczewski et al. 2022; Ishigaki et al. 2020) argued that this was an overly generous threshold. There are papers describing methods to calculate the effective number of independent tests (Gao et al. 2008), and they agreed that the gold standard is the permutation-based method (Hoh et al. 2001). Permutation-based methods permute the phenotype N times and do the association analysis for those permuted phenotypes. Then the minimum p-value (𝑝_=:7_) was recorded for all these permuted phenotypes. If we want to control the type I error rate to be 0.05, the new threshold should be the 95th percentile of the 𝑝_=:7_ distribution. Here we used N = 1000 permutations and targeted a type I error rate of 0.05 (Gao et al. 2008).

We simulated 1000 normally distributed quantitative traits, ran SMT first, and used the subset of variants with 𝑝_()*_ < 10^-4.5^ for LOCATER association. Based on the 𝑝_=:7_ distribution, the experiment-wise significance threshold should be 7.17 x 10^-9^ for 𝑝_()*_ and 1.41 x 10^-8^ for 𝑝_-./0*12_ if the type I error rate is 0.05. To directly compare LOCATER and SMT, the final p-values of LOCATER were standardized so that its effective number of tests matches that of SMT and both tests use the same threshold 7.17 x 10^-9^.

### Association Screening

Since the Speidel version of the LS model requires ancestral allele calls (Speidel et al. 2019), we only included SNPs with ancestral allele calls in the 10-way EPO alignment of primates from Ensembl v106. Indels were ignored due to their lower quality of phasing and variant calling. After removing monomorphic sites and singletons, we obtained a final dataset of 18.9 million variants.

Kinship among individuals was calculated and only unrelated individuals were included. For kinship analysis, we used the Hail function ‘hl.pc_relatè to compute pairwise kinship coefficients, and individual pairs with kinship ≥ 0.05 were flagged as related outliers. Principal components (PCs) were calculated in all Finnish samples, and a sample is considered a PC outlier and later removed if its score for any of the top 10 PCs falls outside the range of ±8 standard deviations (SD) from the mean score of that PC. This yielded a final dataset of 101 traits and 6,795 individuals. Background covariates in this study were the top 10 principal components.

For computational efficiency, we adopted the following three strategies. First, we divided the whole genome into 4,587 segments, with ± ∼6,000 variants of overlap between segments. The 4,587 segment boundaries were defined so that each segment has a relatively similar number of variants, to ensure similar run times during analysis. Second, while all of our variants were used for ancestry inference by kalis, we only ran LOCATER on a relatively small subset of target variants. We skipped variants where the SMT p-values from all phenotypes were greater than 10^-3^ while ensuring that the recombination distance between consecutive target variants was at most 0.1 cM. Third, we avoided any expensive eigendecomposition steps in calculating 𝑝*_Q_* at our target loci by using the Satterthwaite approximation to 𝑝*_Q_* (Christ et al. 2025) in our first round of screening.

We identified any variant with a LOCATER combined p-value smaller than 7.17 x 10^-8^ in our first round of screening as putative variants. We then merged putative variants in all the phenotypes to generate putative loci (see main text) for follow-up. During association follow-up, we only focused on 10 cM regions centered around putative loci, which doubly ensured reliable ancestry inference. During LOCATER testing, we used partial eigendecomposition and Chi-square-based approximation (details in Equation 11 in the Methods of (Christ et al. 2025) on the local relatedness matrix to obtain precise 𝑝_Q_

Because of the stochastic nature of the SD procedure, the p-values 𝑝*_SD_* and 𝑝*_Q_* returned by LOCATER are a function of the seed of the R environment. Setting the same seed for all segments along the genome is not recommended, since it will cause additional correlation between variants, which will cause the p-value distribution to deviate from the expected uniform distribution. Instead, we strongly suggest setting different seeds for different segments along the genome. However, for reproducibility or for follow up experiments of the same region or segment, we suggest using the same seed across experiments for consistent results.

To assess the novelty of the association results we used the GWAS catalog (April 22nd, 2024 version; https://www.ebi.ac.uk/gwas/home)

### Rare variant association experiment with STAAR

We applied STAAR with the sliding window method using default parameters. The window size is 2 kb and the sliding step length is 1 kb. For each window, we included all rare variants with AF < 0.01 and removed any window with fewer than two rare variants. We reported the STAAR-O p-value.

### Hierarchical clustering and visualization of local distance matrices

At loci with significant LOCATER associations driven by SD, we constructed a local tree based on the local relatedness matrix obtained by kalis in order to understand the relative placement of the sprigs driving that SD signal. Following the method used in Speidel et al. (Speidel et al. 2019), we did this using mean-based hierarchical clustering (UPGMA) implemented in the ‘fastcluster’ R package (Müllner 2013). For plotting clarity, 95% of haplotypes under insignificant sprigs were pruned from the displayed dendrograms.

### Residual analysis to visualize SD signals

We employed a residual analysis procedure to visualize the association signals that contributed to the SD signal at a locus. Recall that LOCATER is a three-stage procedure where a phenotype vector is passed between each step. In order to isolate the signals extracted by SD, we made a local Manhattan plot regressing the phenotype vector passed to SD (above we refer to the resulting p-value at a given variant as 𝑝_(_) and overlaid it with a Manhattan plot regressing the phenotype vector returned by SD (above we refer to the resulting p-value at a given variant as 𝑝*_D_*). We highlighted variants where 𝑝_S_ <1 x 10^-3^ and 𝑝*_D_* > 10 * 𝑝*_D_*.

## DATA ACCESS

The code used to perform the analyses in this paper is available on a GitHub page: https://github.com/Xinxin-Wang-0128/LOCATER_real_data_vignette. The link to the LOCATER software is: https://ryanchrist.github.io/locater/. METSIM data are available from dbGaP (accession number for genotype data: phs001579; phenotype data: phs000752).

## Data Availability

There is no new data generated.

## ACKNOWLEDGEMENTS

RC and XW were supported by NIH grants R01HG013371 and UM1HG008853 to IH. LJMA was partially supported by the EPSRC research grant “PINCODE”, reference EP/X028100/1, and UKRI grant, “OCEAN”, reference EP/Y014650/1. DS was supported by BBSRC research grant BB/S001824/1. For the purpose of Open Access, the authors have applied a CC BY public copyright license to any Author Accepted Manuscript version arising from this submission.

## Author Contributions

XW, RC and IMH conceived and designed the analysis. IMH and NOS designed the sequencing study. ML created the METSIM cohort. EY, CJK, ID, EABJ generated the variant callset, EY curated the phenotype data, XW, RC, LJMA and DS contributed analysis tools, and XW performed the analysis. XW, RC and IMH wrote the paper with input from NOS, DS and LJMA.

