## Supplemental Figures for "Genealogy based trait association with LOCATER boosts power at loci with allelic heterogeneity"

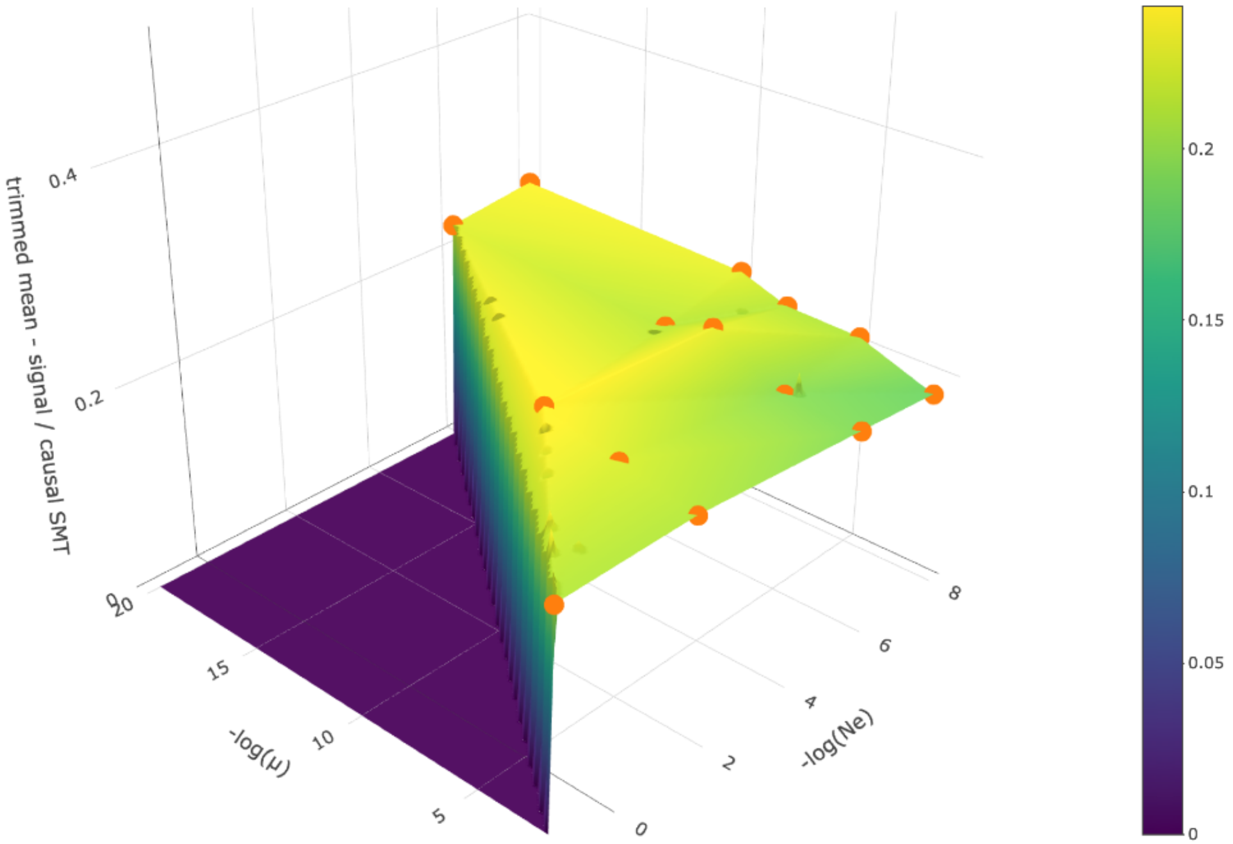


**Figure S1**: The LOCATER tuning surface averaged across all allele frequencies. Orange points show the parameters evaluated. The X-axis corresponds to the recombination penalty, -log(Ne); the Y-axis is the mutational penalty, -log(μ); the Z-axis is the trimmed mean of relative efficiency, obtained by removing the largest and smallest 10% of values before calculating the mean. Relative efficiency is defined as $-log(p^{l}_{SD}p^{l}_{Q})/(-log(p^{l'}_{SMT}))$, where $l$ is the target variant and $l'$ is the causal variant. The phenotype vector was simulated based on a single causal variant ($l'$) with a strong effect.

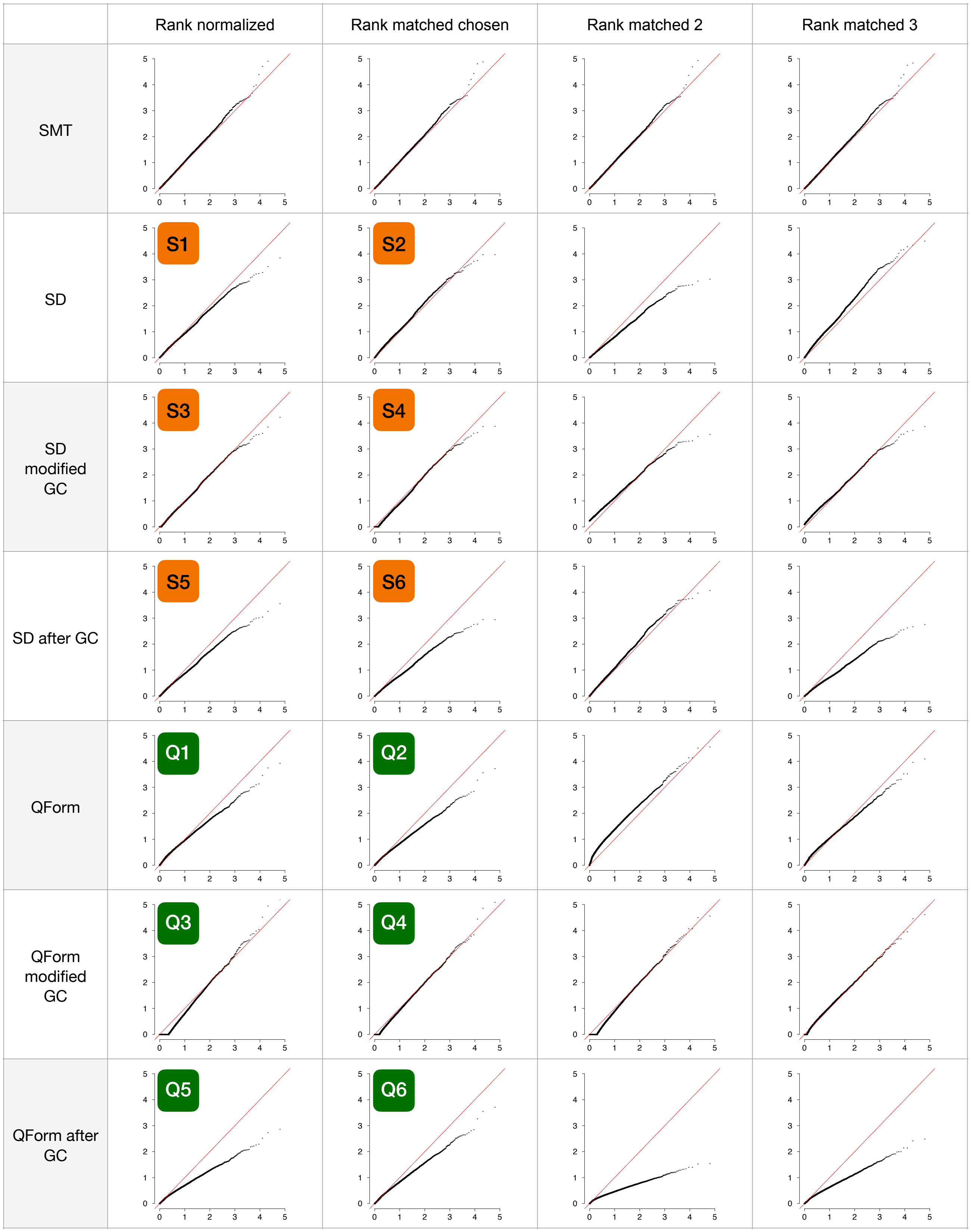
Rank normalized
Rank matched chosen
Rank matched 2
Rank matched 3
SMT
SD
SD modified GC
SD after GC
QForm
QForm modified GC
QForm after GC
S1
S2
S3
S4
S5
S6
Q1
Q2
Q3
Q4
Q5
Q6

**Figure S2**: Q-Q plots for “triglycerides in medium HDL”. The column “Rank normalized” shows results for the original rank normalized phenotype vector; “Rank matched chosen” shows the rank matched phenotype vector used for the association study; “Rank matched 2 and 3" shows results of two additional rank matched phenotype vectors, selected as examples from the 100 versions to contrast with the best version shown in column 2. From the top, the rows show results for single marker tests (SMT), stable distillation (SD), SD after modified genomic control (GC), SD after traditional GC, QForm, QForm after modified GC and QForm after traditional GC. To adjust for confounding, we applied a more general genomic control that achieves a more accurate fit to the tail of the null distribution of $-\log_{10}$(p-values). See details in Methods and Results. GC: genomic control. All data points where both the x and y values exceed 5 are ignored.

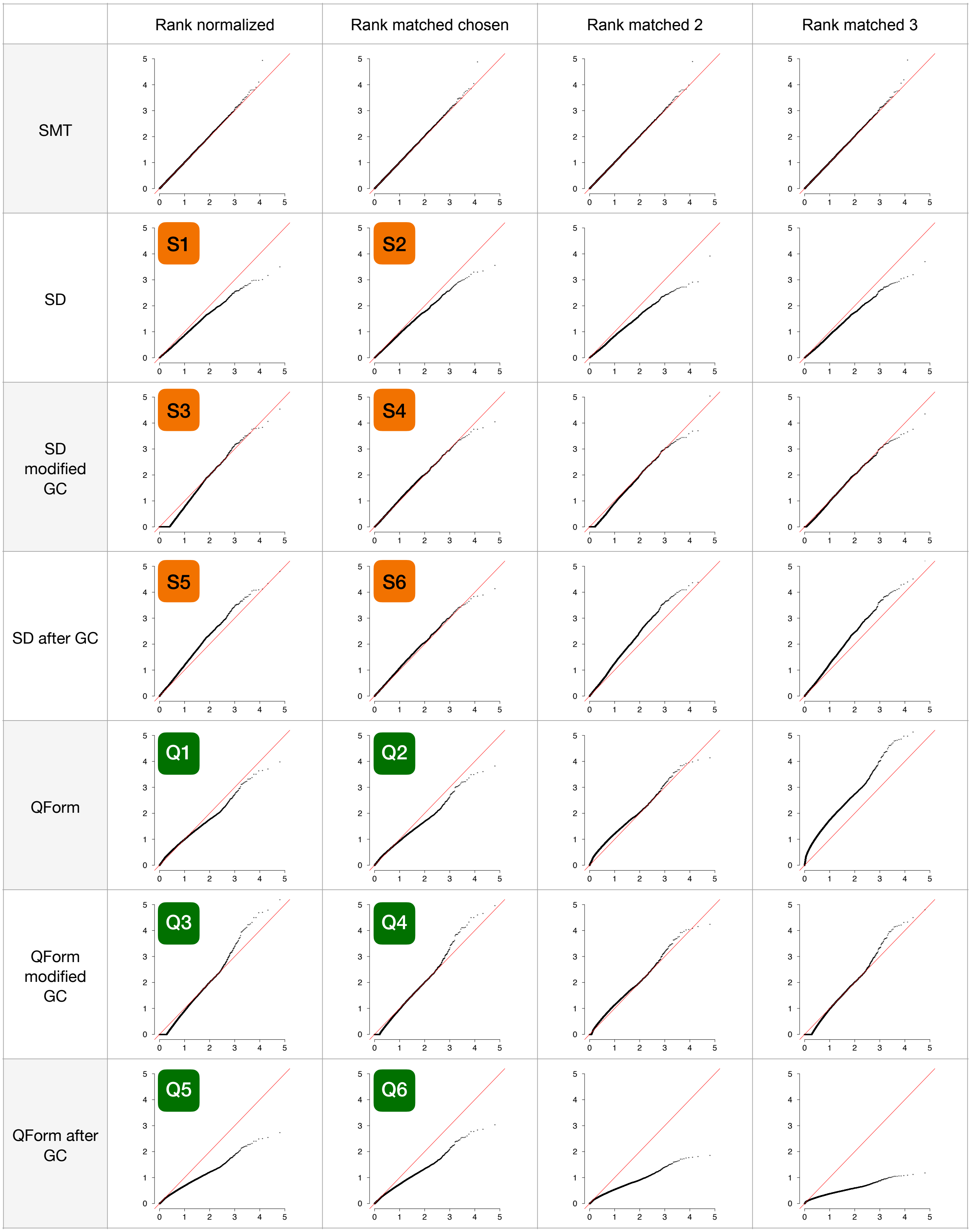
Rank normalized
Rank matched chosen
Rank matched 2
Rank matched 3
SMT
SD
S1
S2
SD modified GC
S3
S4
SD after GC
S5
S6
QForm
Q1
Q2
QForm modified GC
Q3
Q4
QForm after GC
Q5
Q6

**Figure S3**: Q-Q plots for Apolipoprotein A1, with the same columns and rows as in Figure S2. GC: genomic control. All data points where both the x and y values exceed 5 are ignored.

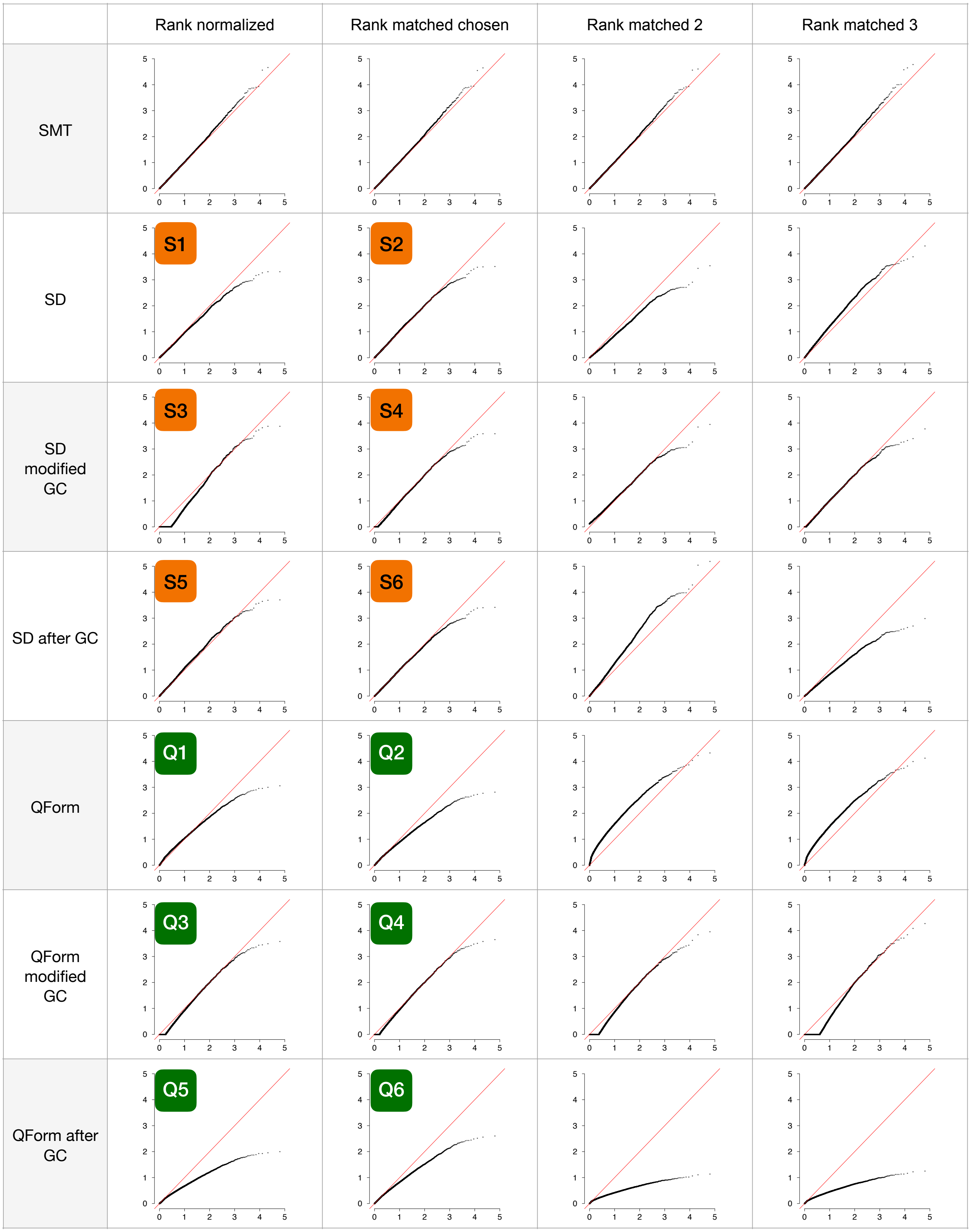
Rank normalized
Rank matched chosen
Rank matched 2
Rank matched 3
SMT
SD
S1
S2
SD modified GC
S3
S4
SD after GC
S5
S6
QForm
Q1
Q2
QForm modified GC
Q3
Q4
QForm after GC
Q5
Q6

**Figure S4**: Q-Q plots for “Triglycerides in medium VLDL”, with the same columns and rows as in Figure S2. GC: genomic control. All data points where both the x and y values exceed 5 are ignored.

Figure S5: triglycerides in medium HDL at *LIPG*
Figure S5A-B

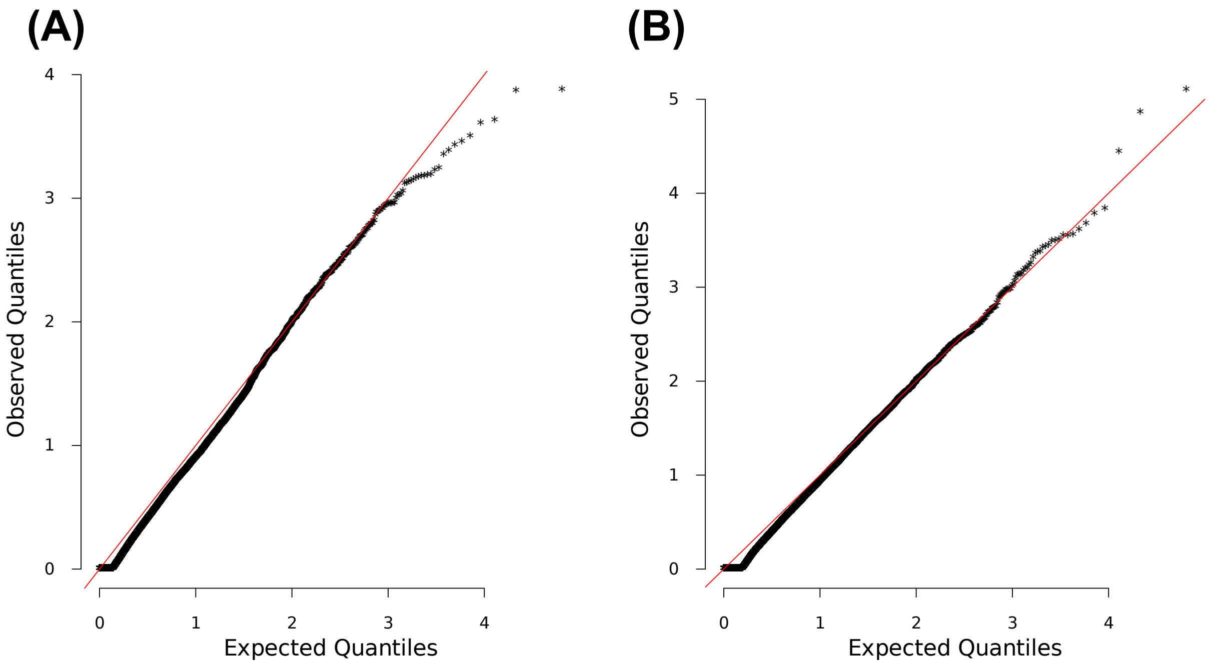


**(A)** and **(B)** are Q-Q inflation plots after modified GC of the LOCATER sub-tests stable distillation (SD) and quadratic form (QForm), respectively, for “triglycerides in medium HDL”. The red line on the diagonal corresponds to x=y.

Figure S5C-D

**(C)**

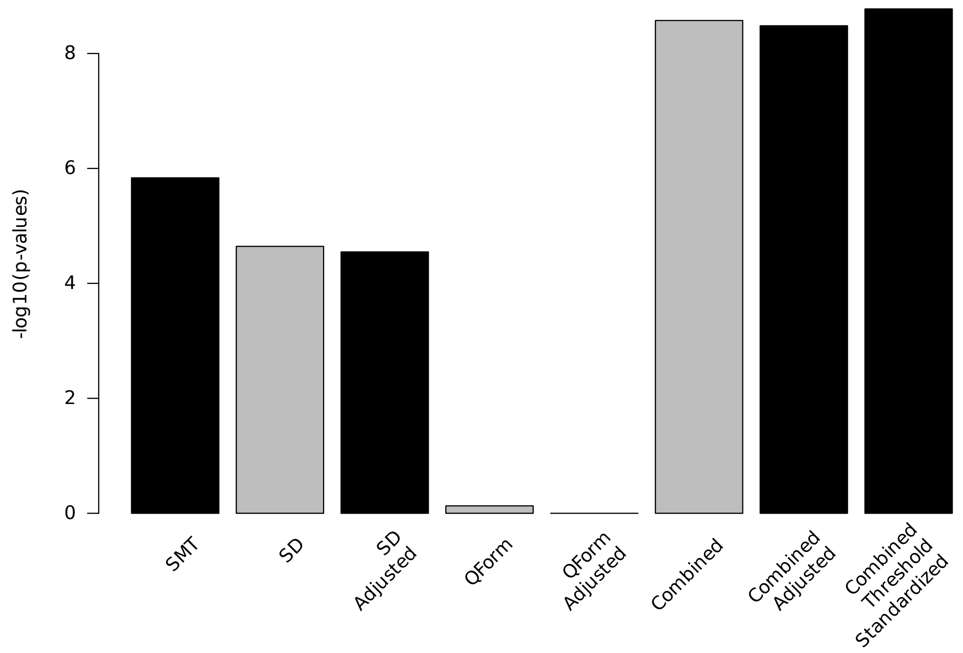


**(D)**

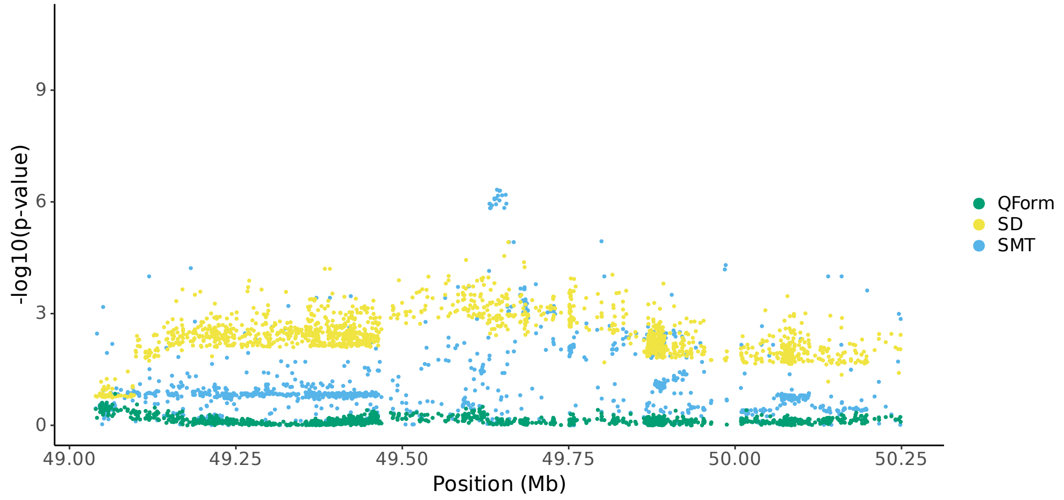


**(C)** Bar plot of -$\log_{10}$(P) for association results before and after modified GC at lead marker chr18:49653146. Shown are the three individual sub-tests, including single marker test (SMT), stable distillation (SD) and quadratic form (Qform), as well as the three tests combined. Grey bars show -$\log_{10}$(P) for SD, QForm and "combined" before modified GC by the slope and intercept of Q-Q plots, black bars show results after modified GC, and the final black bar at right shows the final combined -$\log_{10}$(P) used for all final results, which also accounts for the different number of independent tests performed by SMT and LOCATER. **(D)** Local Manhattan plot of triglycerides in medium HDL on chr18:49038347-50253146, showing modified GCed -$\log_{10}$(P) for the 3 LOCATER sub-tests.

Figure S5E-F

**(E)**

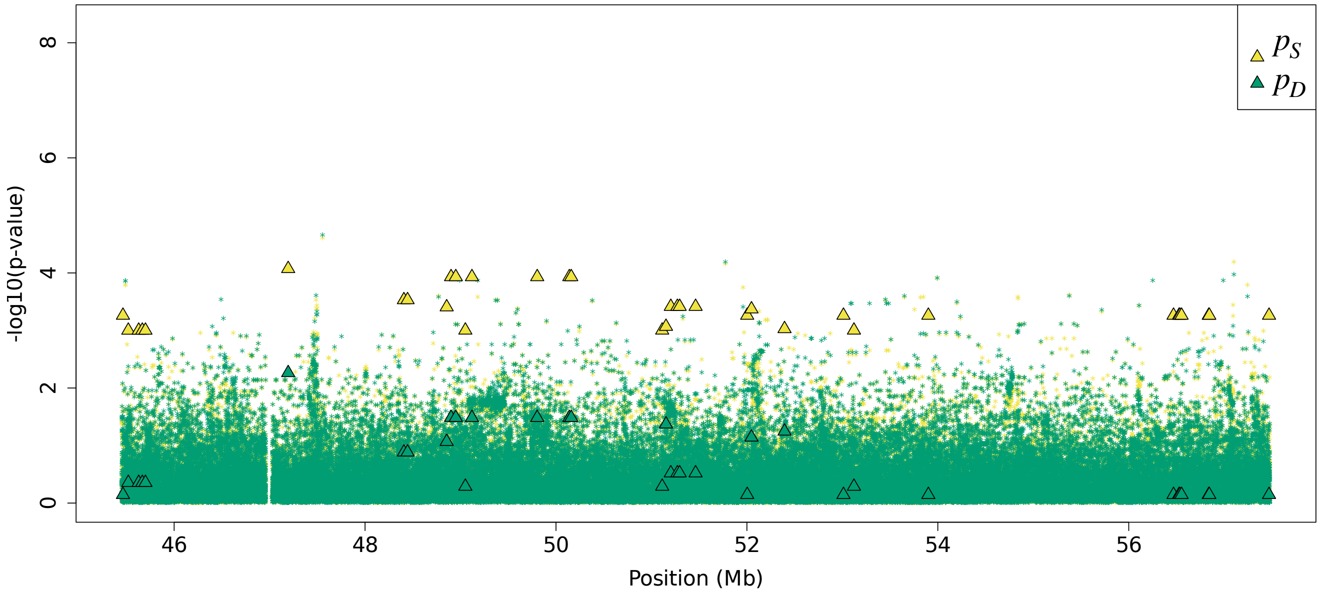


**(F)**

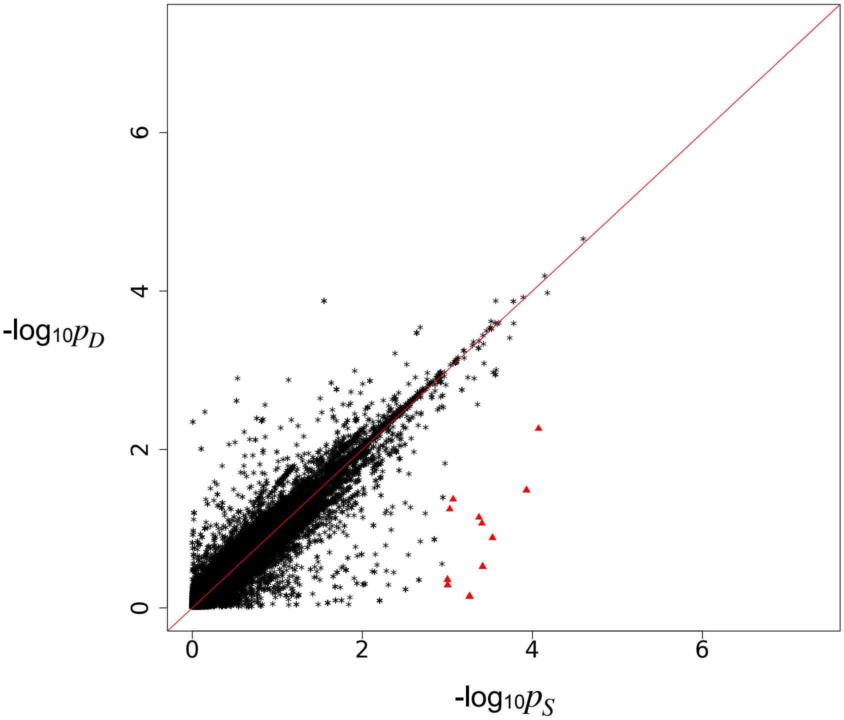


Residual association signals at the *LIPG* locus after accounting for signal from the LOCATER lead marker. $p_S$ is defined as the SMT p-value based on a residualized phenotype orthogonal against the LOCATER lead marker; $p_D$ is the SMT p-value based on a residualized phenotype orthogonal against the LOCATER lead marker and also the SD signal. The difference between $p_S$ and $p_D$ thus shows the contribution of genomic variants to the SD signal. Triangles: genomic variants with $p_S < 10^{-3}$ and $p_D > 10 * p_S$. **(E)** Local Manhattan plot of $p_S$ and $p_D$. **(F)** Scatter plot of $p_S$ and $p_D$.

Figure S5G

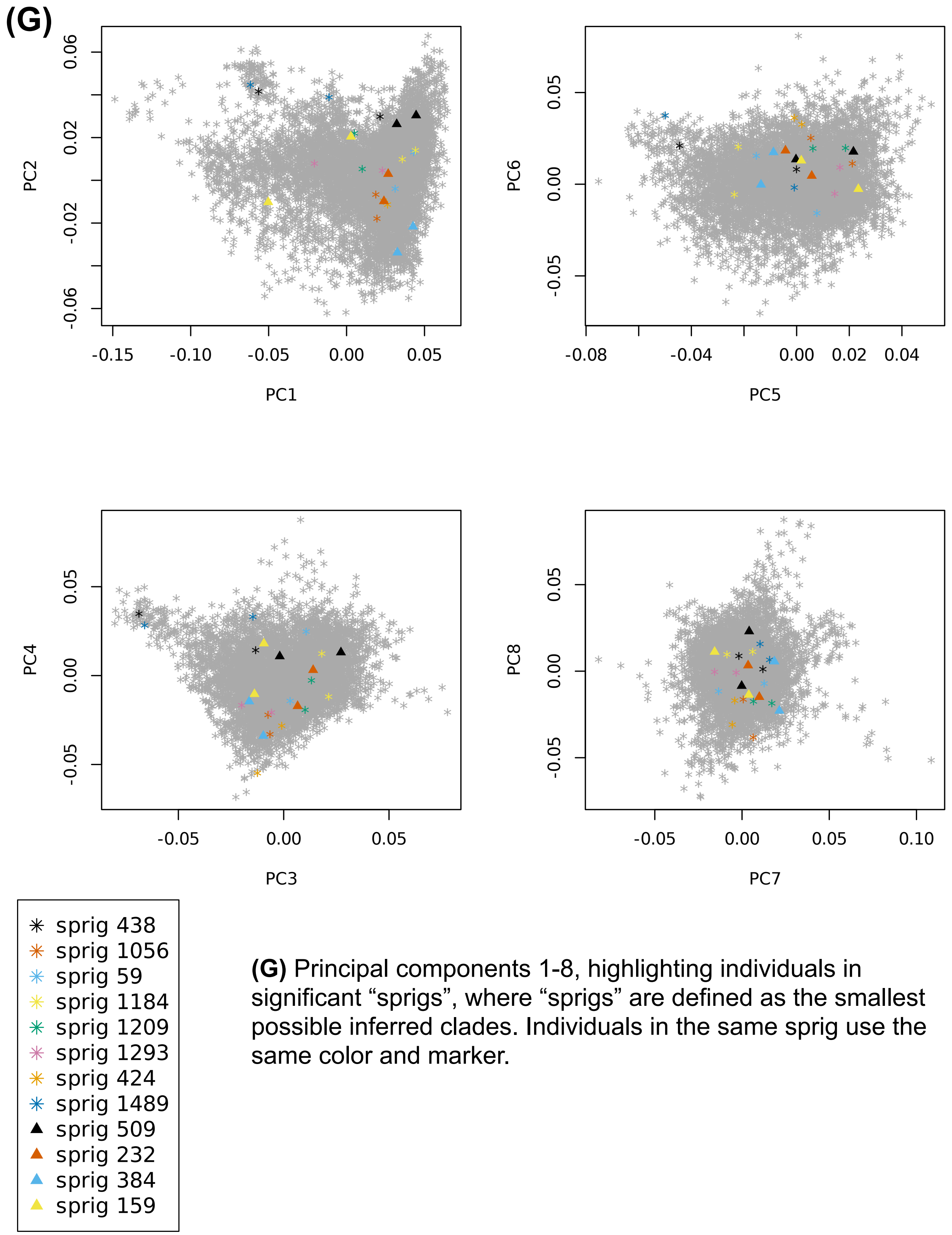


**(G)** Principal components 1-8, highlighting individuals in significant “sprigs”, where “sprigs” are defined as the smallest possible inferred clades. Individuals in the same sprig use the same color and marker.

### Figure S5H-I

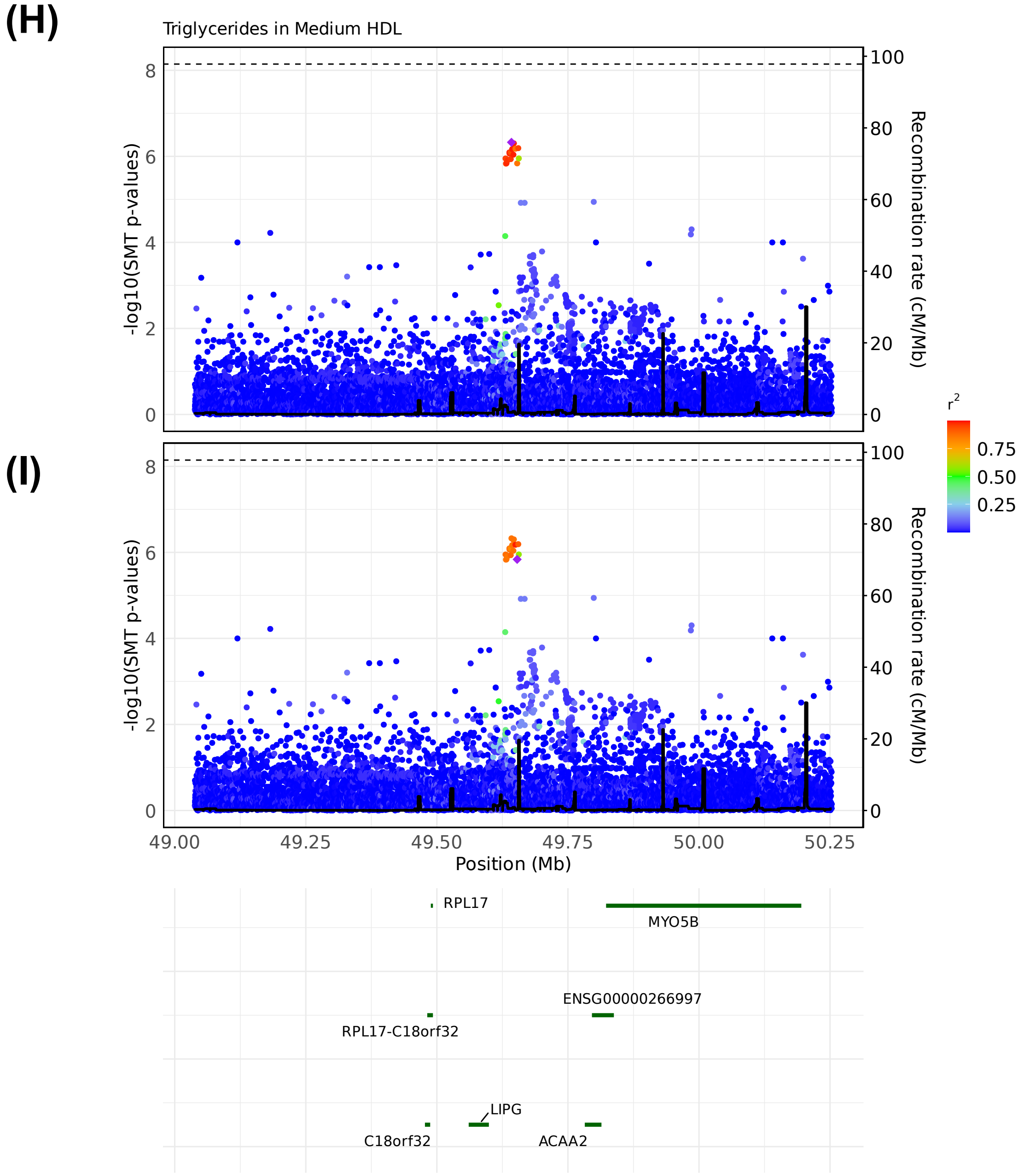


LocusZoom plots of SMT results for “triglycerides in medium HDL” at chr18:49038347-50253146. Variants are colored based on their $r^2$ with the focal marker (purple diamond), where LD is calculated in the studied samples. The black line shows the recombination rate in Finns (See Methods). Gene annotations are from GENCODE v45. **(H)** LocusZoom plot based on lead marker chr18:49642278. **(I)** LocusZoom plot based on the GWAS catalog lead marker chr18:49653146.

Figure S5J

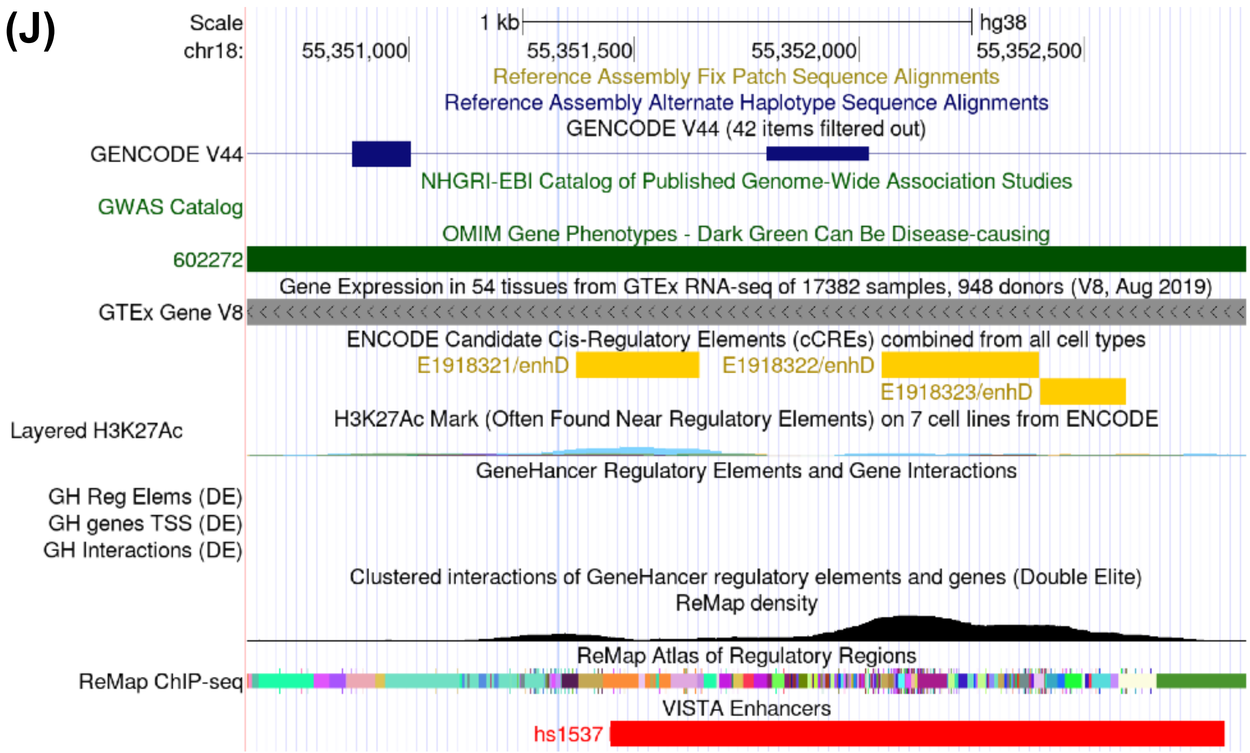


**(J)** Screenshot of the UCSC genome browser, highlighting chr18:55351333. This variant is in complete LD with sprig #384 of lead marker chr18:49653146.

Figure S6 Apolipoprotein A1 at *LIPG*
Figure S6A

**(A)**

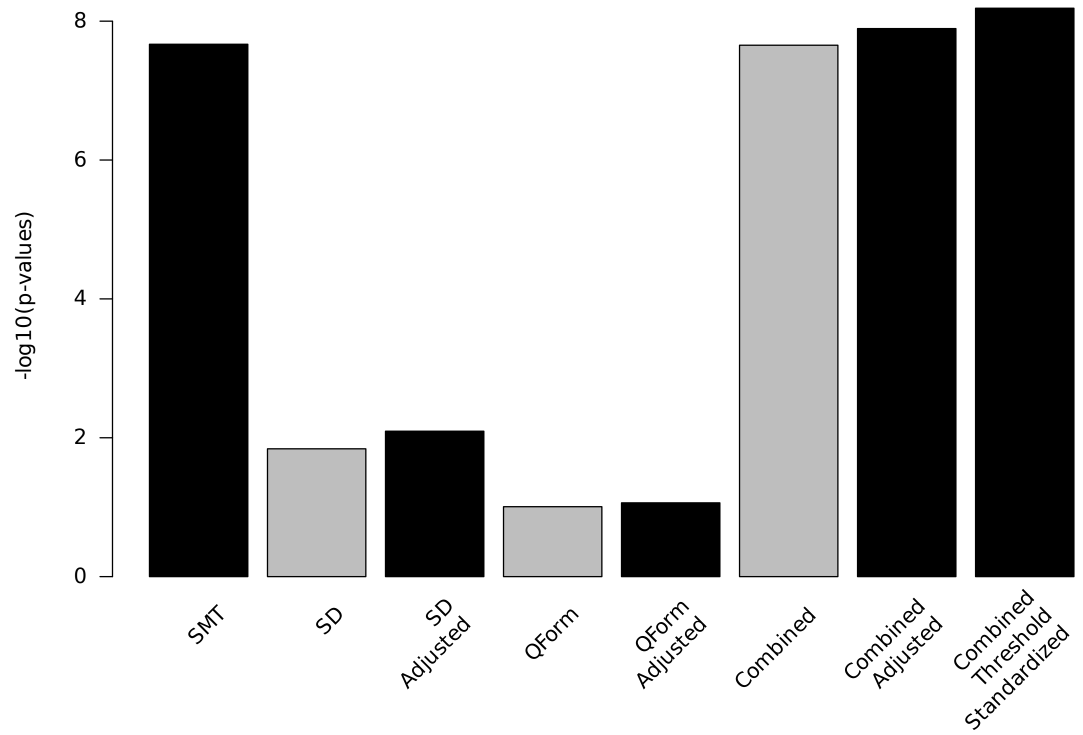


**(A)** Bar plot of $-\log_{10}(P)$ for association results before and after modified GC at lead marker chr18:49817040. Shown are the three individual sub-tests, including single marker test (SMT), stable distillation (SD) and quadratic form (Qform), as well as the three tests combined. Grey bars show $-\log_{10}(P)$ for SD, QForm and "combined" before modified GC by the slope and intercept of Q-Q plots, black bars show results after modified GC, and the final black bar at right shows the final combined $-\log_{10}(P)$ used for all final results, which also accounts for the different number of independent tests performed by SMT and LOCATER.

Figure S6B

**(B)**

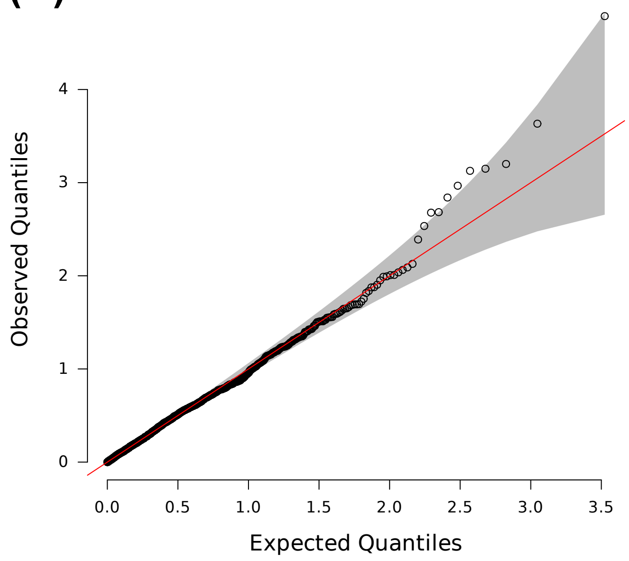


**(B)** Q-Q inflation plot of -log10(p-values) from all “sprigs” at the lead marker chr18:49817040, where “sprigs” are defined as the smallest possible inferred clades. The gray area corresponds to the 95% confidence interval, and the red line denotes x=y.

Figure S6C

**(C)**

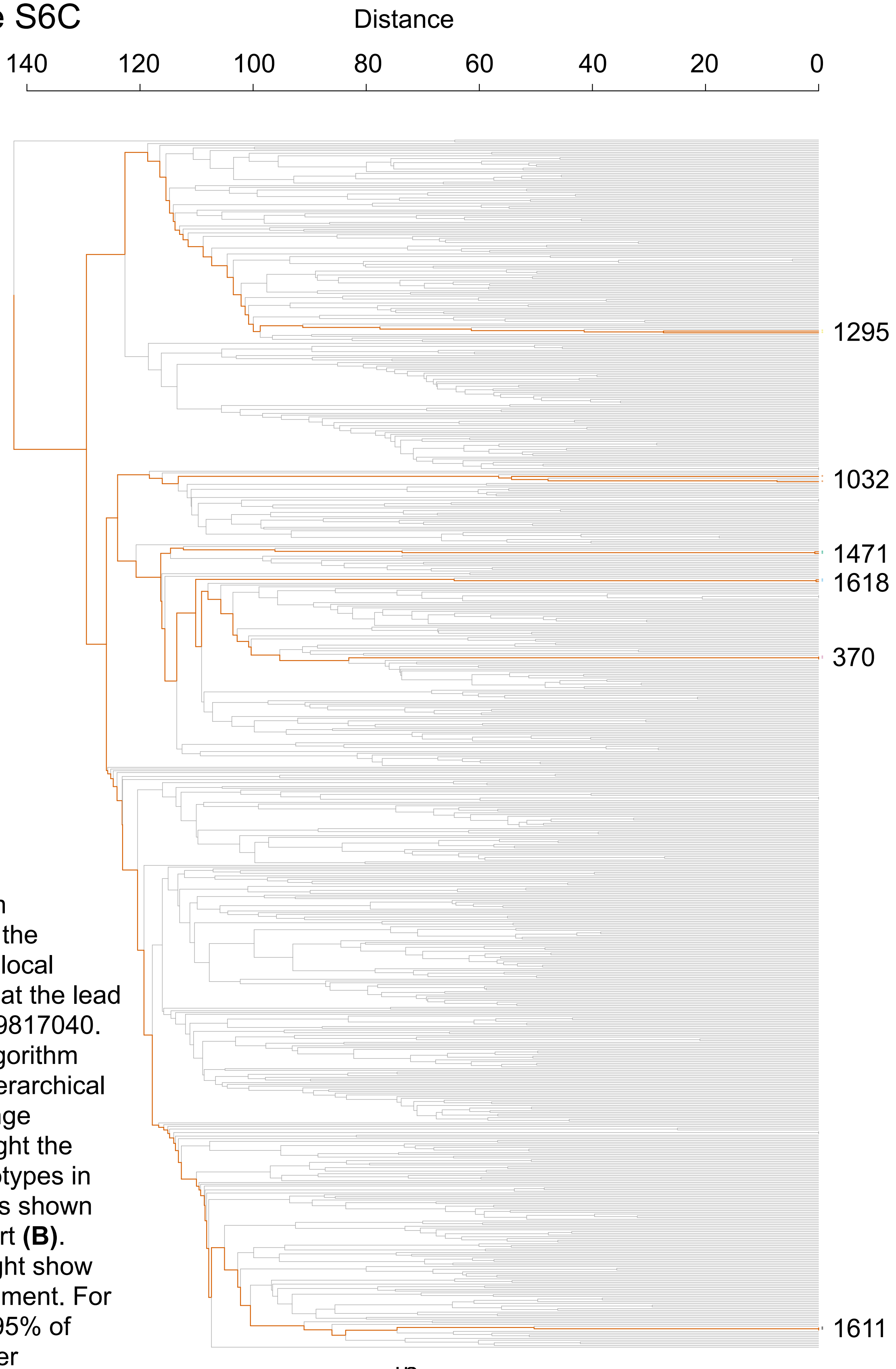


**(C)** Dendrogram generated from the haplotype-level local distance matrix at the lead marker chr18:49817040. The UPGMA algorithm was used for hierarchical clustering. Orange branches highlight the path of all haplotypes in significant sprigs shown previously in part **(B)**. Labels at the right show the sprig assignment. For plotting clarity, 95% of haplotypes under insignificant sprigs were pruned.

Figure S6D-F

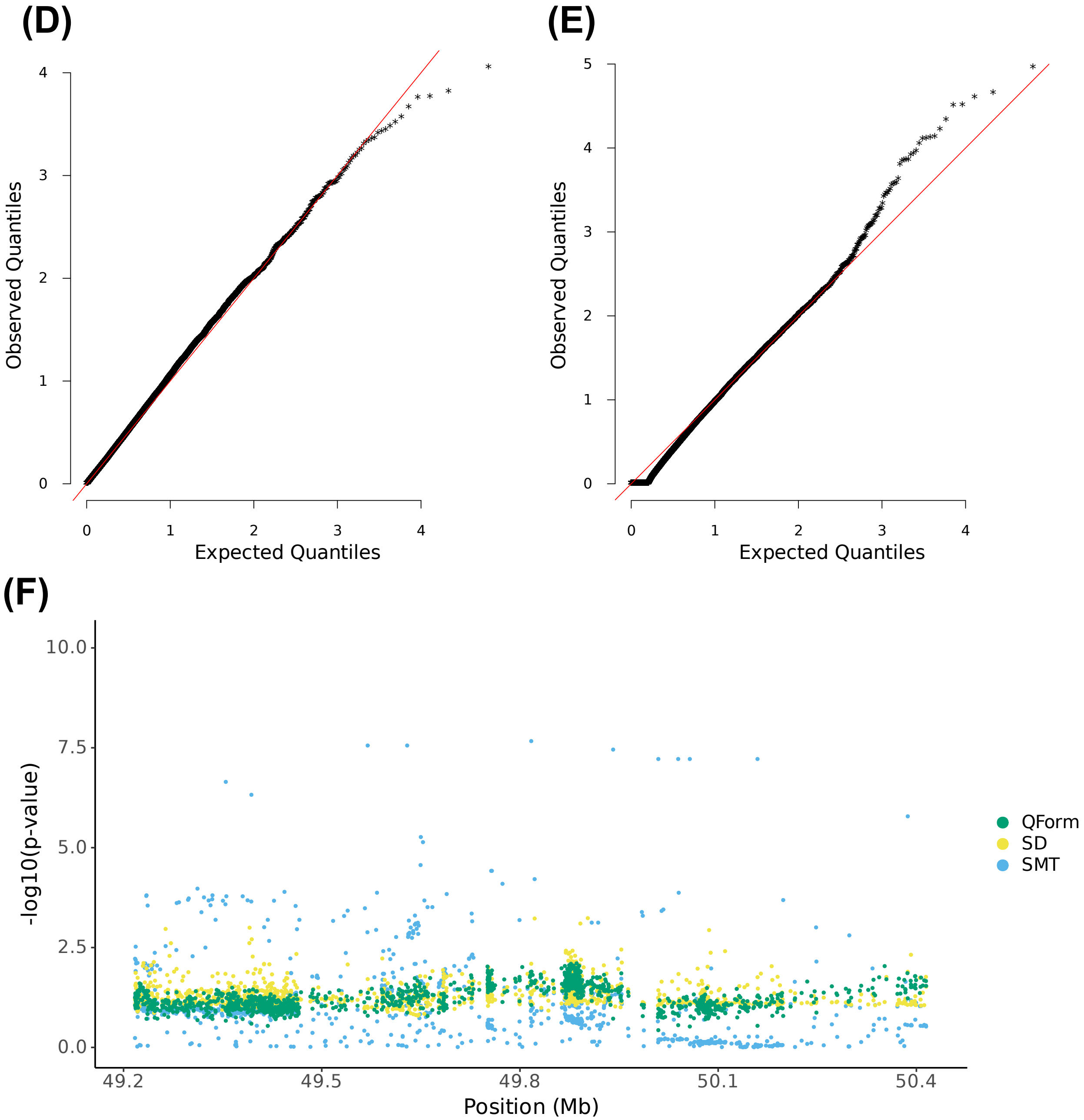


**(D)** and **(E)** are Q-Q inflation plots after modified GC of the LOCATER sub-tests stable distillation (SD) and quadratic form (QForm), respectively, for apolipoprotein A1. The red line on the diagonal corresponds to x=y. **(F)** Local Manhattan plot of apolipoprotein A1 on chr18:49217040-50417040, showing modified GCed -$\log_{10}$(P) for the 3 LOCATER sub-tests.

Figure S6G

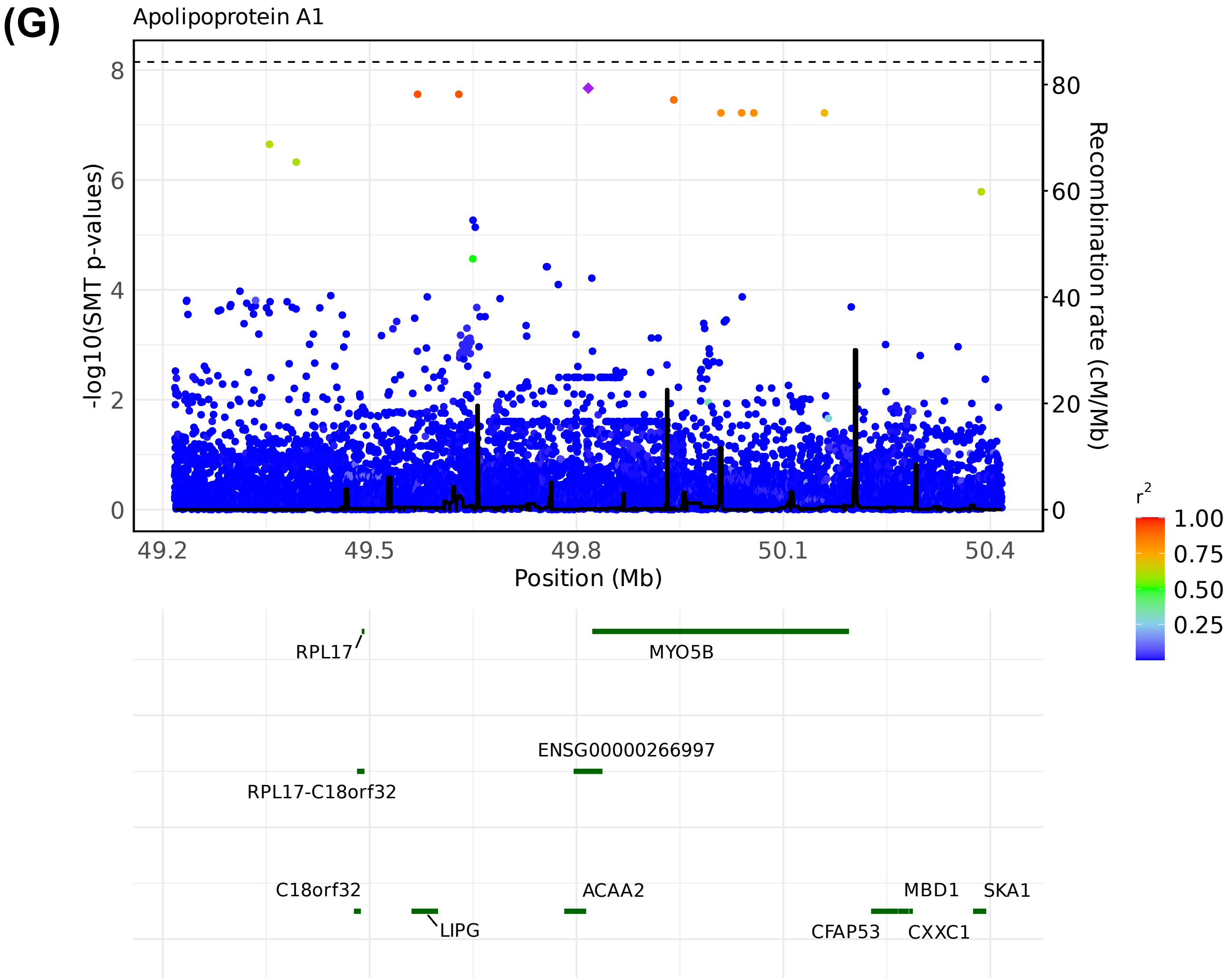


LocusZoom plots of SMT results for apolipoprotein A1 at chr18:49217040-50417040. Variants are colored based on their $r^2$ with the focal marker (purple diamond), where LD is calculated in the studied samples. The black line shows the recombination rate in Finns (See Methods). Gene annotations are from GENCODE v45. **(G)** LocusZoom plot based on lead marker chr18:49817040. This variant is also documented in GWAS catalog as associated with Apolipoprotein A1.

#### Figure S6H-I

### (H)

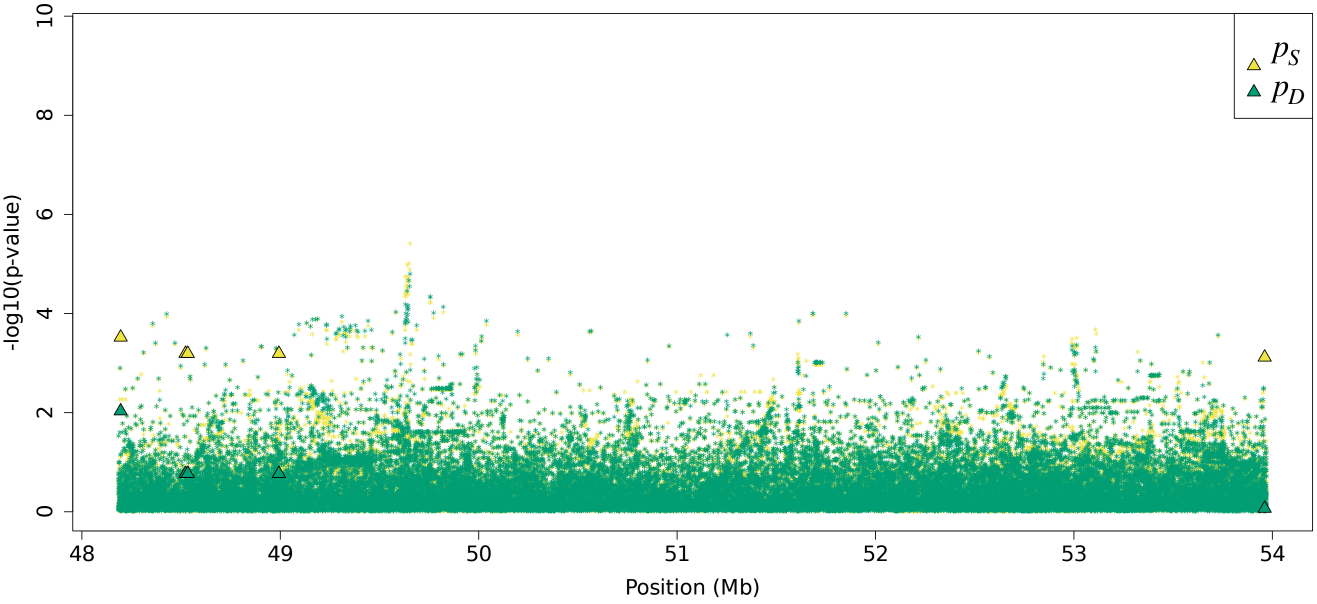


### (I)

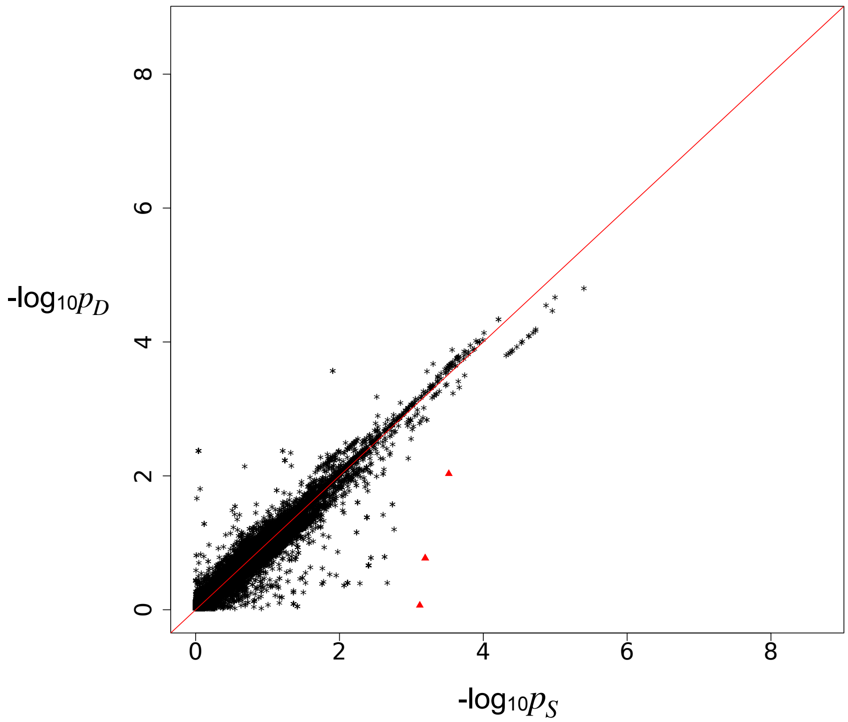


Residual association signals at the *LIPG* locus after accounting for signal from the LOCATER lead marker. $p_S$ is defined as the SMT p-value based on a residualized phenotype orthogonal against the LOCATER lead marker; $p_D$ is the SMT p-value based on a residualized phenotype orthogonal against the LOCATER lead marker and also the SD signal. The difference between $p_S$ and $p_D$ thus shows the contribution of genomic variants to the SD signal. Triangles: genomic variants with $p_S < 10^{-3}$ and $p_D > 10 * p_S$. **(H)** Local Manhattan plot of $p_S$ and $p_D$. **(I)** Scatter plot of $p_S$ and $p_D$.

Figure S6J

**(J)**

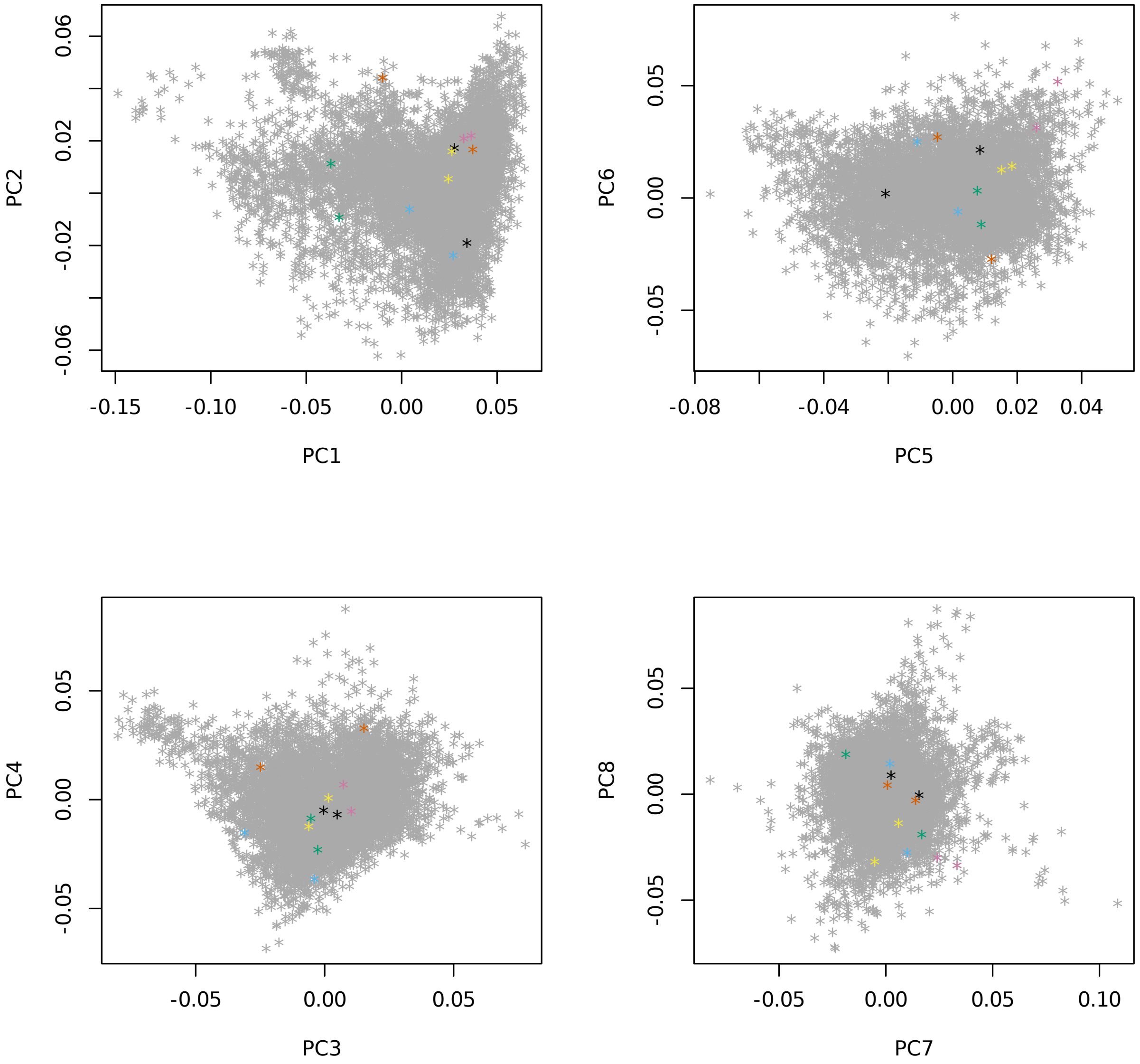


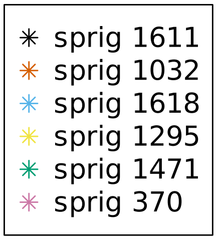


**(J)** Principal components 1-8, highlighting individuals in significant “sprigs”, where “sprigs” are defined as the smallest possible inferred clades. Individuals in the same sprig use the same color and marker.

Figure S6K
**(K)**

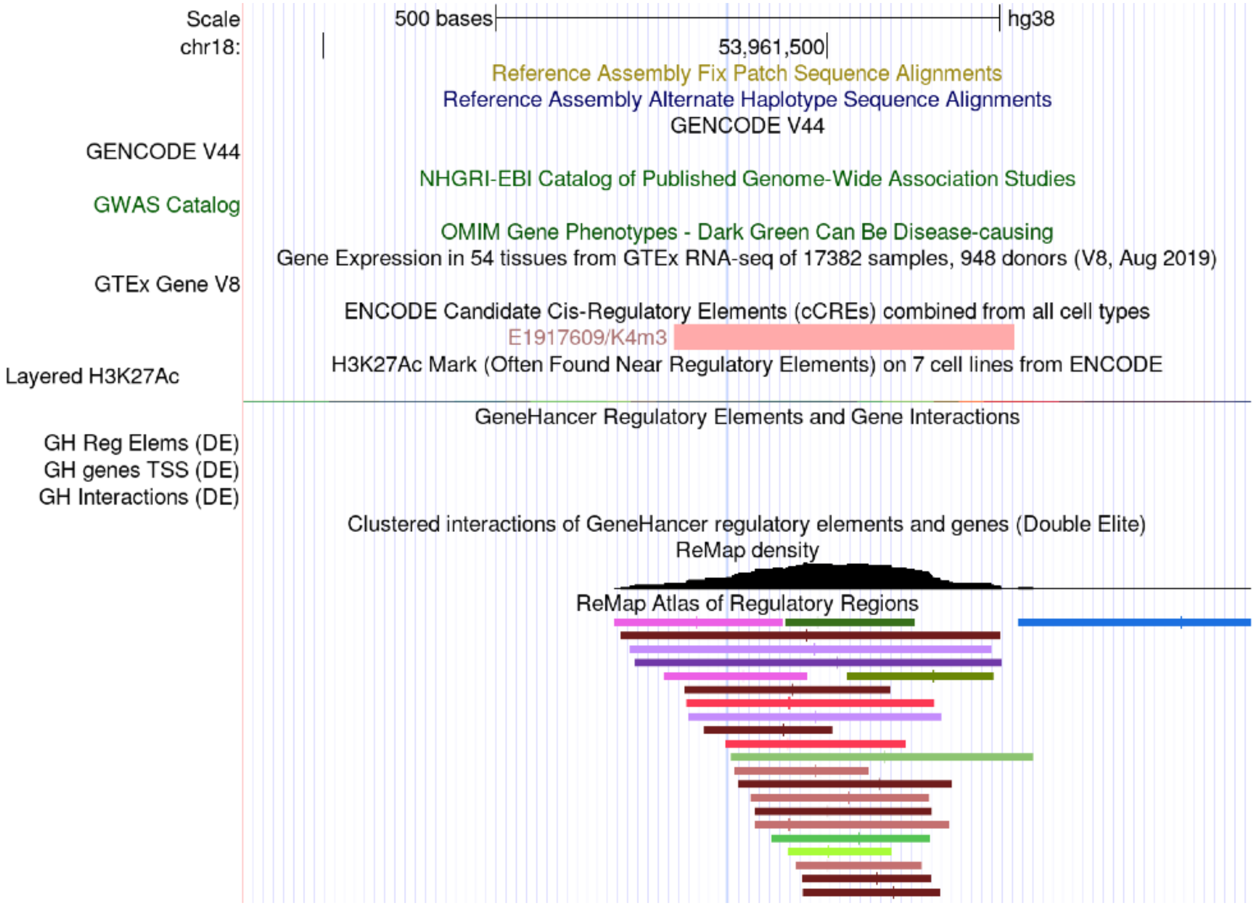


**(K)** Screenshot of the UCSC genome browser, highlighting chr18:53961401. This variant is in complete LD with sprig #1611 of lead marker chr18:49817040.

Figure S6L

**(L)**

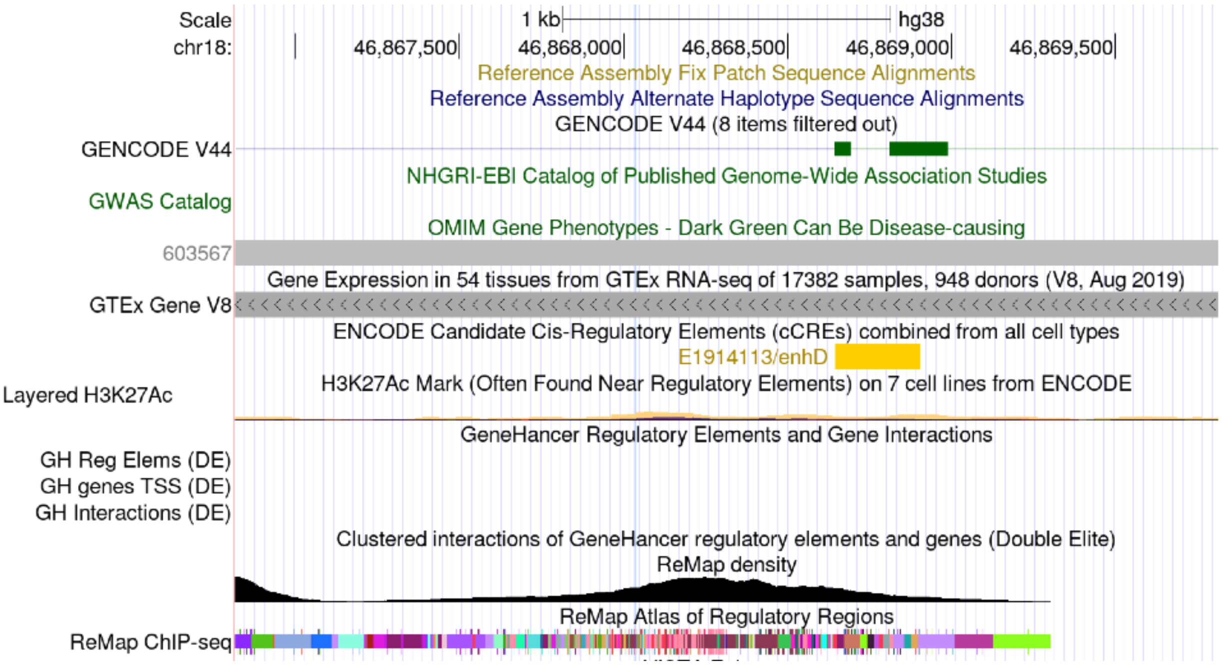


**(L)** Screenshot of the UCSC genome browser, highlighting chr18:46868035. This variant is in complete LD with sprig #370 of lead marker chr18:49817040.

Figure S7 Triglycerides in medium VLDL at *CARD18*
Figure S7A-B

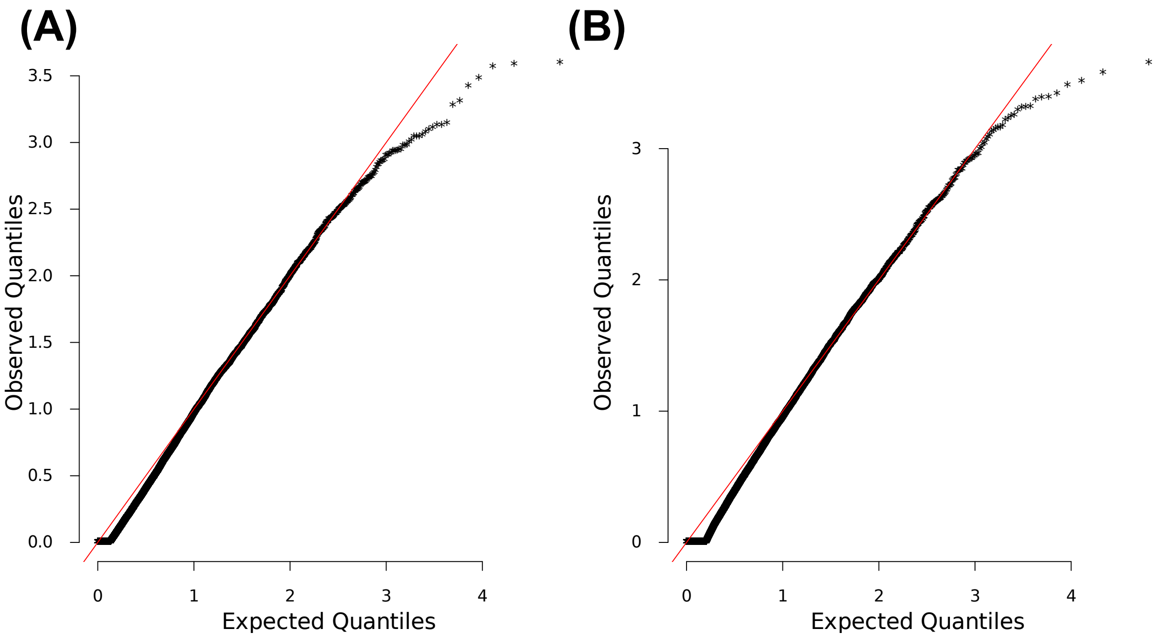


**(A)** and **(B)** are Q-Q inflation plots after modified GC of the LOCATER sub-tests stable distillation (SD) and quadratic form (QForm), respectively, for “triglycerides in medium VLDL”. The red line on the diagonal corresponds to x=y.

Figure S7C

**(C)**

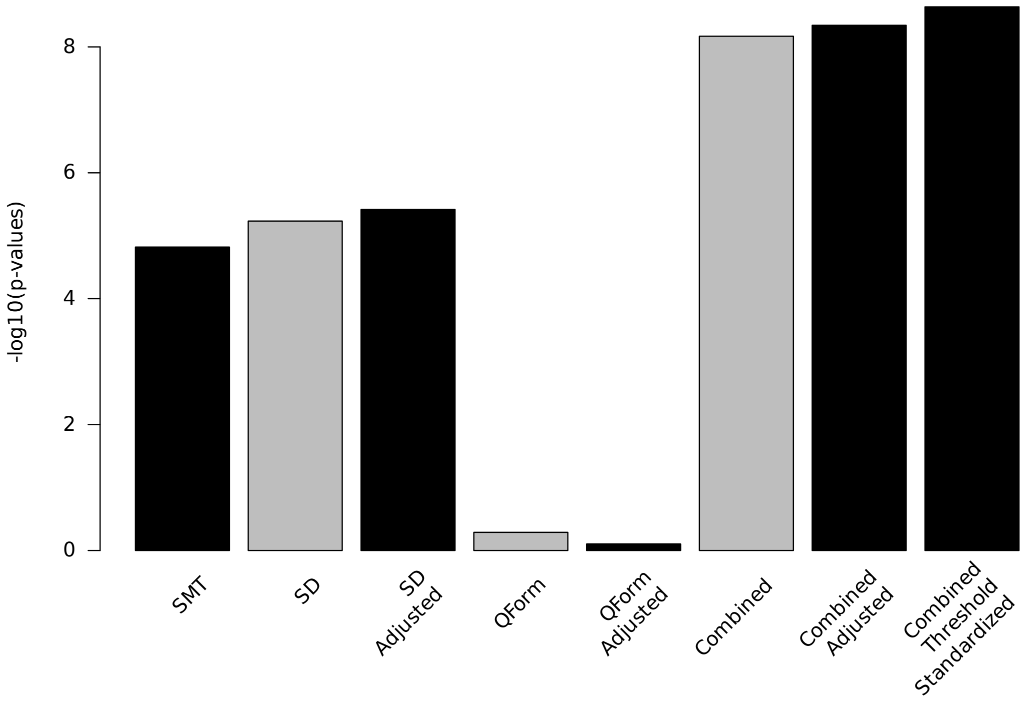


**(C)** Bar plot of -$\log_{10}$(P) for association results before and after modified GC at lead marker chr11:105327888. Shown are the three individual sub-tests, including single marker test (SMT), stable distillation (SD) and quadratic form (Qform), as well as the three tests combined. Grey bars show -$\log_{10}$(P) for SD, QForm and "combined" before modified GC by the slope and intercept of Q-Q plots, black bars show results after modified GC, and the final black bar at right shows the final combined -$\log_{10}$(P) used for all final results, which also accounts for the different number of independent tests performed by SMT and LOCATER.

Figure S7D

**(D)**

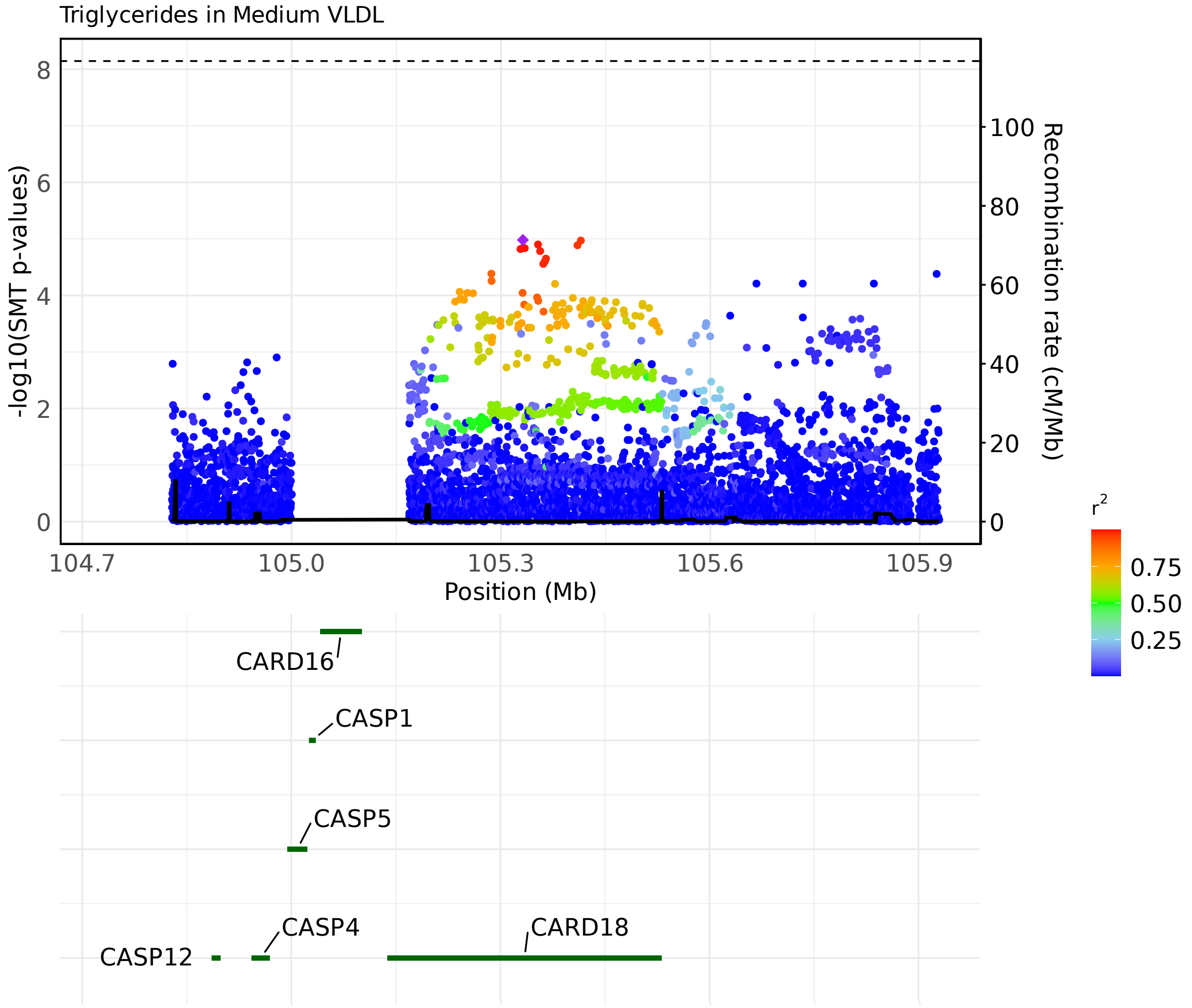


LocusZoom plots of SMT results for “triglycerides in medium VLDL” at chr11:104727888-105927888. Variants are colored based on their r² with the focal marker (purple diamond), where LD is calculated in the studied samples. The black line shows the recombination rate in Finns (See Methods). Gene annotations are from GENCODE v45. **(D)** LocusZoom plot based on lead marker chr11:105331384.

Figure S7E-F

**(E)**

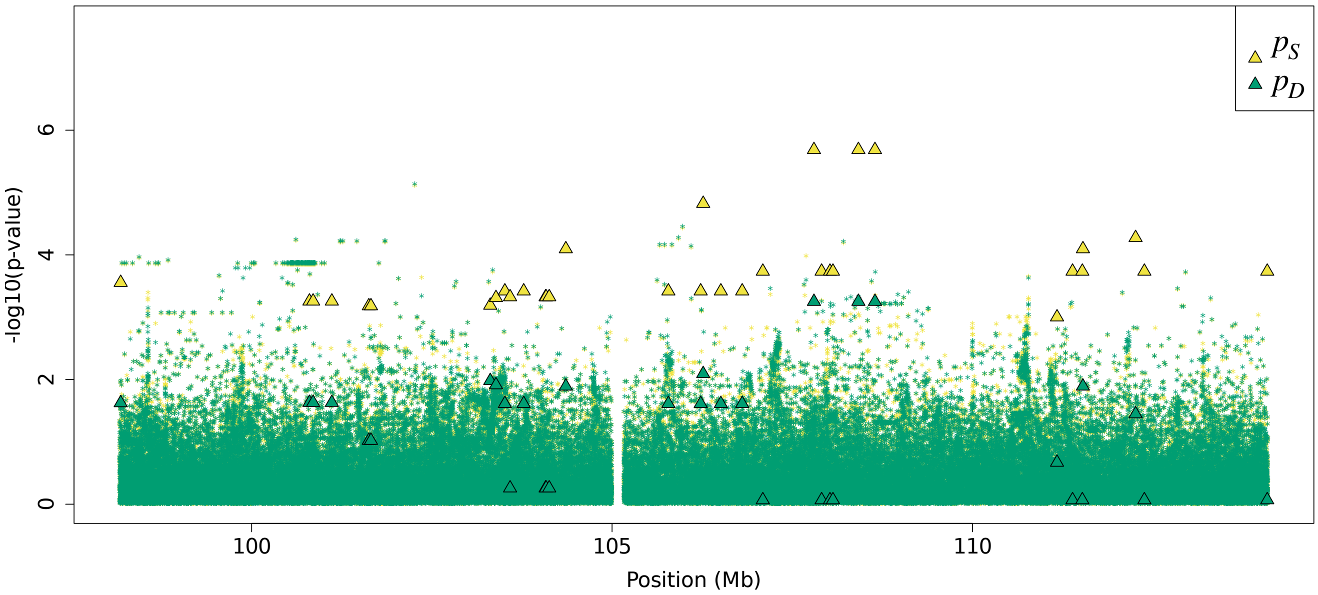


**(F)**

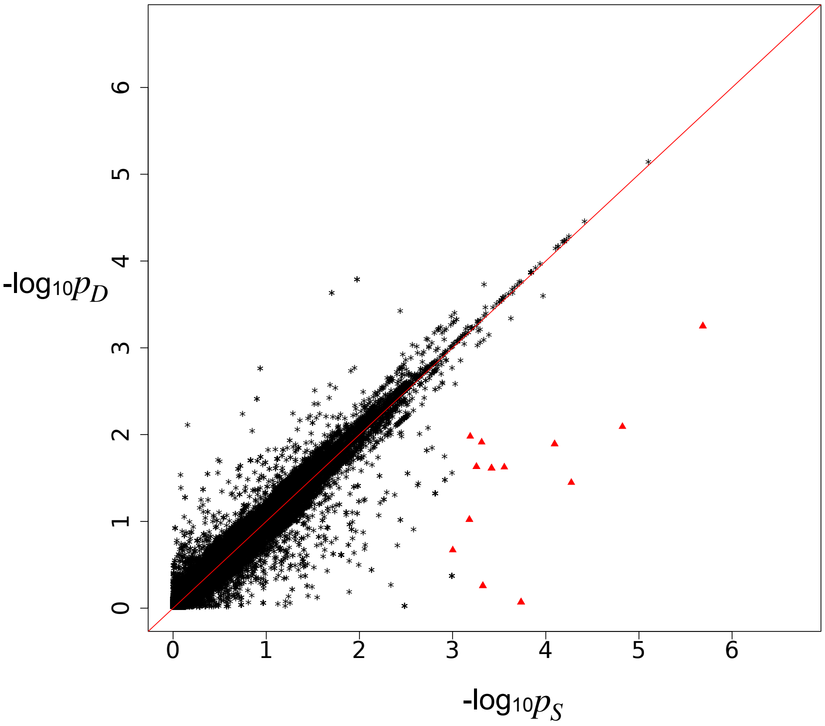


Residual association signals at chr11:98173184-114099685 after accounting for signal from the LOCATER lead marker. $p_S$ is defined as the SMT p-value based on a residualized phenotype orthogonal against the LOCATER lead marker; $p_D$ is the SMT p-value based on a residualized phenotype orthogonal against the LOCATER lead marker and also the SD signal. The difference between $p_S$ and $p_D$ thus shows the contribution of genomic variants to the SD signal. Triangles: genomic variants with $p_S < 10^{-3}$ and $p_D > 10 * p_S$. **(E)** Local Manhattan plot of $p_S$ and $p_D$. **(F)** Scatter plot of $p_S$ and $p_D$.

Figure S7G

**(G)**

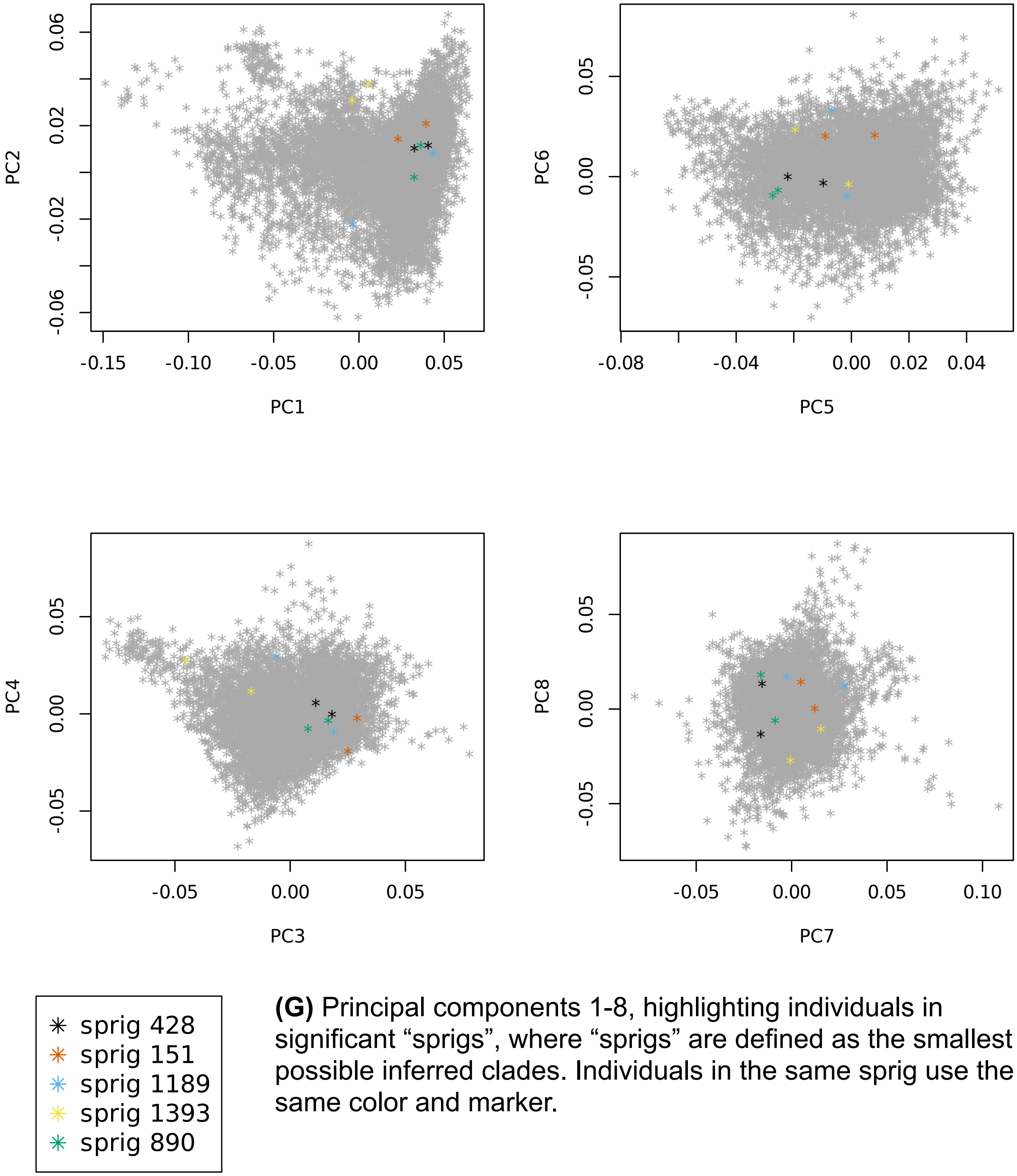


**(G)** Principal components 1-8, highlighting individuals in significant "sprigs", where "sprigs" are defined as the smallest possible inferred clades. Individuals in the same sprig use the same color and marker.

Figure S8 Remnant cholesterol at *APOE* cluster
Figure S8A-B

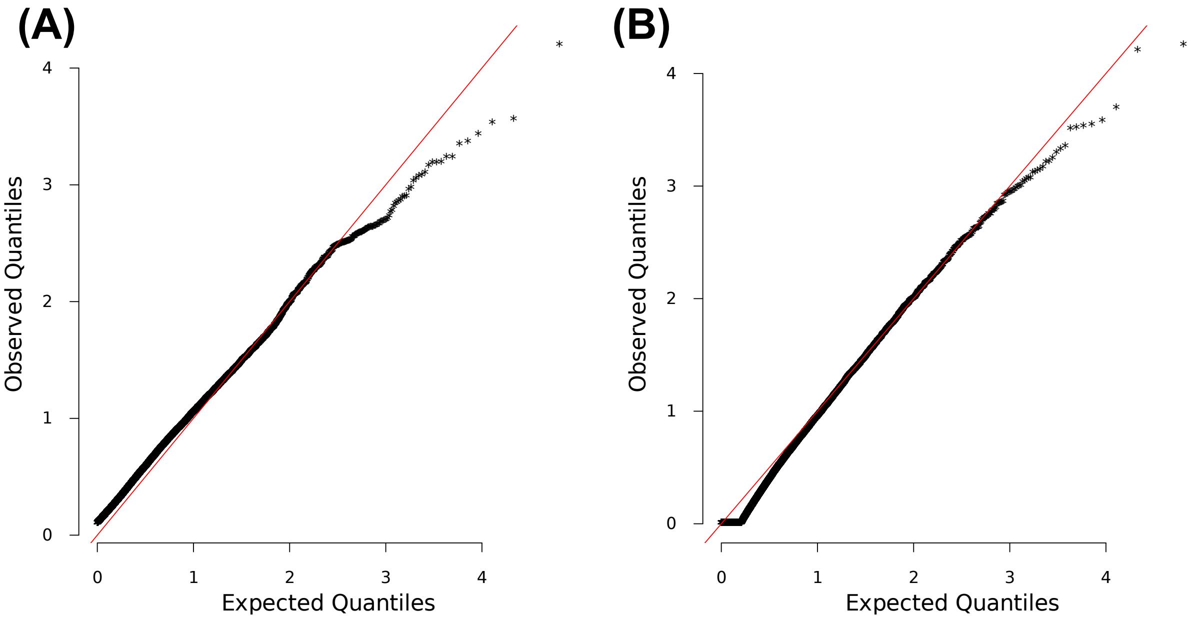


**(A)** and **(B)** are Q-Q inflation plots after modified GC of the LOCATER sub-tests stable distillation (SD) and quadratic form (QForm), respectively, for remnant cholesterol. The red line on the diagonal corresponds to x=y.

Figure S8C

**(C)**

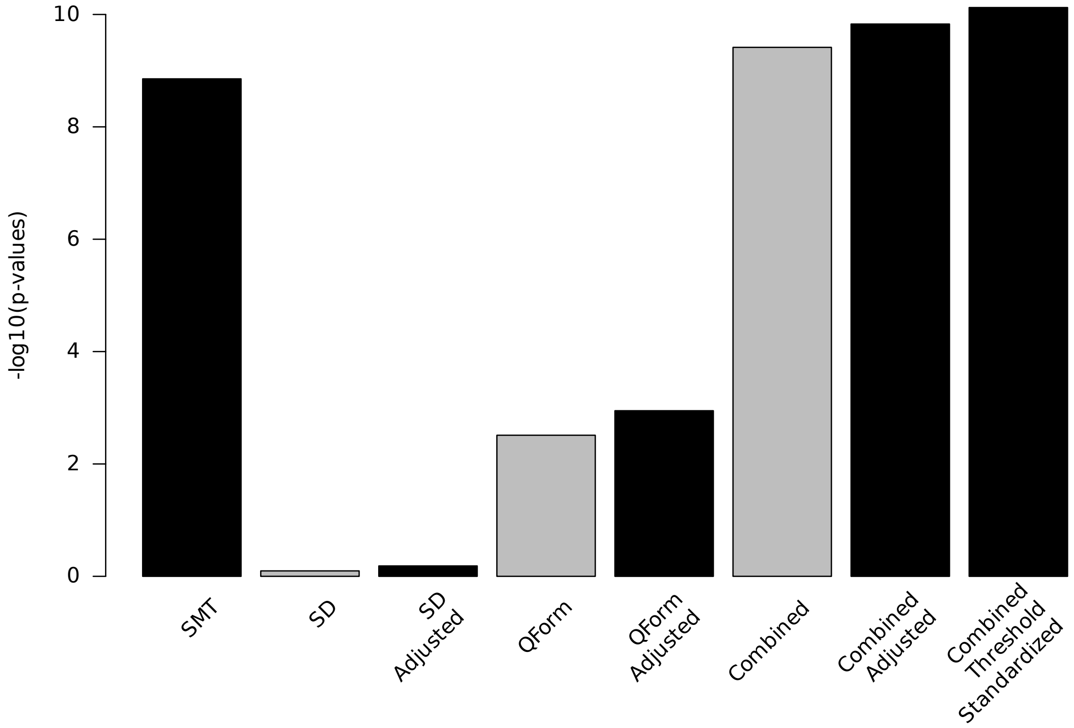


**(C)** Bar plot of $-\log_{10}(P)$ for association results before and after modified GC at lead marker chr19:44922203. Shown are the three individual sub-tests, including single marker test (SMT), stable distillation (SD) and quadratic form (Qform), as well as the three tests combined. Grey bars show $-\log_{10}(P)$ for SD, QForm and "combined" before modified GC by the slope and intercept of Q-Q plots, black bars show results after modified GC, and the final black bar at right shows the final combined $-\log_{10}(P)$ used for all final results, which also accounts for the different number of independent tests performed by SMT and LOCATER.

#### Figure S8D-G

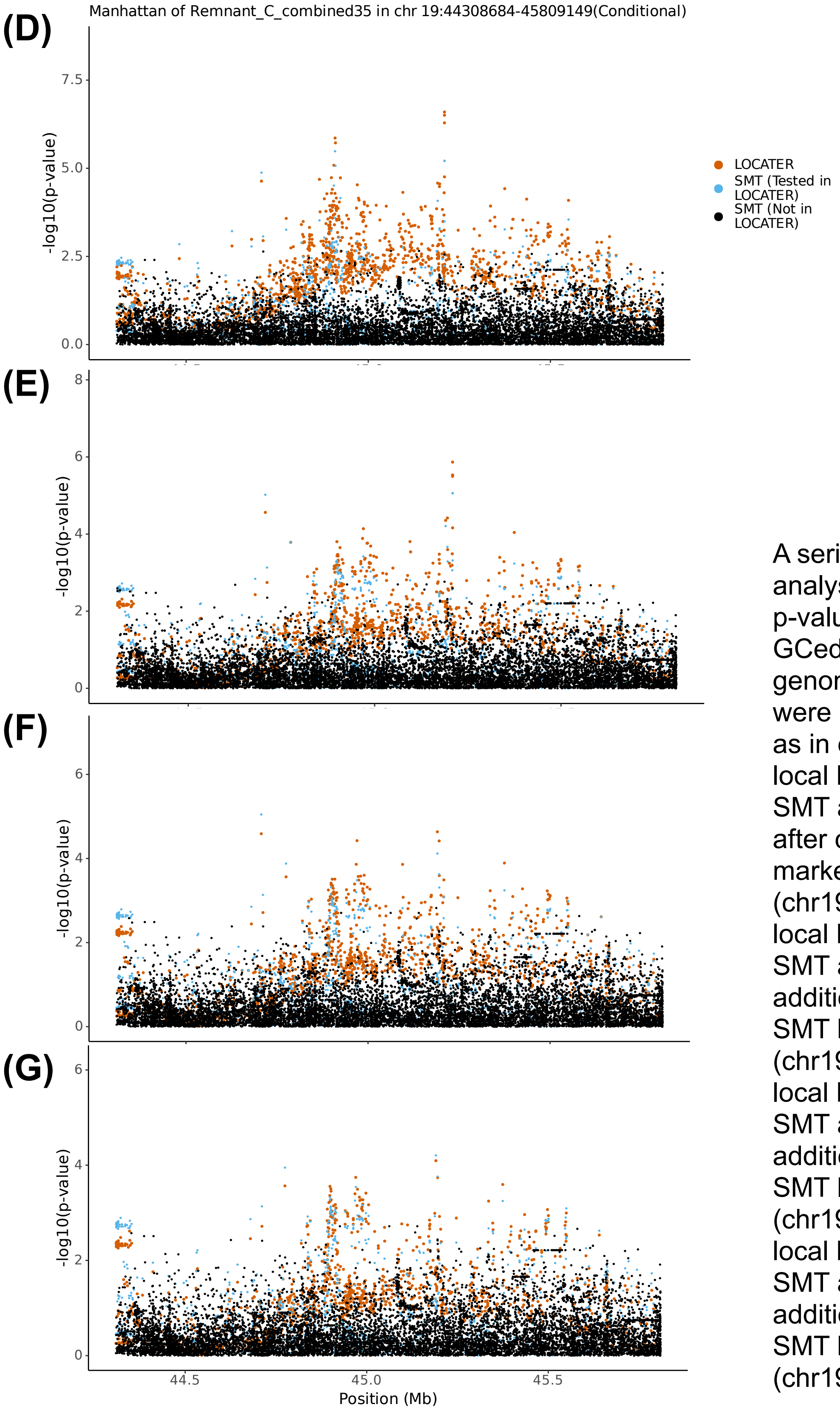


A series of conditional analyses. All LOCATER p-values were modified GCed based on general genomic control (but were not standardized as in other figures). **(D)** local Manhattan plot of SMT and LOCATER after condition on lead marker (chr19:44922203). **(E)** local Manhattan plot of SMT and LOCATER additionally condition on SMT lead marker in **(D)** (chr19:44908822) **(F)** local Manhattan plot of SMT and LOCATER additionally condition on SMT lead marker in **(E)** (chr19:45209149) **(G)** local Manhattan plot of SMT and LOCATER additionally condition on SMT lead marker in **(F)** (chr19:44706760)

### Figure S9 MUFA on chr7
### Figure S9A-C

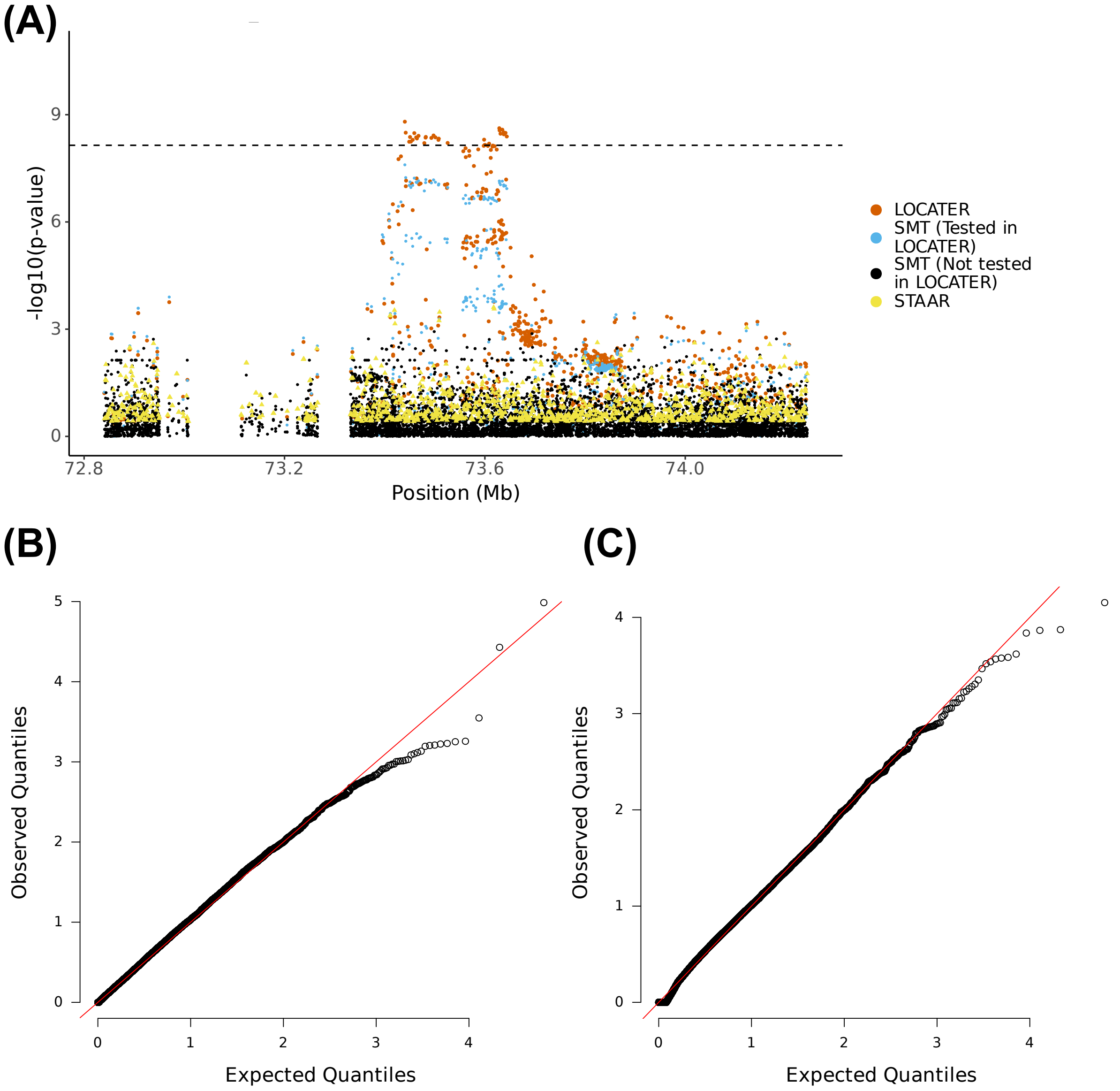


**(A)** Local Manhattan plot of the association signal for monounsaturated fatty acids (MUFA) on chr7:72840219-74243687, including results for single marker test (SMT; blue and black), LOCATER (orange) and STAAR (yellow). Note that LOCATER results are only shown for variants with an SMT p-value less than $1x10^{-3}$, since for computational efficiency only these variants were tested by LOCATER (see Methods). SMT results from variants tested by LOCATER are shown in blue, and those from variants not tested by LOCATER are shown in black. The black dashed line corresponds to the genome-wide significance threshold for SMT, standardized LOCATER, and standardized STAAR. **(B)** and **(C)** are Q-Q inflation plots after modified GC of the LOCATER sub-tests stable distillation (SD) and quadratic form (QForm), respectively, for MUFA. The red line on the diagonal corresponds to x=y.

Figure S9D-E

(D)

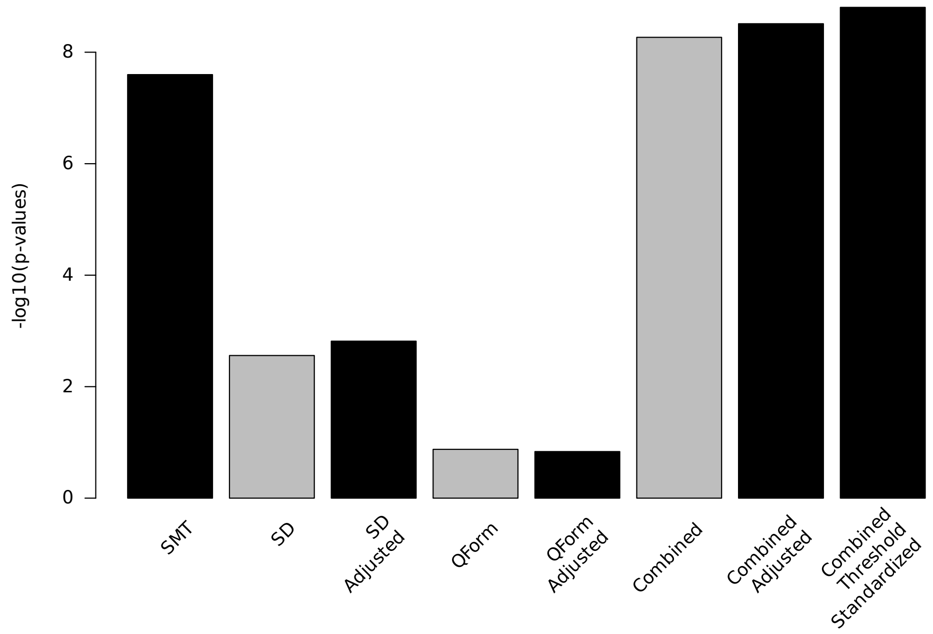


(E)

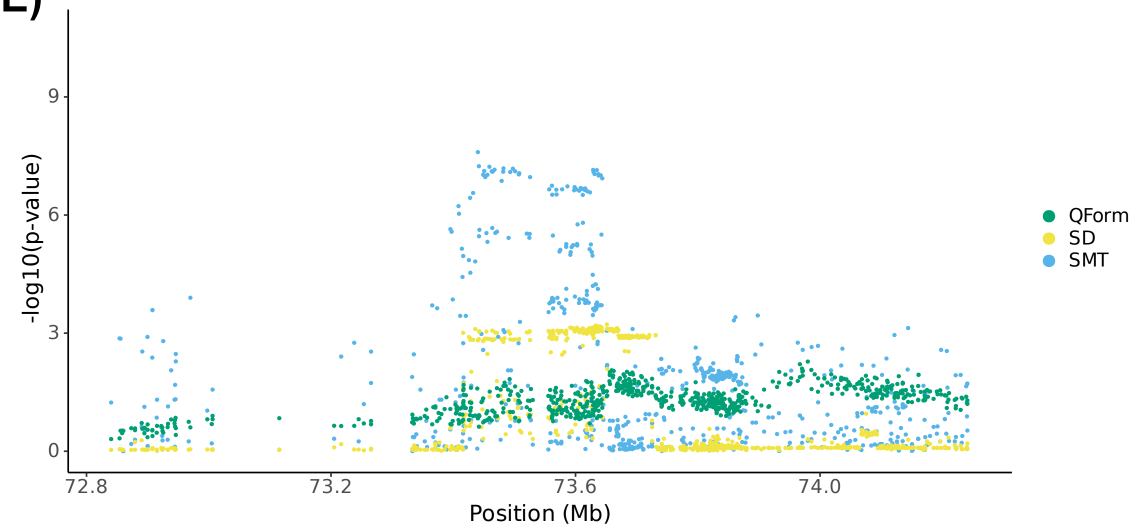


**(D)** Bar plot of -log$_{10}$(P) for association results before and after modified GC at lead marker chr7:73440219. Shown are the three individual sub-tests, including single marker test (SMT), stable distillation (SD) and quadratic form (Qform), as well as the three tests combined. Grey bars show -log$_{10}$(P) for SD, QForm and “combined” before modified GC by the slope and intercept of Q-Q plots, black bars show results after modified GC, and the final black bar at right shows the final combined -log$_{10}$(P) used for all final results, which also accounts for the different number of independent tests performed by SMT and LOCATER. **(E)** Local Manhattan plot of MUFA on chr7:72840219-74243687, showing modified GCed -log$_{10}$(P) for the 3 LOCATER sub-tests.

Figure S9F-G

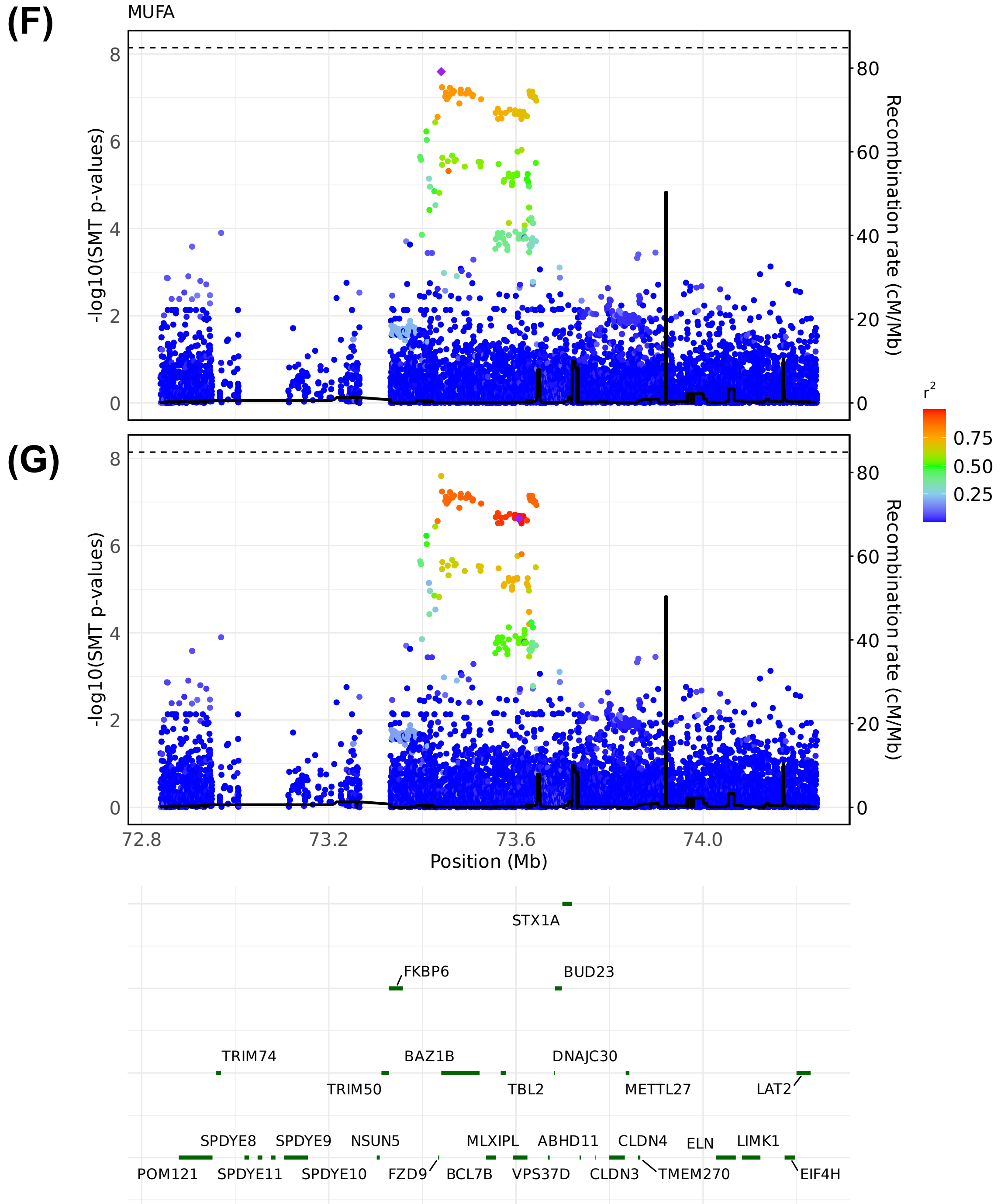


LocusZoom plots of SMT results for MUFA at chr7:72840219-74243687. Variants are colored based on their $r^2$ with the focal marker (purple diamond), where LD is calculated in the studied samples. The black line shows the recombination rate in Finns (See Methods). Gene annotations are from GENCODE v45. **(F)** LocusZoom plot based on lead marker chr7:73440219. **(G)** LocusZoom plot based on GWAS catalog lead marker chr7:73606007.

#### Figure S9H-I

**(H)**

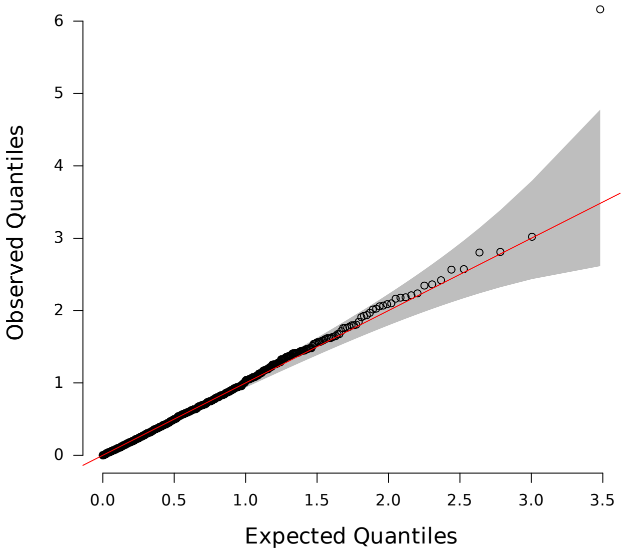


**(H)** Q-Q inflation plot of -log10(p-values) from all “sprigs” at the lead marker chr7:73440219, where “sprigs” are defined as the smallest possible inferred clades. The gray area corresponds to the 95% confidence interval, and the red line denotes x=y. **(I)** Histogram of phenotype values after projecting out the genotype vector of the LOCATER lead marker (chr7:73440219), thus removing signal that can be accounted for by the SMT sub-test. Connected dots show the phenotype value of individuals assigned to significant sprigs.

**(I)**

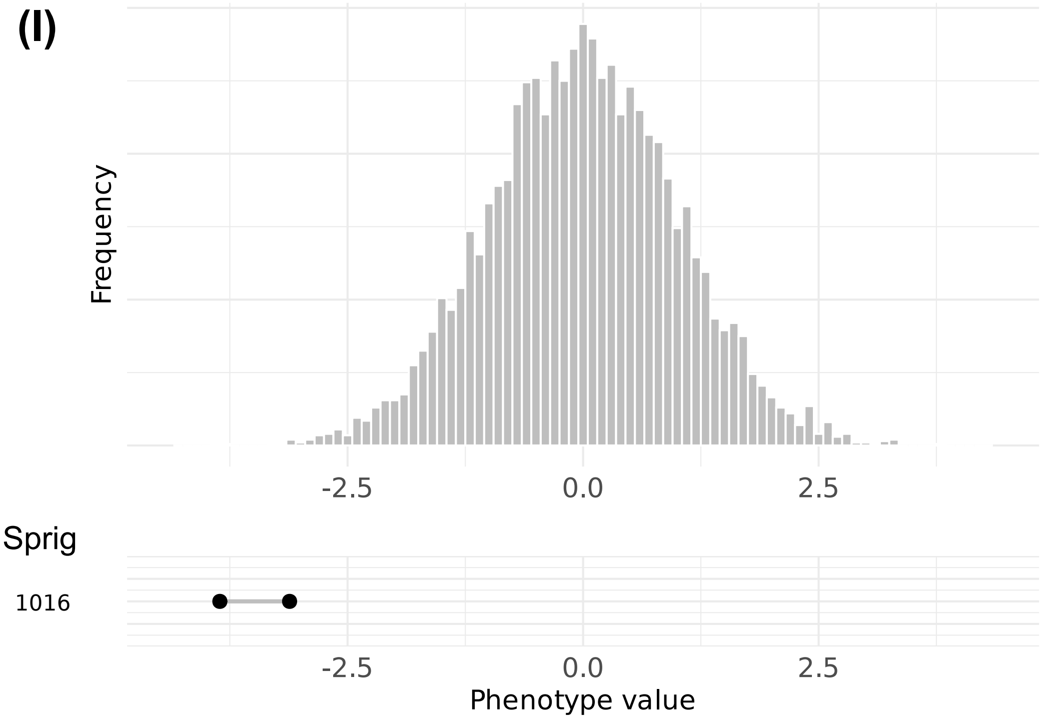

Figure S9J

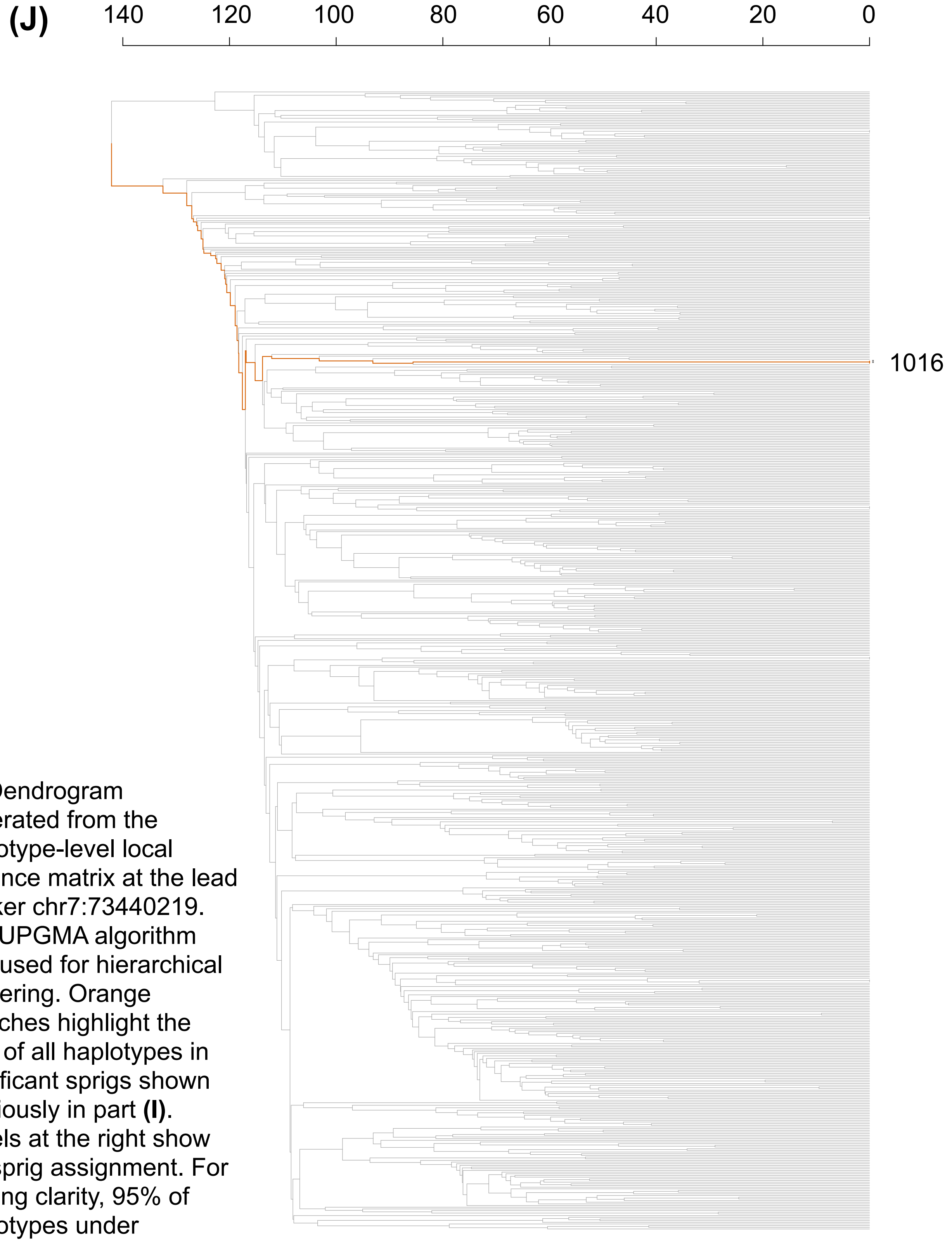


**(J)** Dendrogram generated from the haplotype-level local distance matrix at the lead marker chr7:73440219. The UPGMA algorithm was used for hierarchical clustering. Orange branches highlight the path of all haplotypes in significant sprigs shown previously in part **(I)**. Labels at the right show the sprig assignment. For plotting clarity, 95% of haplotypes under insignificant sprigs were pruned.

Figure S9K-L

**(K)**

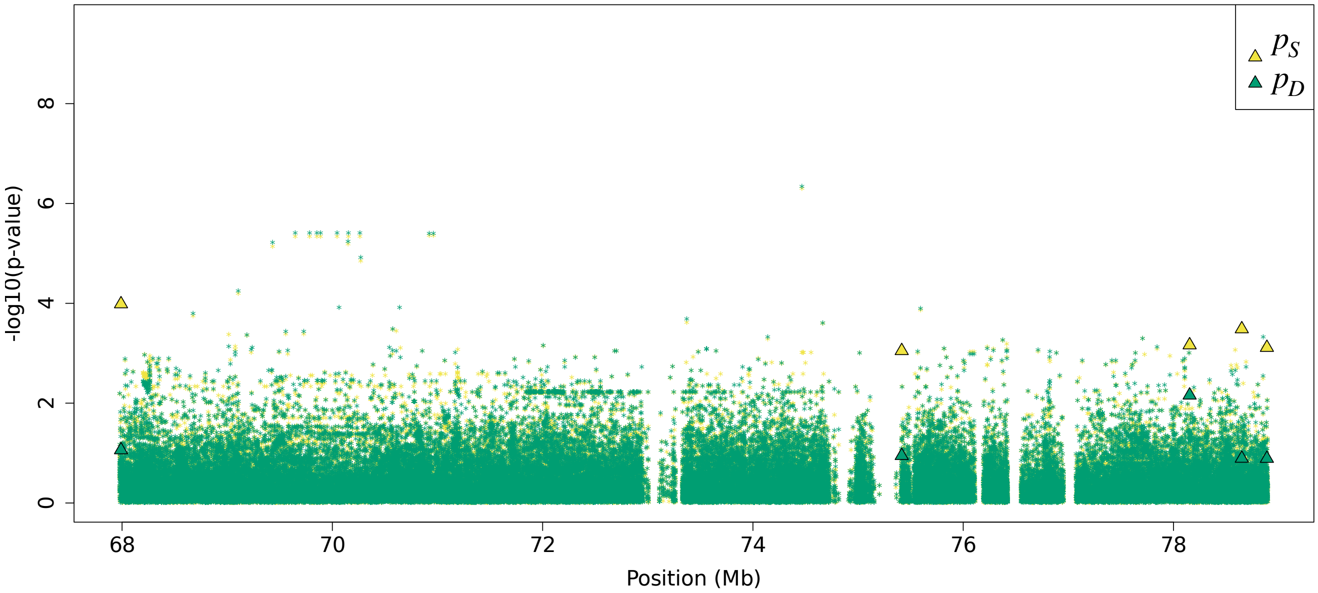


**(L)**

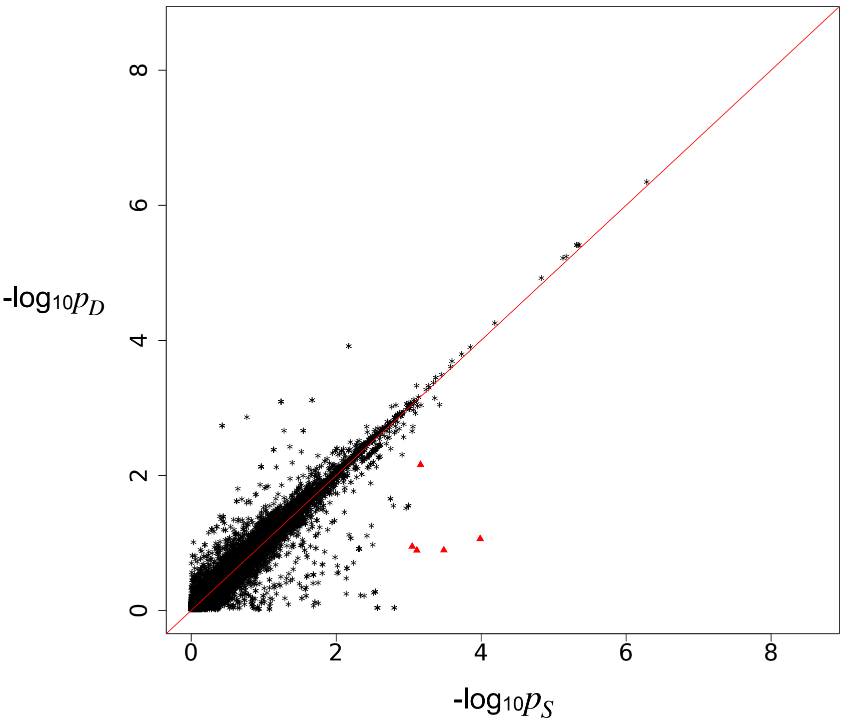


Residual association signals at the BAZ1B locus after accounting for signal from the LOCATER lead marker. $p_S$ is defined as the SMT p-value based on a residualized phenotype orthogonal against the LOCATER lead marker; $p_D$ is the SMT p-value based on a residualized phenotype orthogonal against the LOCATER lead marker and also the SD signal. The difference between $p_S$ and $p_D$ thus shows the contribution of genomic variants to the SD signal. Triangles: genomic variants with $p_S < 10^{-3}$ and $p_D > 10 * p_S$. **(K)** Local Manhattan plot of $p_S$ and $p_D$. **(L)** Scatter plot of $p_S$ and $p_D$.

Figure S9M

**(M)**

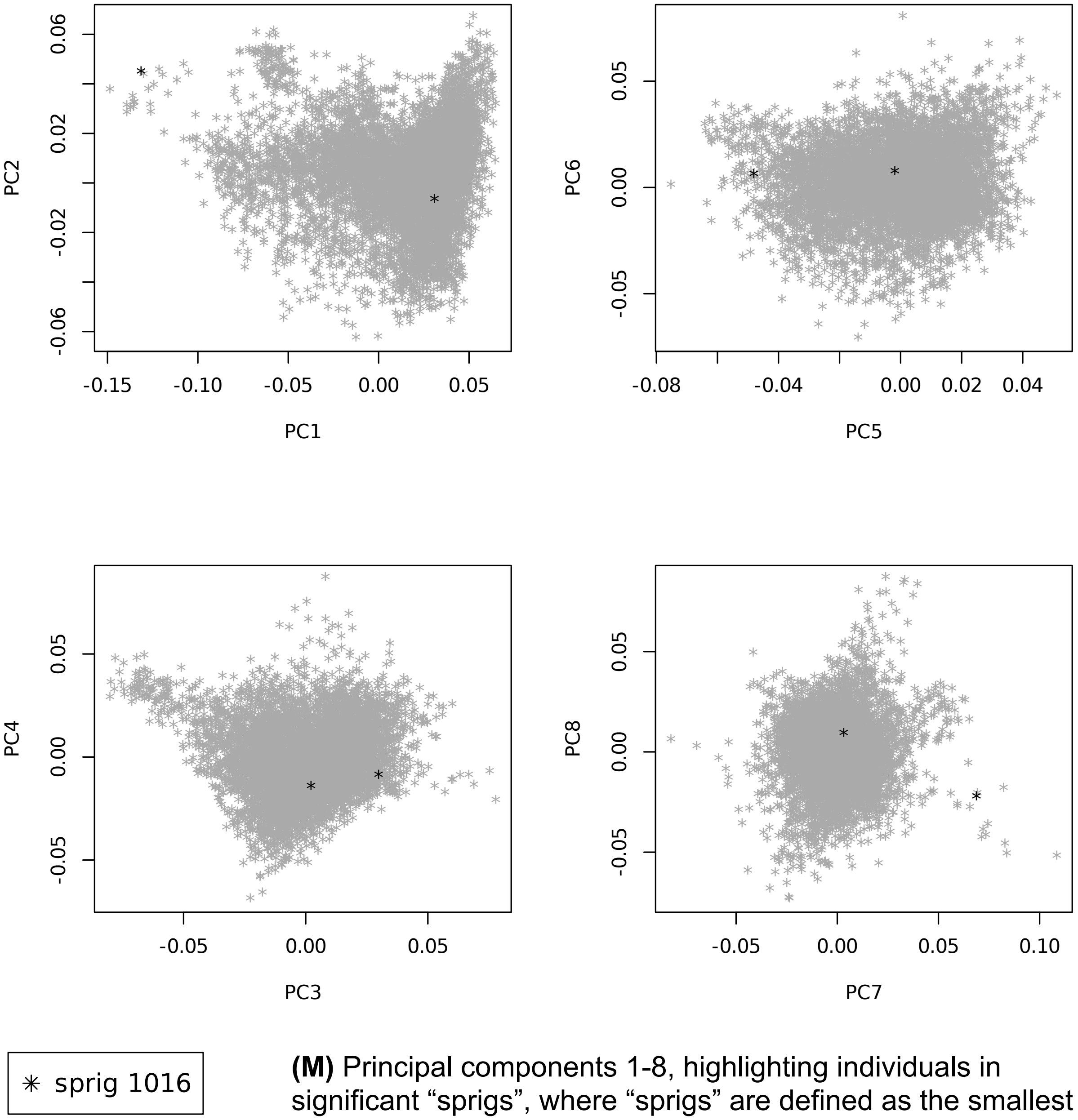


**(M)** Principal components 1-8, highlighting individuals in significant “sprigs”, where “sprigs” are defined as the smallest possible inferred clades. Individuals in the same sprig use the same color and marker.

Figure S10 Triglycerides in small HDL at *PLTP* locus
Figure S10A-C

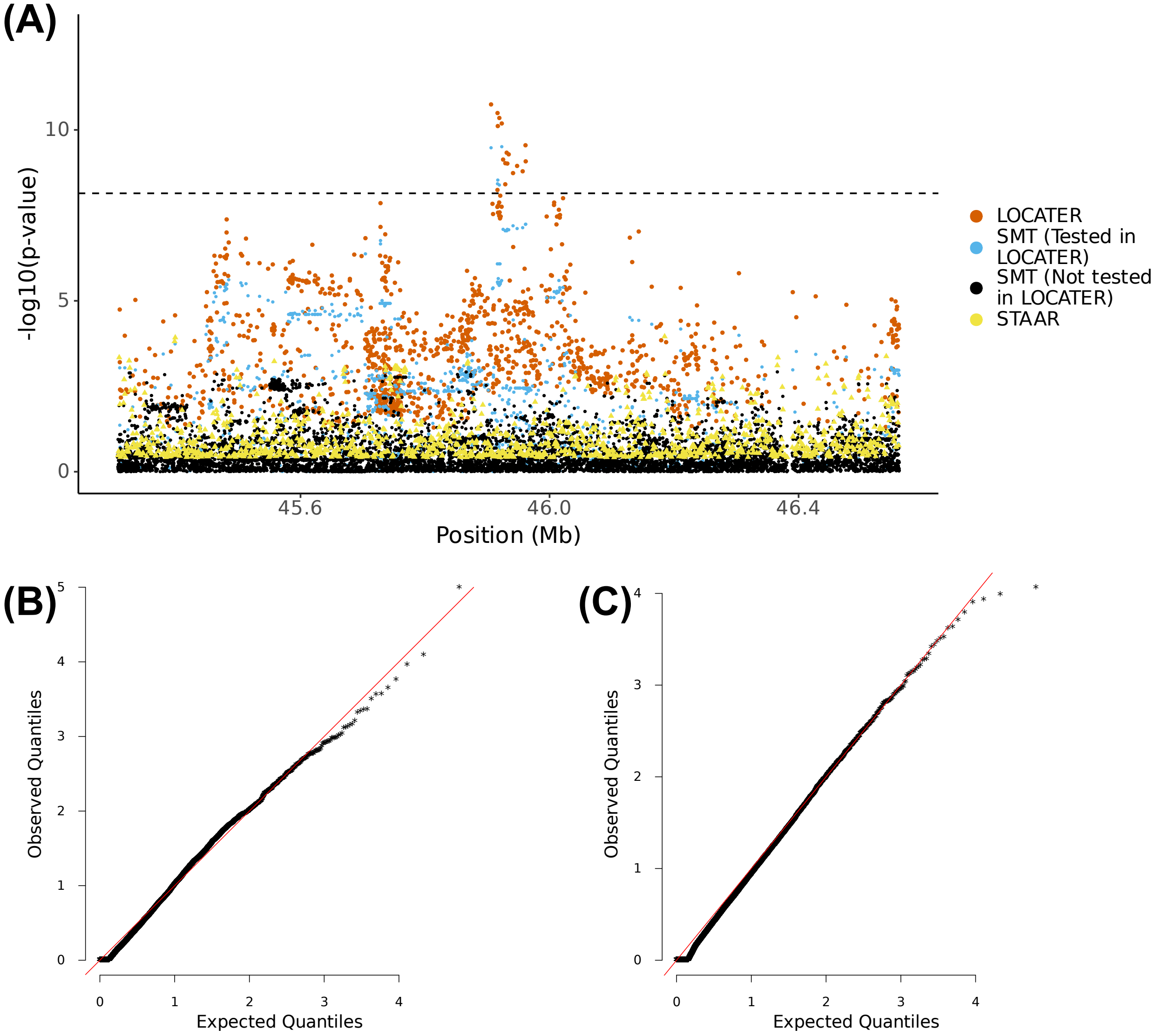


**(A)** Local Manhattan plot of the association signal for "triglycerides in small HDL" on chr20:45306012-46561659, including results for single marker test (SMT; blue and black), LOCATER (orange) and STAAR (yellow). Note that LOCATER results are only shown for variants with an SMT p-value less than $1x10^{-3}$, since for computational efficiency only these variants were tested by LOCATER (see Methods). SMT results from variants tested by LOCATER are shown in blue, and those from variants not tested by LOCATER are shown in black. The black dashed line corresponds to the genome-wide significance threshold for SMT, standardized LOCATER, and standardized STAAR. **(B)** and **(C)** are Q-Q inflation plots after modified GC of the LOCATER sub-tests stable distillation (SD) and quadratic form (QForm), respectively, for "triglycerides in small HDL". The red line on the diagonal corresponds to x=y.

#### Figure S10D-E

**(D)**

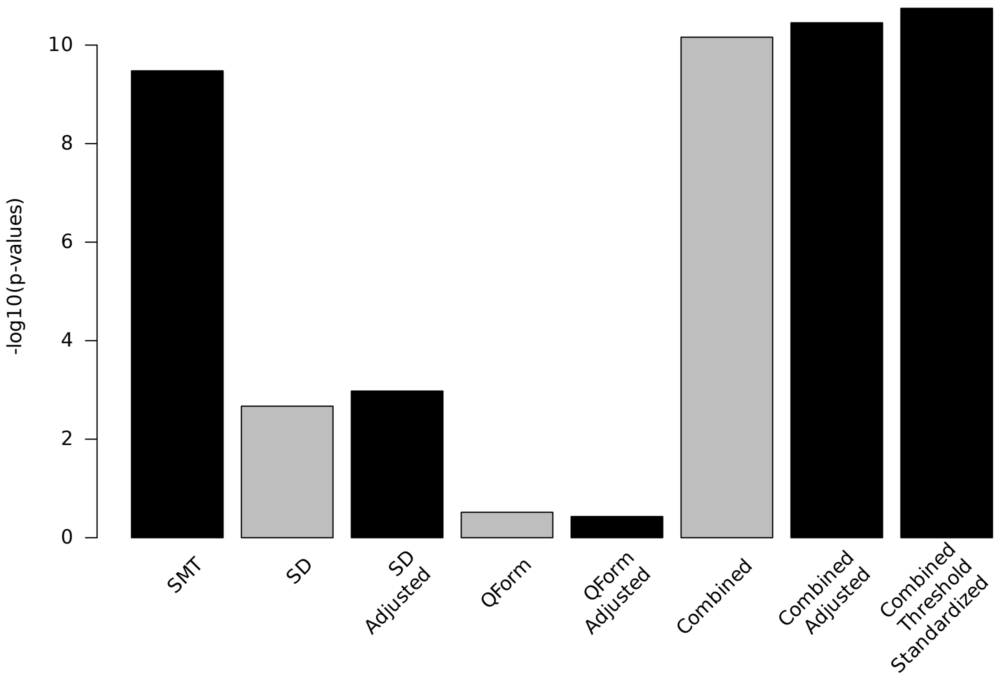


**(E)**

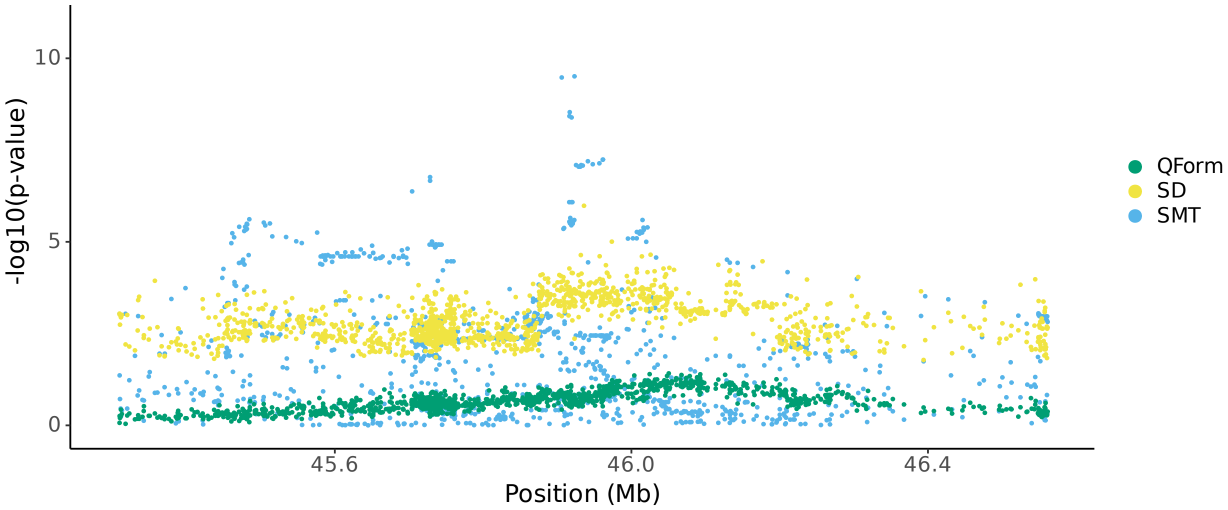


**(D)** Bar plot of -log$_{10}$(P) for association results before and after modified GC at lead marker chr20:45906012. Shown are the three individual sub-tests, including single marker test (SMT), stable distillation (SD) and quadratic form (Qform), as well as the three tests combined. Grey bars show -log$_{10}$(P) for SD, QForm and "combined" before modified GC by the slope and intercept of Q-Q plots, black bars show results after modified GC, and the final black bar at right shows the final combined -log$_{10}$(P) used for all final results, which also accounts for the different number of independent tests performed by SMT and LOCATER. **(E)** Local Manhattan plot of "triglycerides in small HDL" on chr20:45306012-46561659, showing modified GCed -log$_{10}$(P) for the 3 LOCATER sub-tests.

#### Figure S10F

**(F)**

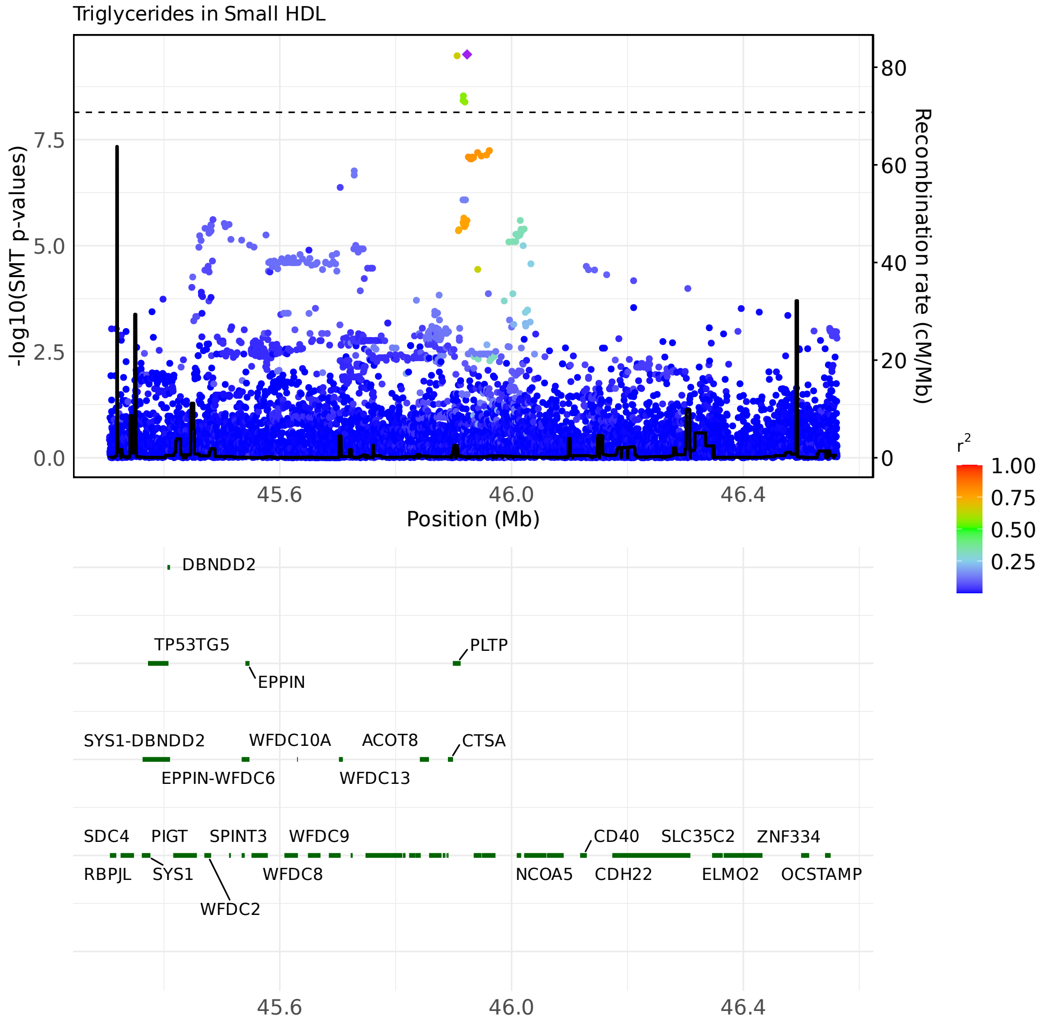


LocusZoom plots of SMT results for triglycerides in small HDL at chr20:44881796-46728887. Variants are colored based on their r² with the focal marker (purple diamond), where LD is calculated in the studied samples. The black line shows the recombination rate in Finns (See Methods). Gene annotations are from GENCODE v45. **(F)** LocusZoom plot based on SMT lead marker chr20:45923216. This variant is also documented to be associated with this exact phenotype.

#### Figure S10G-H

**(G)**

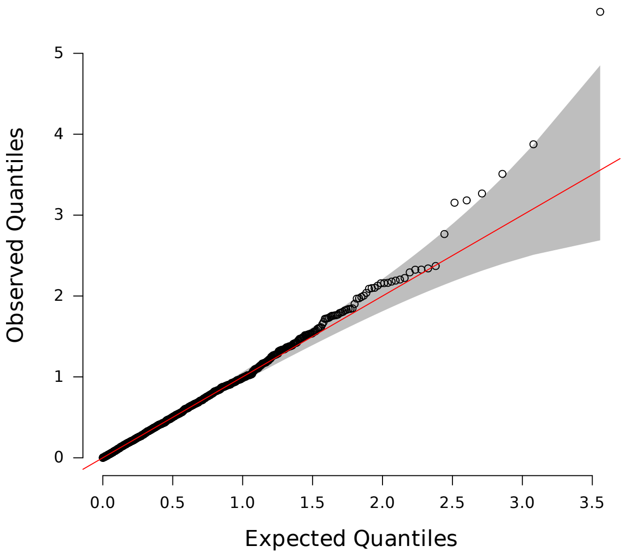


**(G)** Q-Q inflation plot of -log10(p-values) from all “sprigs” at the lead marker chr20:45906012, where “sprigs” are defined as the smallest possible inferred clades. The gray area corresponds to the 95% confidence interval, and the red line denotes x=y. **(H)** Histogram of phenotype values after projecting out the genotype vector of the LOCATER lead marker (chr20:45906012), thus removing signal that can be accounted for by the SMT sub-test. Connected dots show the phenotype value of individuals assigned to significant sprigs.

**(H)**

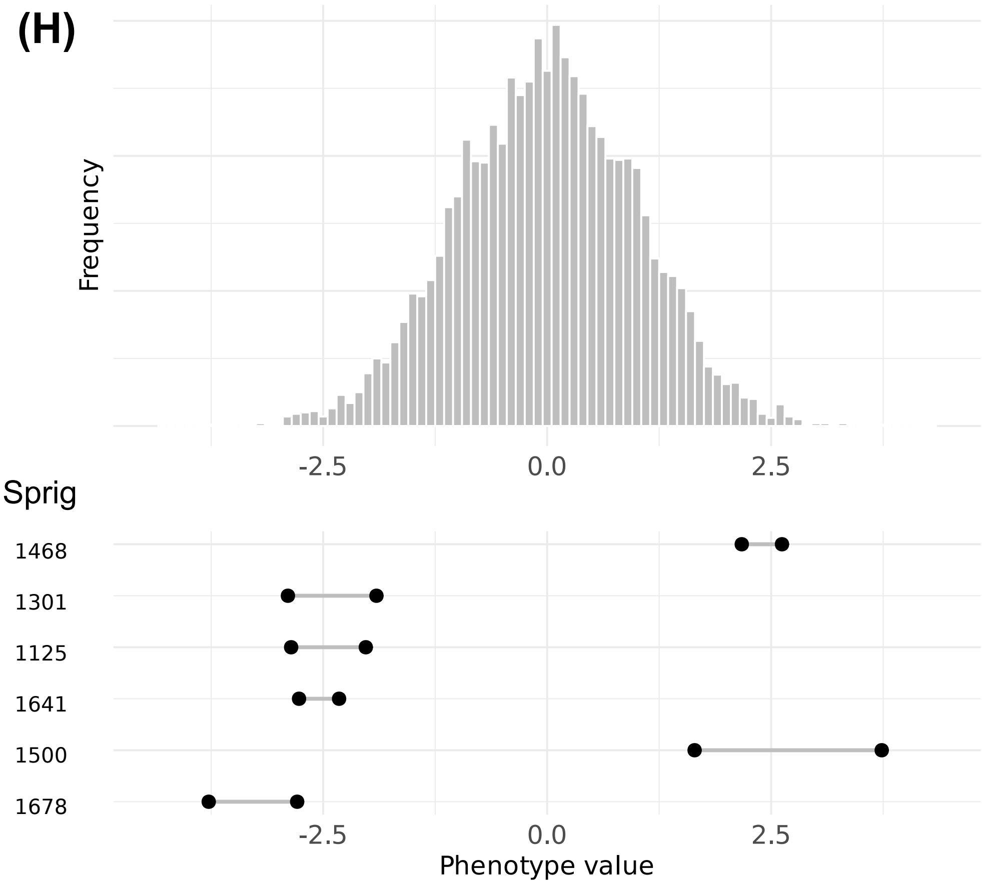

Figure S10I

(I)

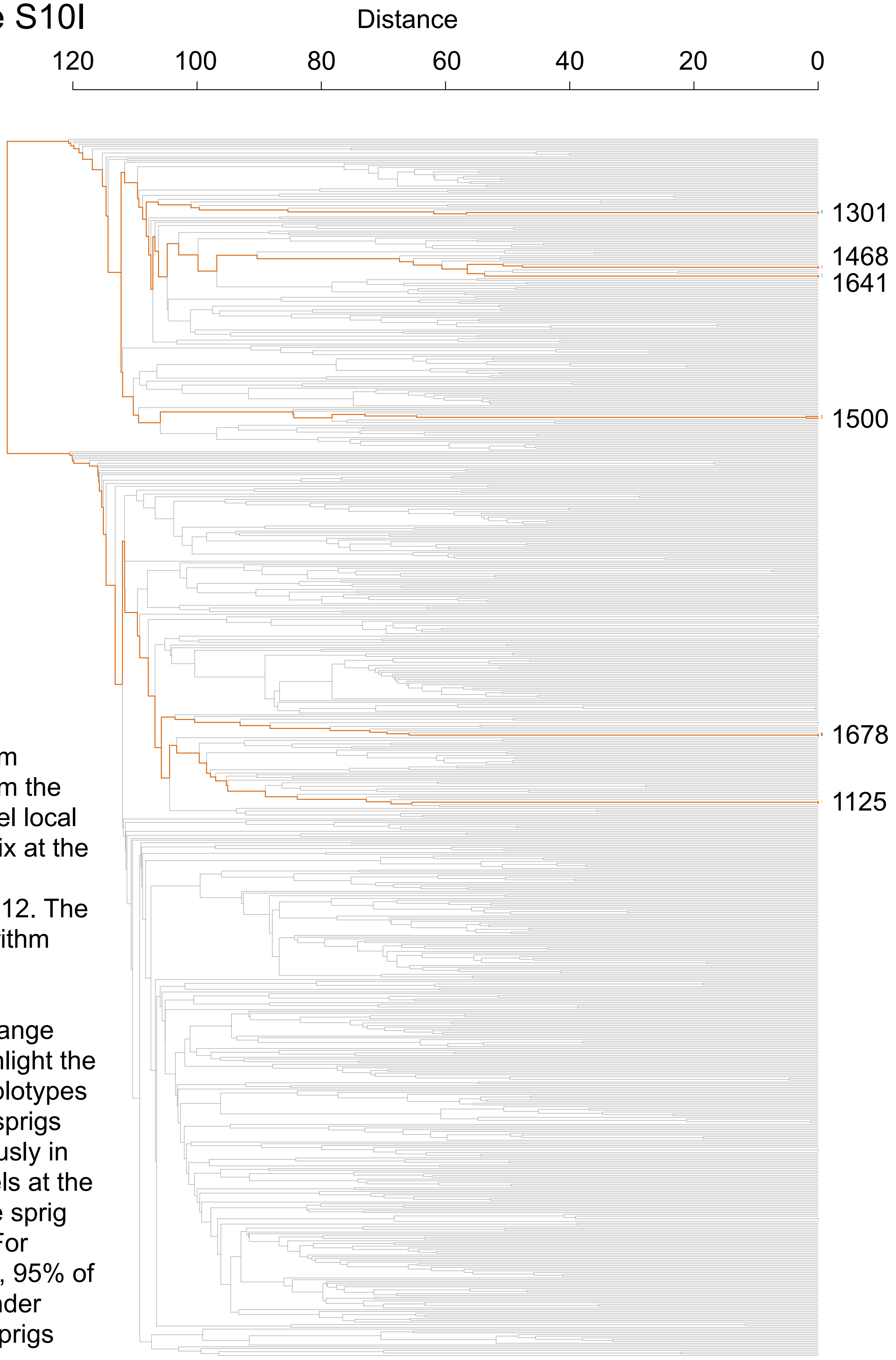


**(I)** Dendrogram generated from the haplotype-level local distance matrix at the lead marker chr20:45906012. The UPGMA algorithm was used for hierarchical clustering. Orange branches highlight the path of all haplotypes in significant sprigs shown previously in part **(H)**. Labels at the right show the sprig assignment. For plotting clarity, 95% of haplotypes under insignificant sprigs were pruned.

### Figure S10J-K

## (J)

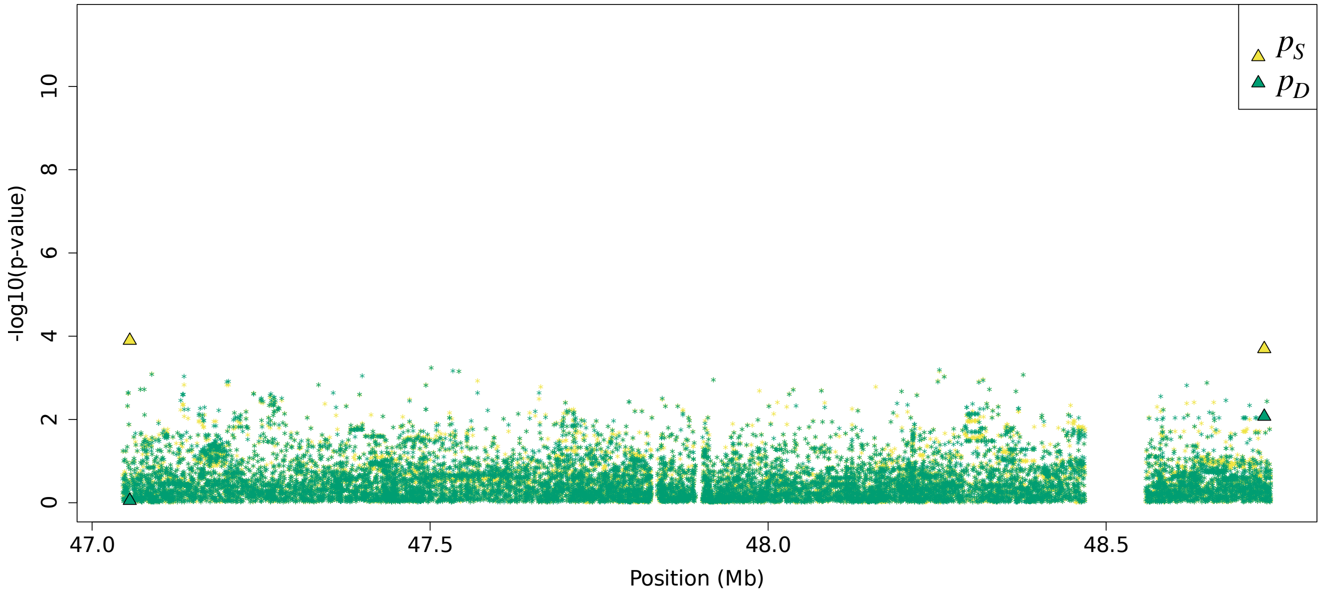


## (K)

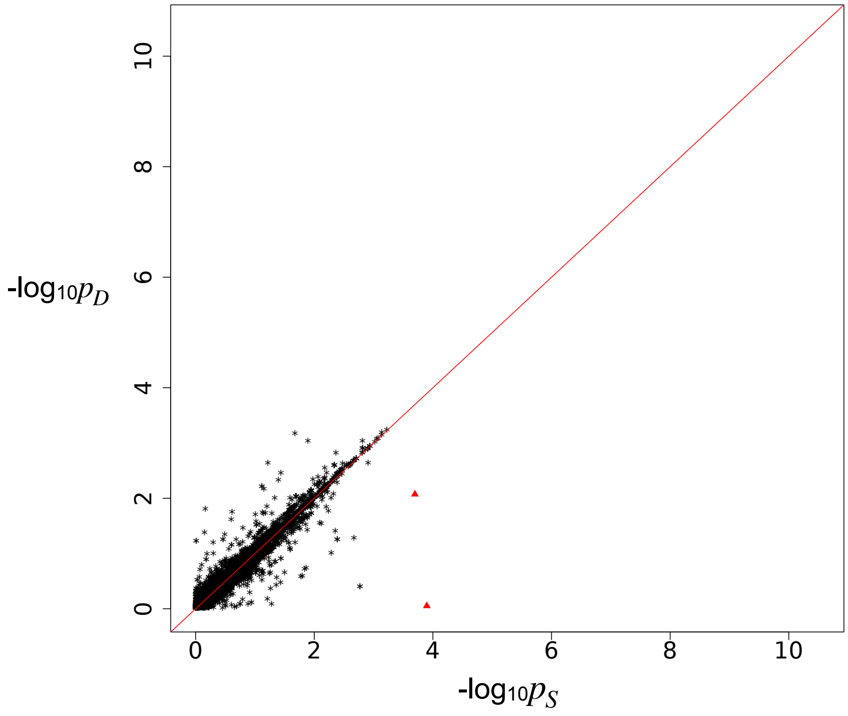


Residual association signals at the *PLTP* locus after accounting for signal from the LOCATER lead marker. $p_S$ is defined as the SMT p-value based on a residualized phenotype orthogonal against the LOCATER lead marker; $p_D$ is the SMT p-value based on a residualized phenotype orthogonal against the LOCATER lead marker and also the SD signal. The difference between $p_S$ and $p_D$ thus shows the contribution of genomic variants to the SD signal. Triangles: genomic variants with $p_S < 10^{-3}$ and $p_D > 10 * p_S$. **(J)** Local Manhattan plot of $p_S$ and $p_D$. **(K)** Scatter plot of $p_S$ and $p_D$.

Figure S10L

**(L)**

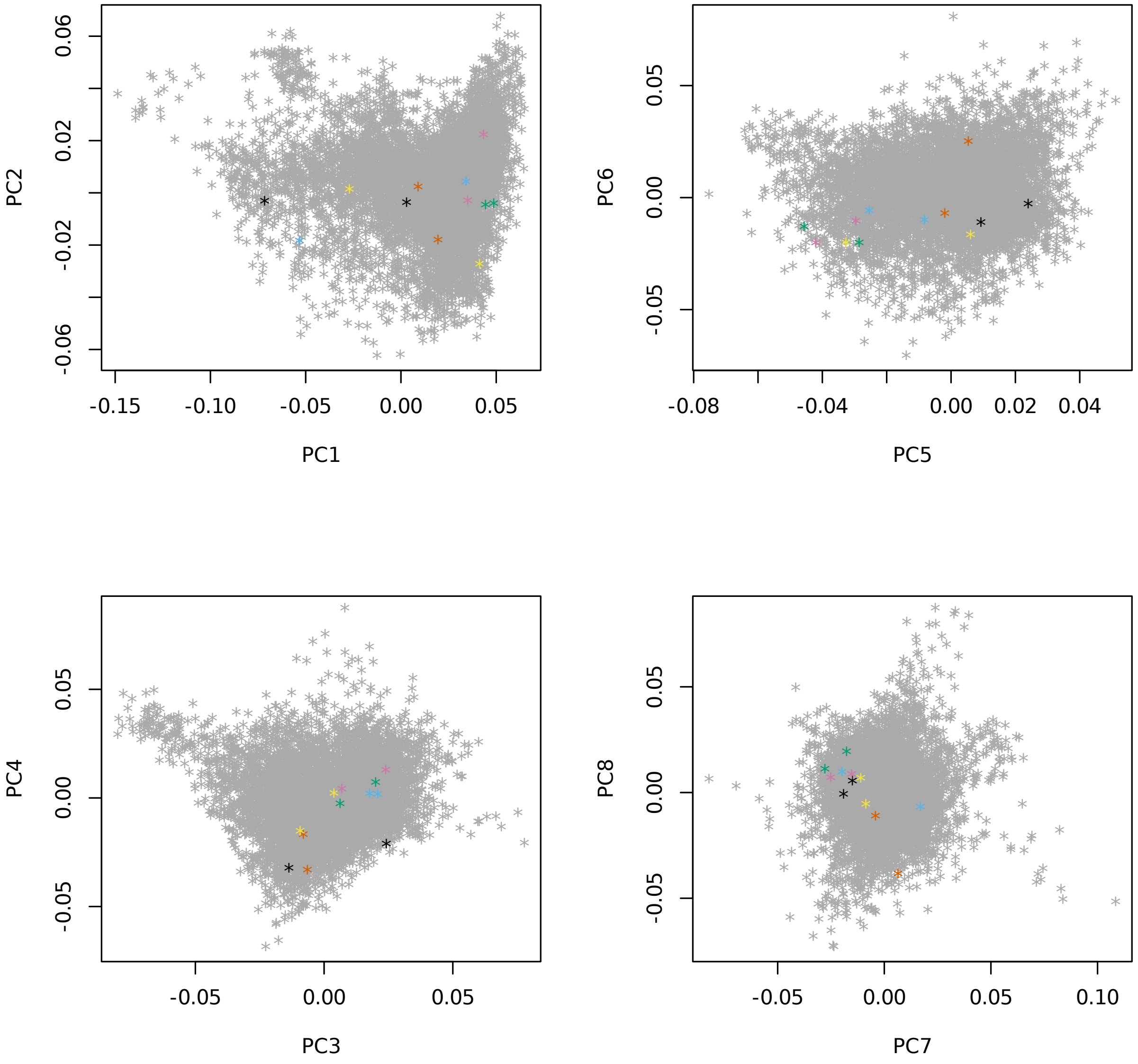


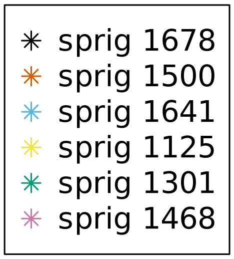


**(L)** Principal components 1-8, highlighting individuals in significant “sprigs”, where “sprigs” are defined as the smallest possible inferred clades. Individuals in the same sprig use the same color and marker.

### Figure S11 Large VLDL particle association on chr7
### Figure S11A-C

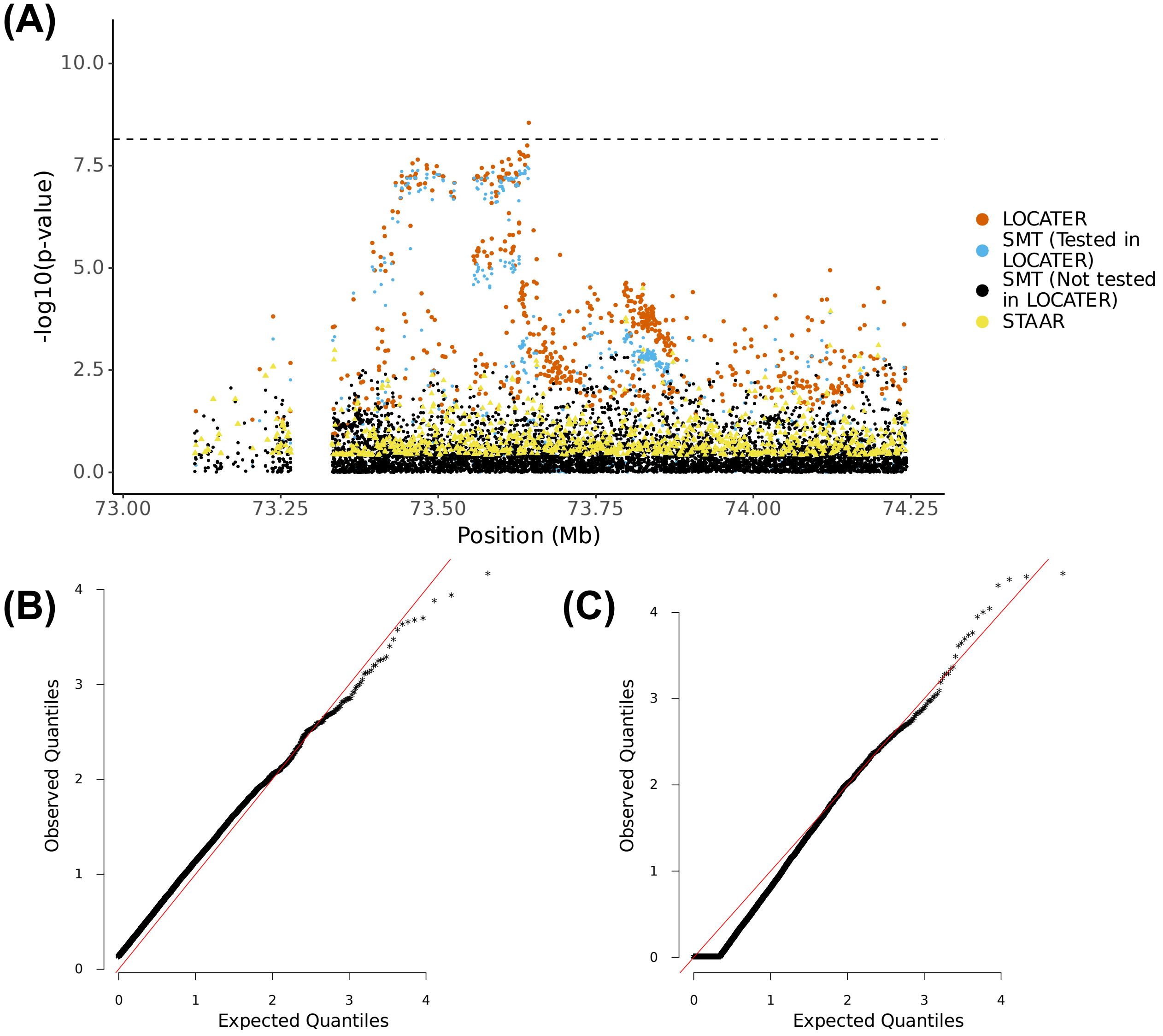


**(A)** Local Manhattan plot of the association signal for "Concentration of large VLDL particles" on chr7:73043687-74243687, including results for single marker test (SMT; blue and black), LOCATER (orange) and STAAR (yellow). Note that LOCATER results are only shown for variants with an SMT p-value less than $1x10^{-3}$, since for computational efficiency only these variants were tested by LOCATER (see Methods). SMT results from variants tested by LOCATER are shown in blue, and those from variants not tested by LOCATER are shown in black. The black dashed line corresponds to the genome-wide significance threshold for SMT, standardized LOCATER, and standardized STAAR. **(B)** and **(C)** are Q-Q inflation plots after modified GC of the LOCATER sub-tests stable distillation (SD) and quadratic form (QForm), respectively, for "Concentration of large VLDL particles". The red line on the diagonal corresponds to x=y.

Figure S11D-E

(D)

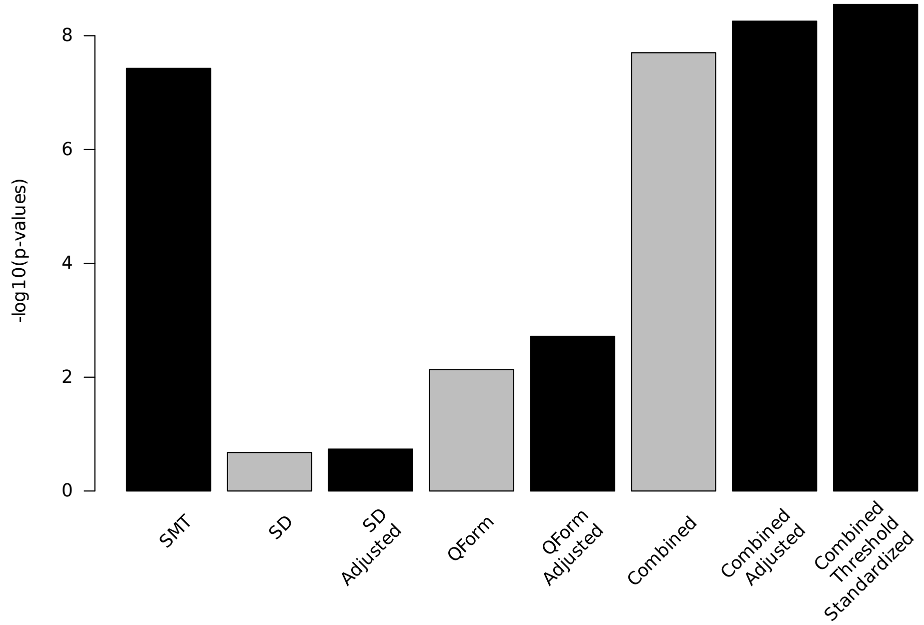


(E)

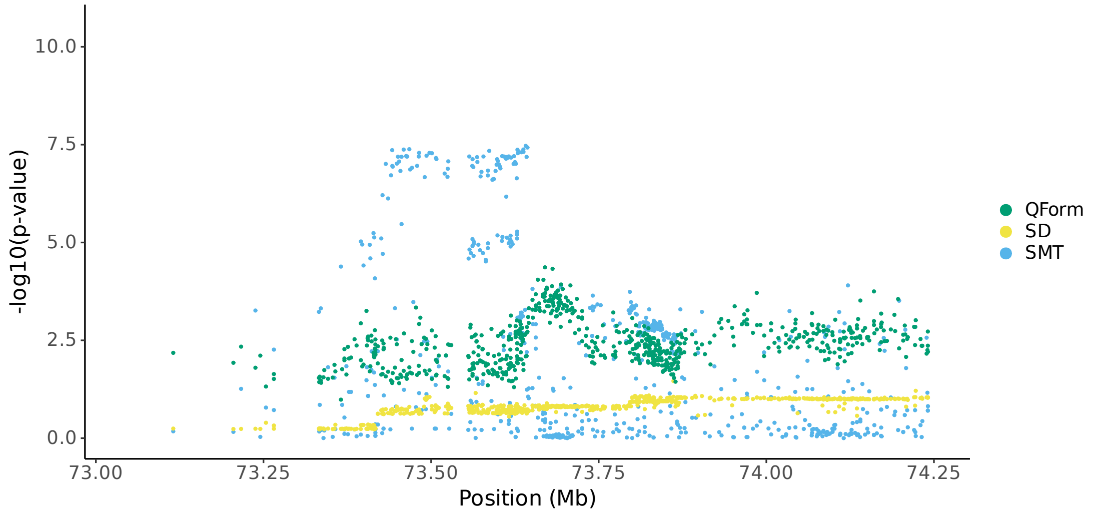


**(D)** Bar plot of -log$_{10}$(P) for association results before and after modified GC at lead marker chr7:73643687. Shown are the three individual sub-tests, including single marker test (SMT), stable distillation (SD) and quadratic form (Qform), as well as the three tests combined. Grey bars show -log$_{10}$(P) for SD, QForm and "combined" before modified GC by the slope and intercept of Q-Q plots, black bars show results after modified GC, and the final black bar at right shows the final combined -log$_{10}$(P) used for all final results, which also accounts for the different number of independent tests performed by SMT and LOCATER. **(E)** Local Manhattan plot of "Concentration of large VLDL particles" on chr7:73043687-74243687, showing modified GCed -log$_{10}$(P) for the 3 LOCATER sub-tests.

#### Figure S11F-H

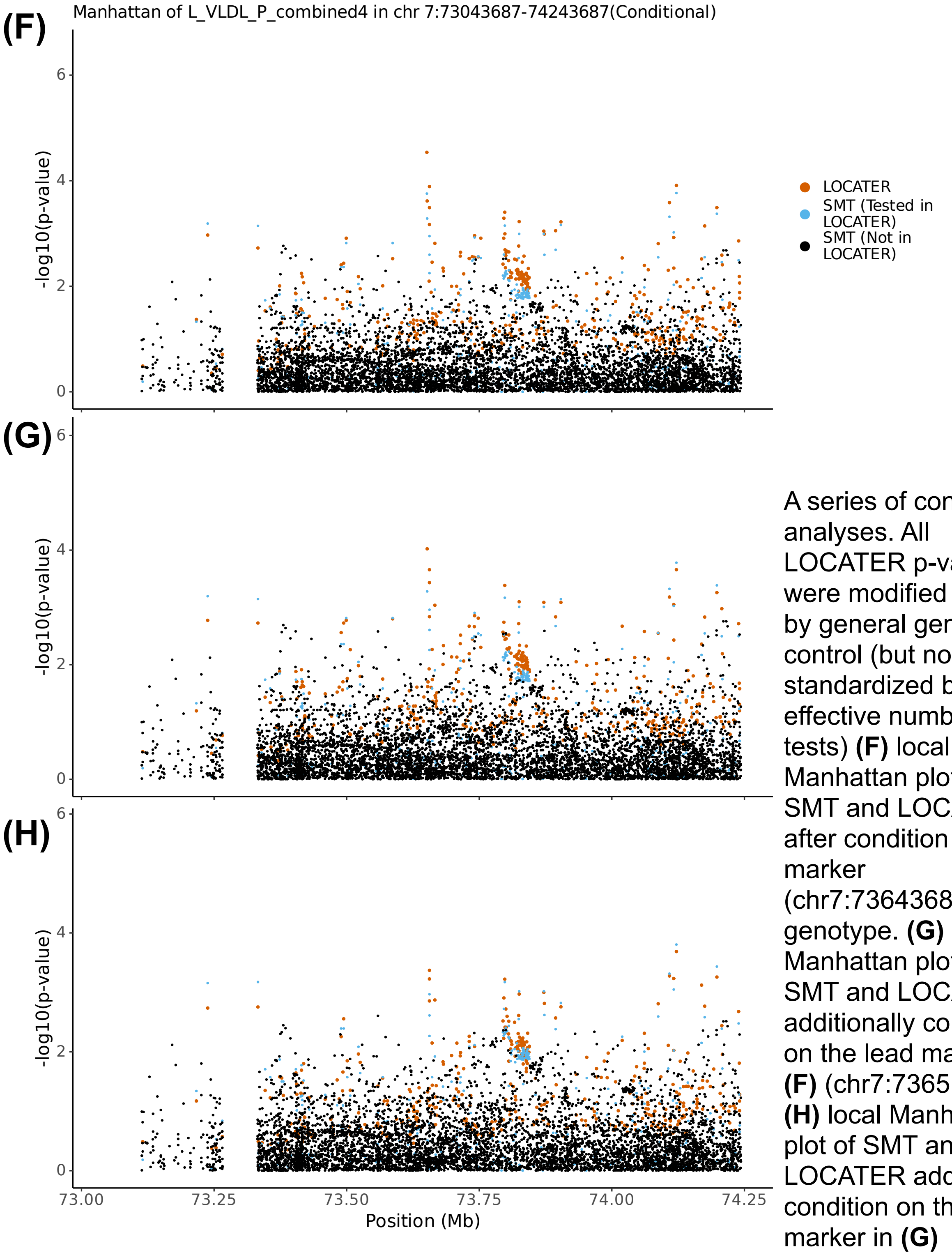


A series of conditional analyses. All LOCATER p-values were modified GCed by general genomic control (but not standardized by the effective number of tests) **(F)** local Manhattan plot of SMT and LOCATER after condition on lead marker (chr7:73643687) genotype. **(G)** local Manhattan plot of SMT and LOCATER additionally condition on the lead marker in **(F)** (chr7:73651197) **(H)** local Manhattan plot of SMT and LOCATER additionally condition on the lead marker in **(G)** (chr7:73651777)

### Figure S11I-J

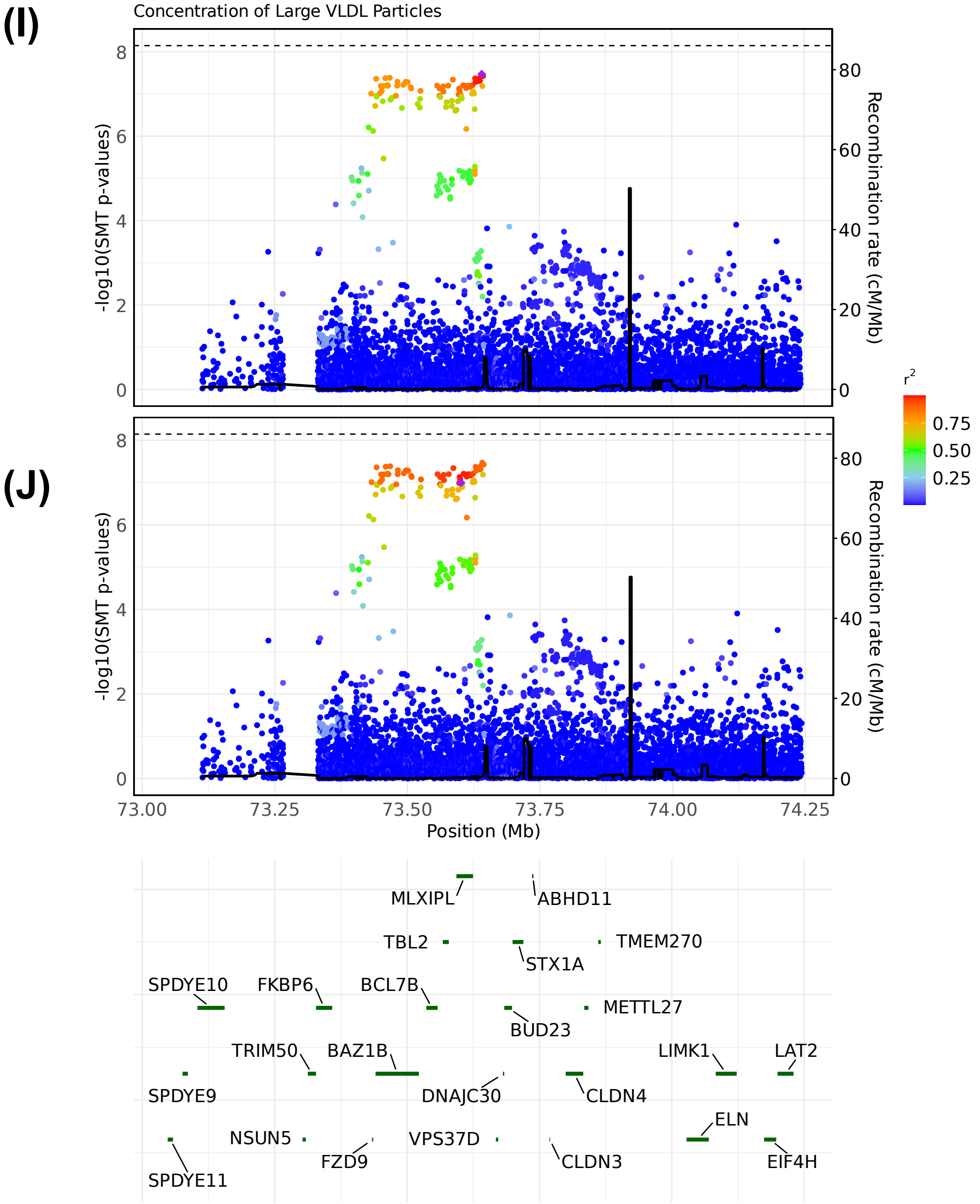


LocusZoom plots of SMT results for “Concentration of large VLDL particles” on chr7:73043687-74243687. Variants are colored based on their $r^2$ with the focal marker (purple diamond), where LD is calculated in the studied samples. The black line shows the recombination rate in Finns (See Methods). Gene annotations are from GENCODE v45. **(I)** LocusZoom plot based on SMT lead marker chr7:73641131. **(J)** LocusZoom plot based on GWAS catalog lead marker chr7:73601039

### Figure S12 Triglycerides in VLDL on chr7
### Figure S12 A-C

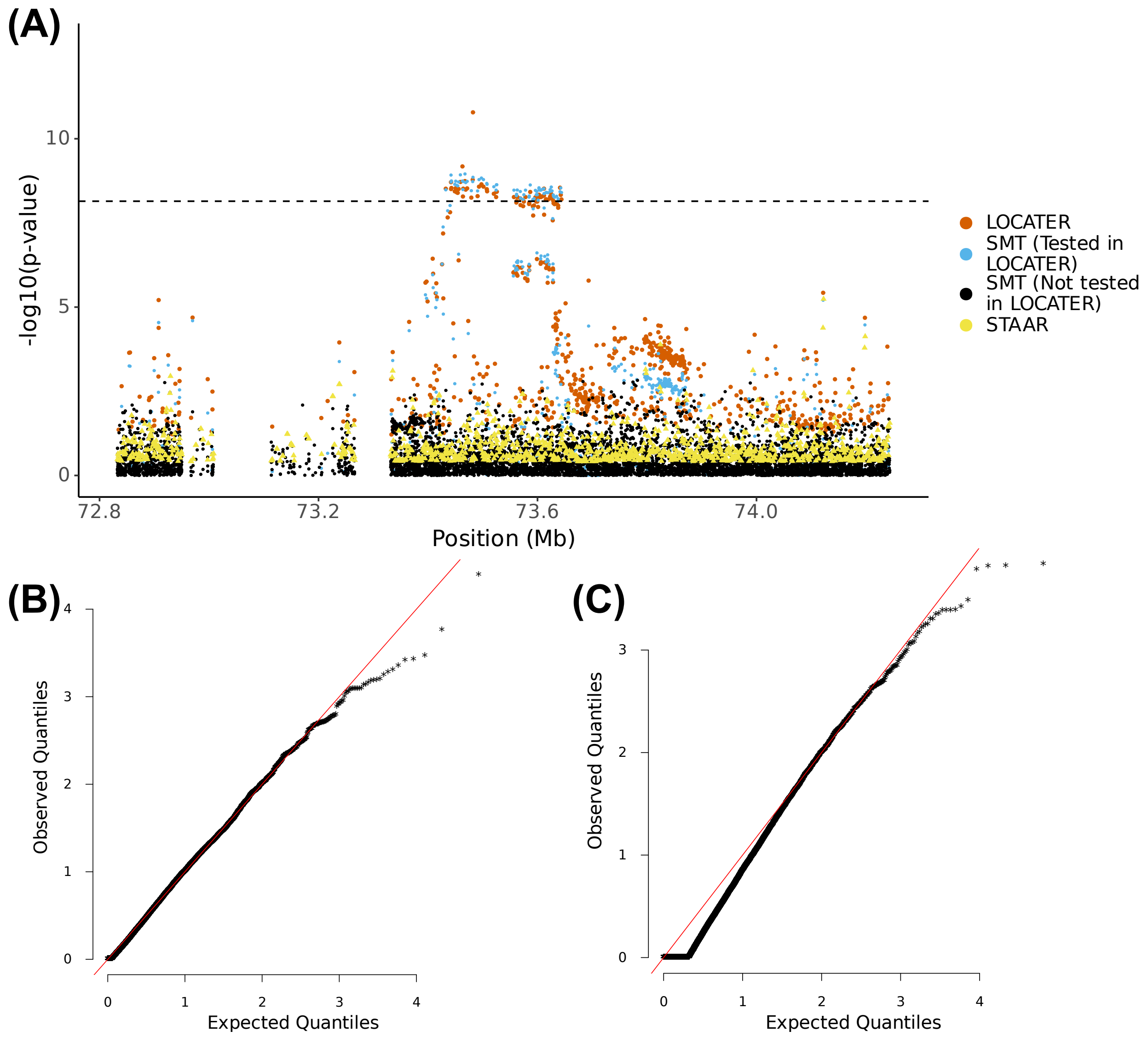


**(A)** Local Manhattan plot of the association signal for “triglycerides in VLDL” on chr7:72832414-74243687, including results for single marker test (SMT; blue and black), LOCATER (orange) and STAAR (yellow). Note that LOCATER results are only shown for variants with an SMT p-value less than $1x10^{-3}$, since for computational efficiency only these variants were tested by LOCATER (see Methods). SMT results from variants tested by LOCATER are shown in blue, and those from variants not tested by LOCATER are shown in black. The black dashed line corresponds to the genome-wide significance threshold for SMT, standardized LOCATER, and standardized STAAR. **(B)** and **(C)** are Q-Q inflation plots after modified GC of the LOCATER sub-tests stable distillation (SD) and quadratic form (QForm), respectively, for “triglycerides in VLDL”. The red line on the diagonal corresponds to x=y.

Figure S12D-E

**(D)**

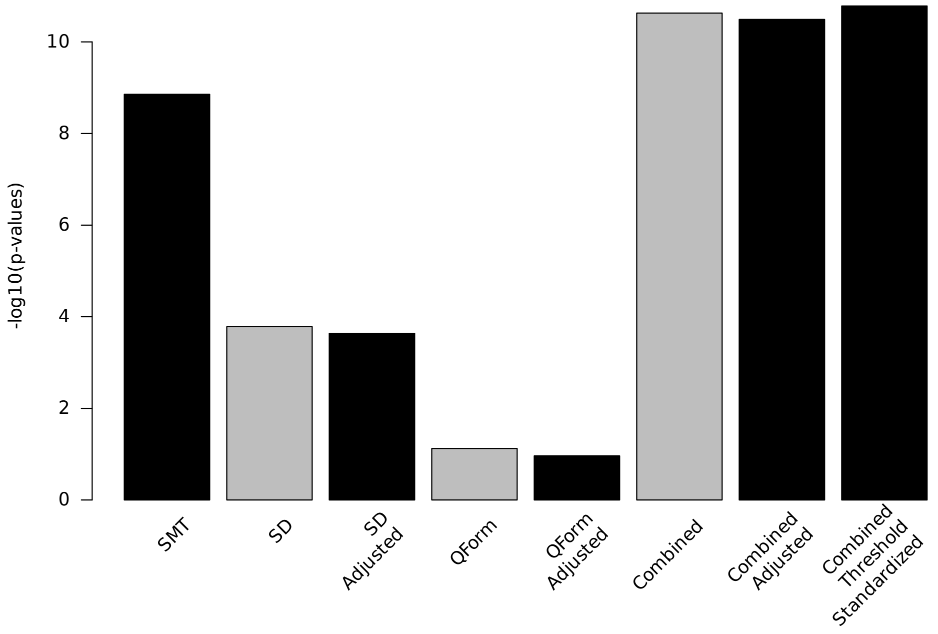


**(E)**

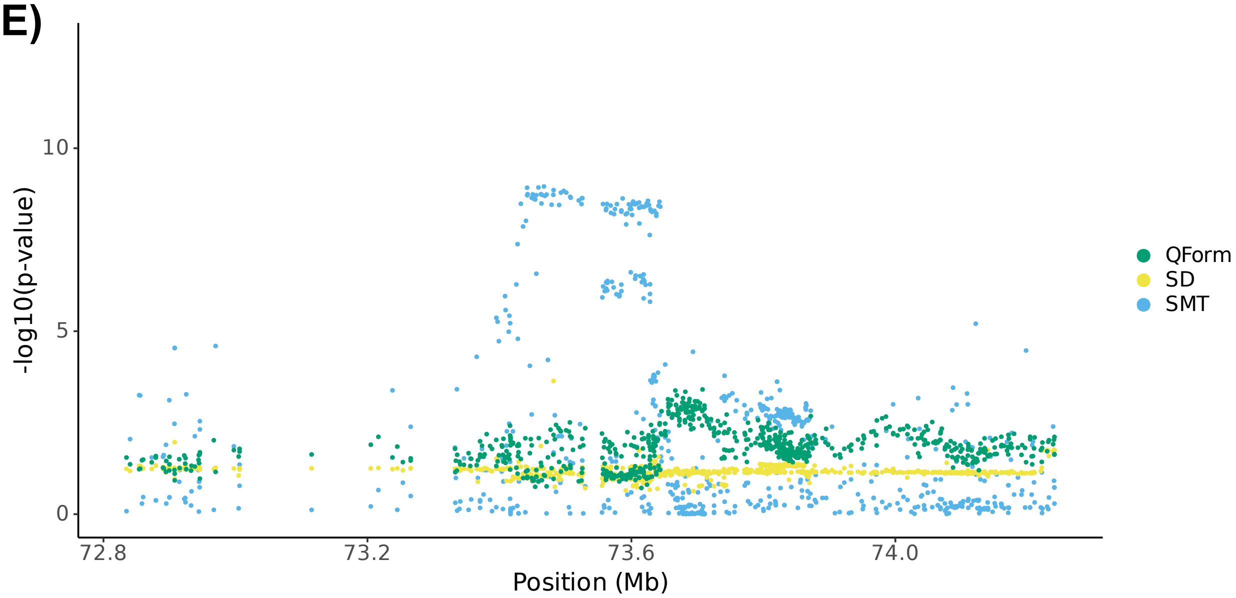


**(D)** Bar plot of -log$_{10}$(P) for association results before and after modified GC at lead marker chr7:73482065. Shown are the three individual sub-tests, including single marker test (SMT), stable distillation (SD) and quadratic form (Qform), as well as the three tests combined. Grey bars show -log$_{10}$(P) for SD, QForm and “combined” before modified GC by the slope and intercept of Q-Q plots, black bars show results after modified GC, and the final black bar at right shows the final combined -log$_{10}$(P) used for all final results, which also accounts for the different number of independent tests performed by SMT and LOCATER. **(E)** Local Manhattan plot of “triglycerides in VLDL” on chr7:72832414-74243687, showing modified GCed -log$_{10}$(P) for the 3 LOCATER sub-tests.

Figure S12F-G

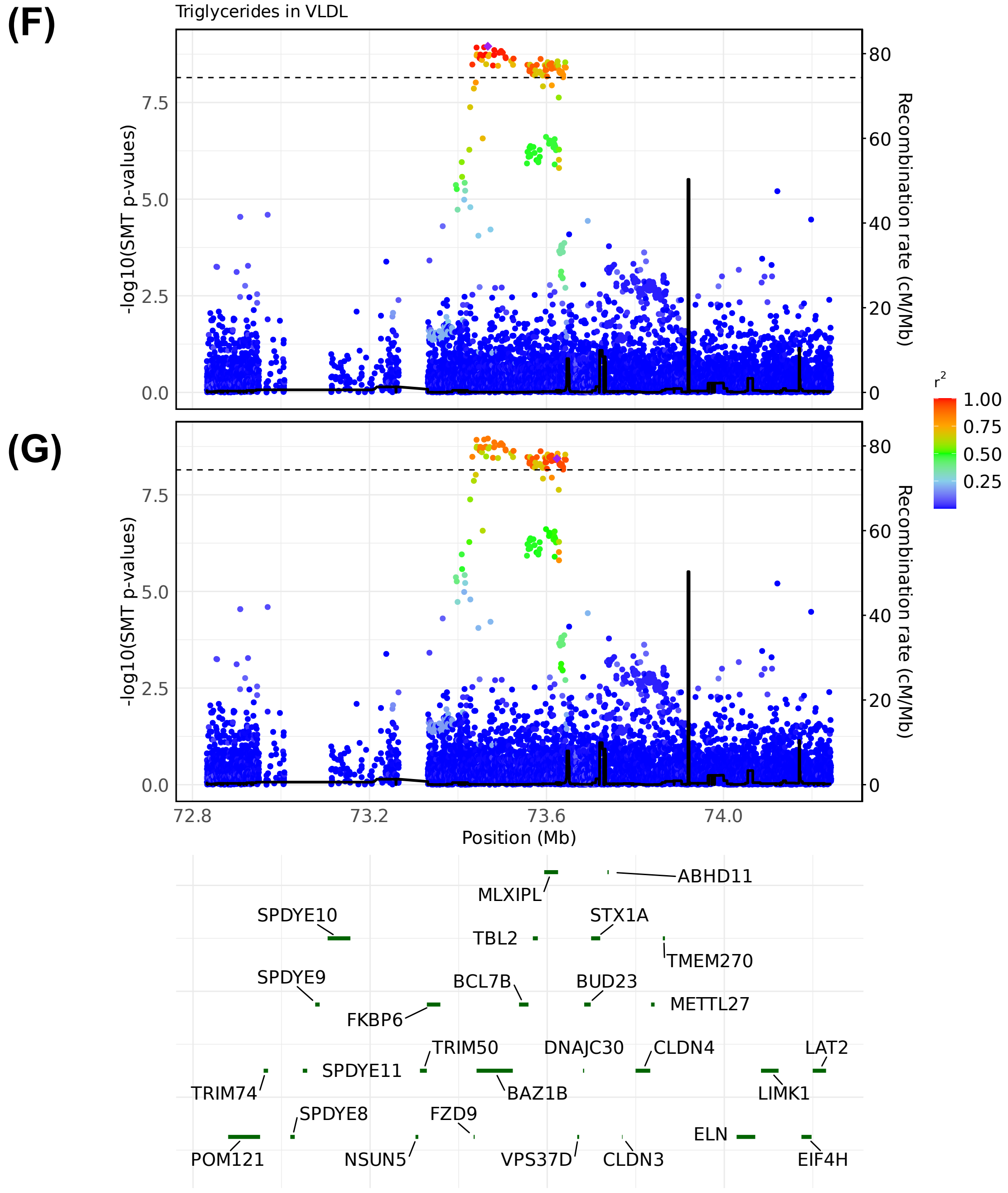


LocusZoom plots of SMT results for “triglycerides in VLDL” on chr7:72832414-74243687. Variants are colored based on their r² with the focal marker (purple diamond), where LD is calculated in the studied samples. The black line shows the recombination rate in Finns (See Methods). Gene annotations are from GENCODE v45. **(F)** LocusZoom plot based on SMT lead marker chr7:73467477. **(G)** LocusZoom plot based on GWAS catalog lead marker chr7:73623626

### Figure S12H-I

**(H)**

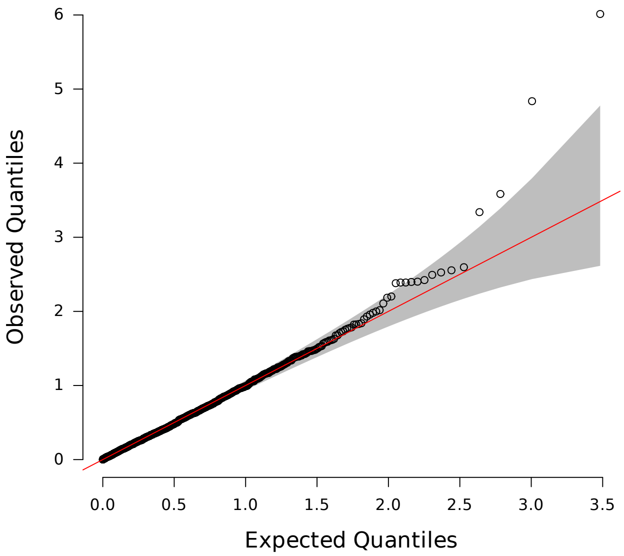


**(H)** Q-Q inflation plot of -log10(p-values) from all "sprigs" at the lead marker chr7:73482065, where "sprigs" are defined as the smallest possible inferred clades. The gray area corresponds to the 95% confidence interval, and the red line denotes x=y. **(I)** Histogram of phenotype values after projecting out the genotype vector of the LOCATER lead marker (chr7:73482065), thus removing signal that can be accounted for by the SMT sub-test. Connected dots show the phenotype value of individuals assigned to significant sprigs.

**(I)**

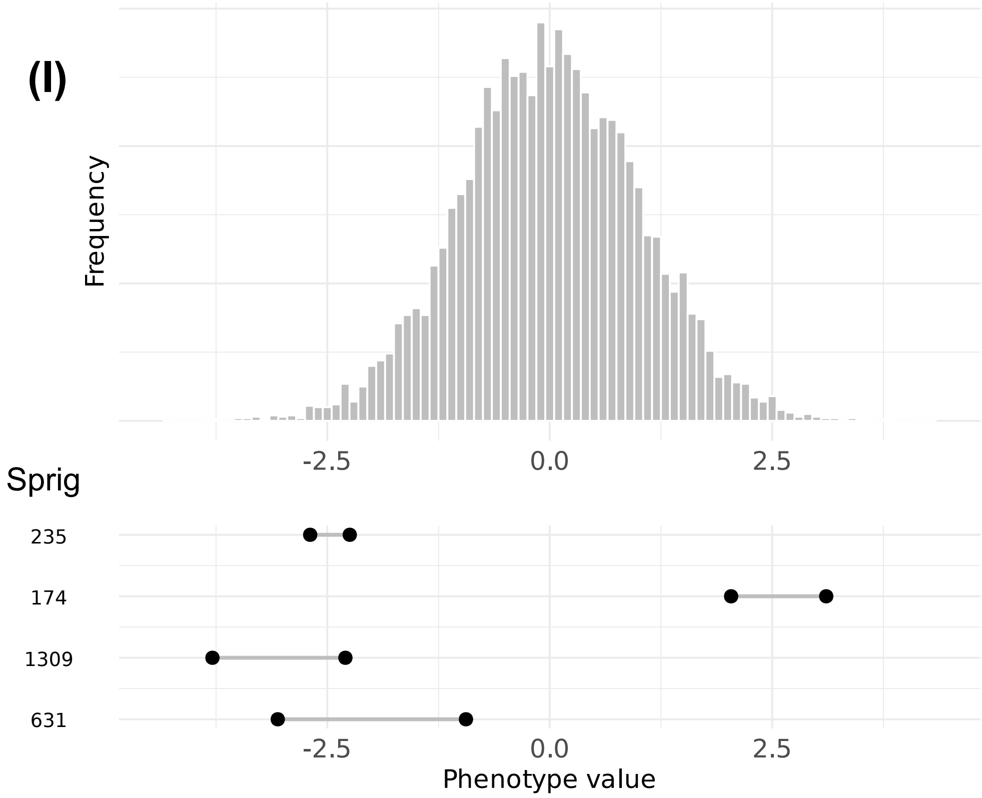

#### Figure S12J

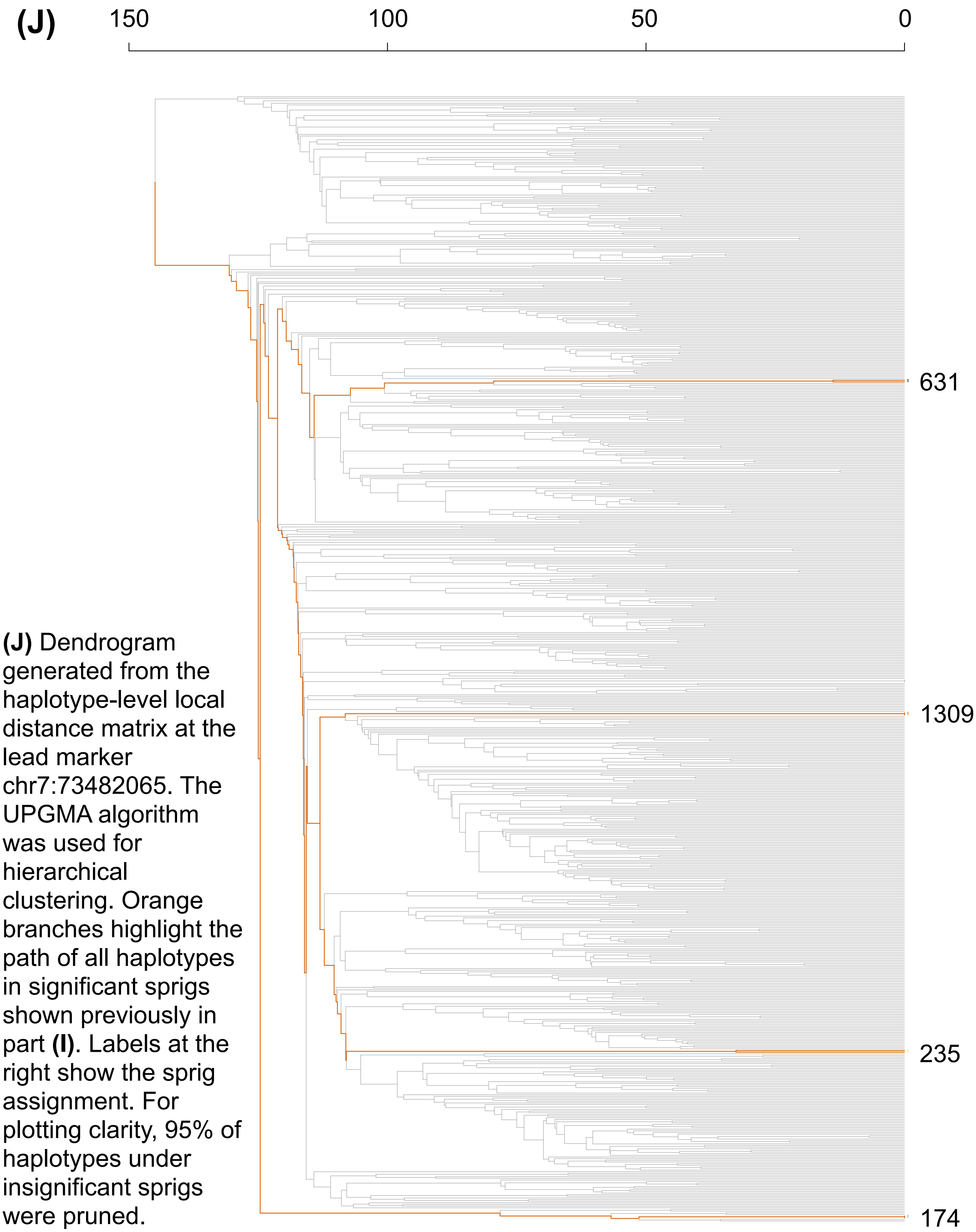


**(J)** Dendrogram generated from the haplotype-level local distance matrix at the lead marker chr7:73482065. The UPGMA algorithm was used for hierarchical clustering. Orange branches highlight the path of all haplotypes in significant sprigs shown previously in part **(I)**. Labels at the right show the sprig assignment. For plotting clarity, 95% of haplotypes under insignificant sprigs were pruned.

Figure S12K-L

**(K)**

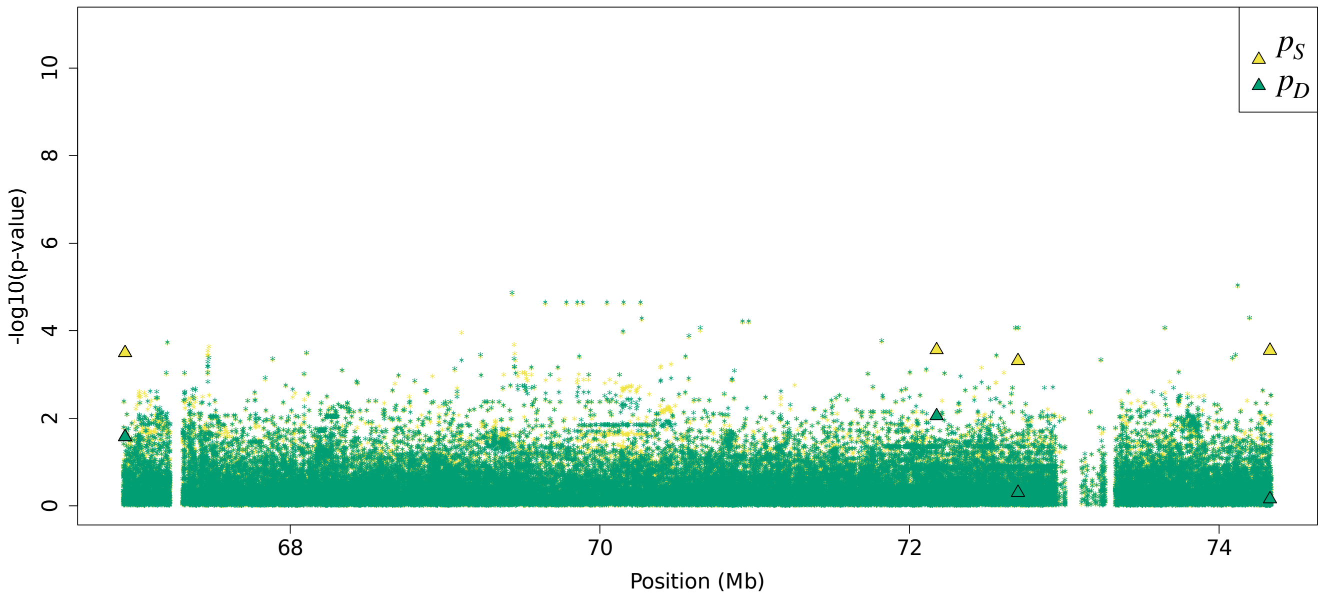


**(L)**

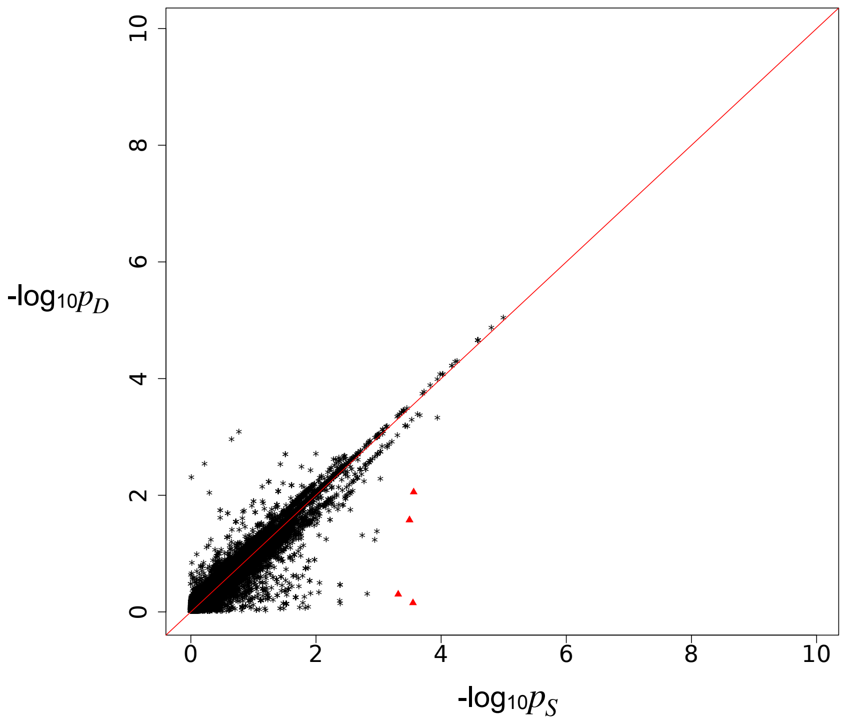


Residual association signals at the *BAZ1B* locus after accounting for signal from the LOCATER lead marker. $p_S$ is defined as the SMT p-value based on a residualized phenotype orthogonal against the LOCATER lead marker; $p_D$ is the SMT p-value based on a residualized phenotype orthogonal against the LOCATER lead marker and also the SD signal. The difference between $p_S$ and $p_D$ thus shows the contribution of genomic variants to the SD signal. Triangles: genomic variants with $p_S < 10^{-3}$ and $p_D > 10 * p_S$. **(K)** Local Manhattan plot of $p_S$ and $p_D$. **(L)** Scatter plot of $p_S$ and $p_D$.

#### Figure S12M

**(M)**

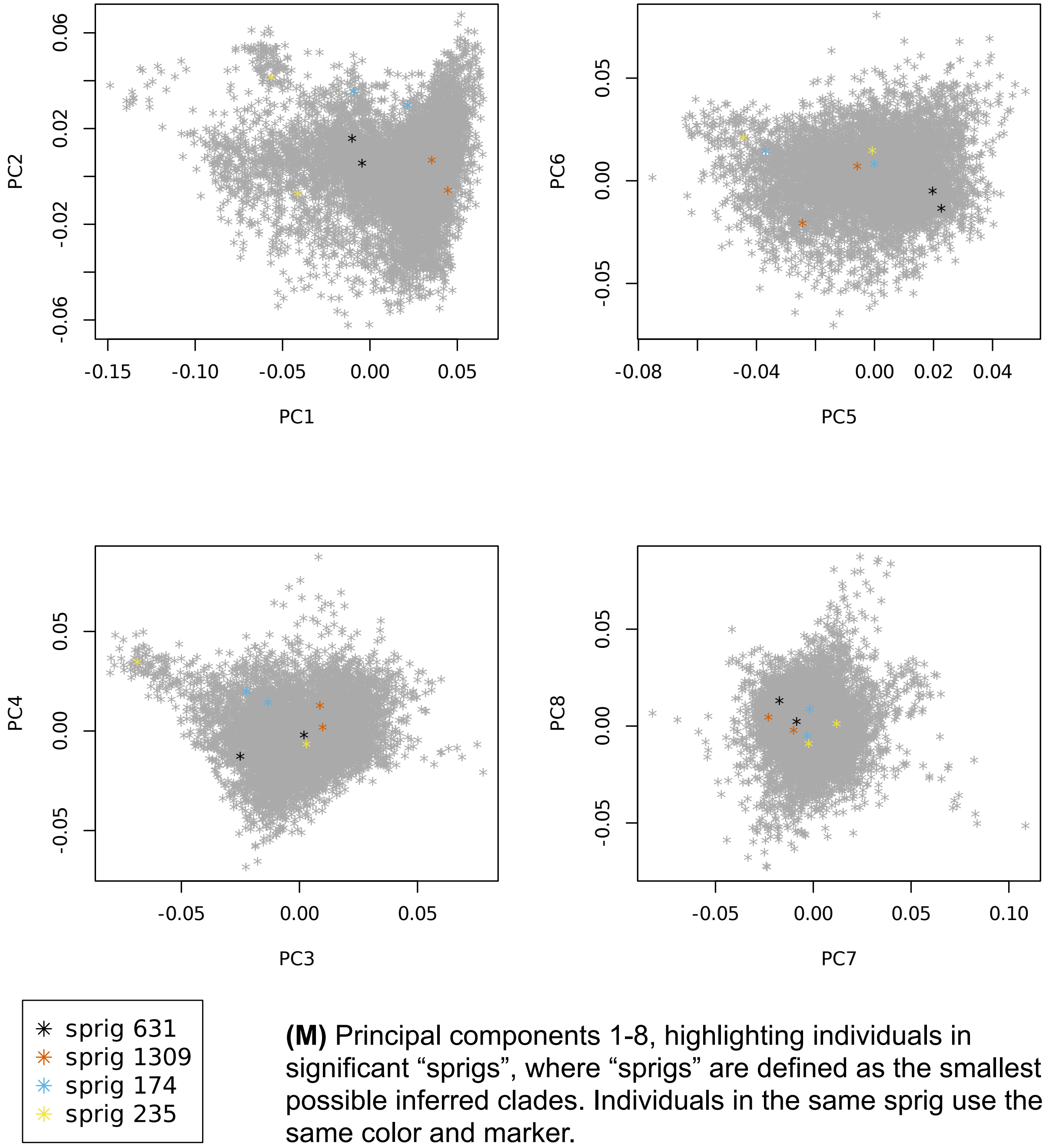


**(M)** Principal components 1-8, highlighting individuals in significant "sprigs", where "sprigs" are defined as the smallest possible inferred clades. Individuals in the same sprig use the same color and marker.

### Figure S13 HDL2-C at *FADS2* locus
### Figure S13A-C

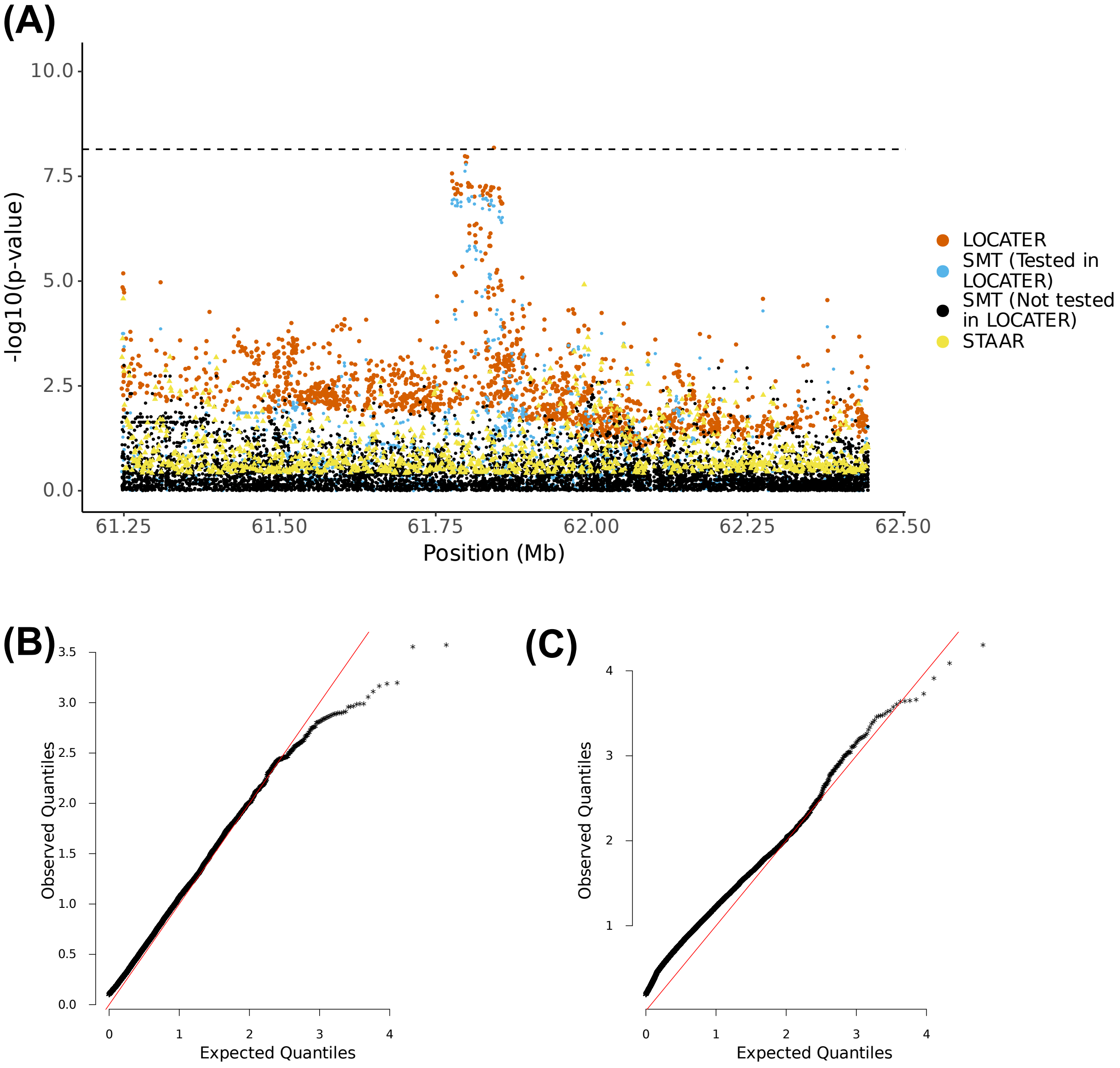


**(A)** Local Manhattan plot of the association signal for HDL2 cholesterol on chr11:61243278-62443278, including results for single marker test (SMT; blue and black), LOCATER (orange) and STAAR (yellow). Note that LOCATER results are only shown for variants with an SMT p-value less than $1x10^{-3}$, since for computational efficiency only these variants were tested by LOCATER (see Methods). SMT results from variants tested by LOCATER are shown in blue, and those from variants not tested by LOCATER are shown in black. The black dashed line corresponds to the genome-wide significance threshold for SMT, standardized LOCATER, and standardized STAAR. **(B)** and **(C)** are Q-Q inflation plots after modified GC of the LOCATER sub-tests stable distillation (SD) and quadratic form (QForm), respectively, for HDL2 cholesterol. The red line on the diagonal corresponds to x=y.

Figure S13D-E

**(D)**

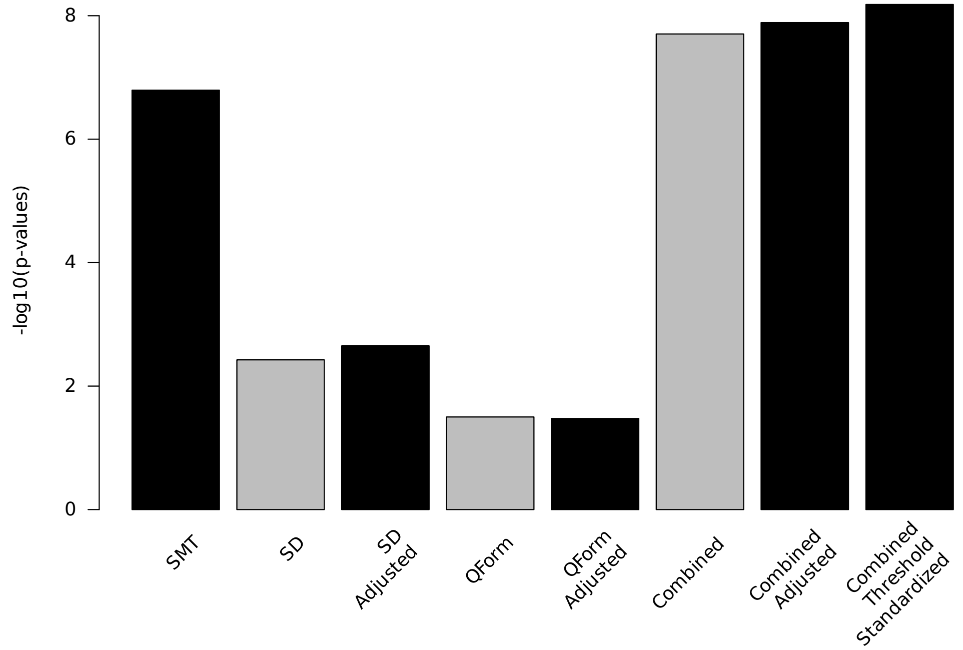


**(E)**

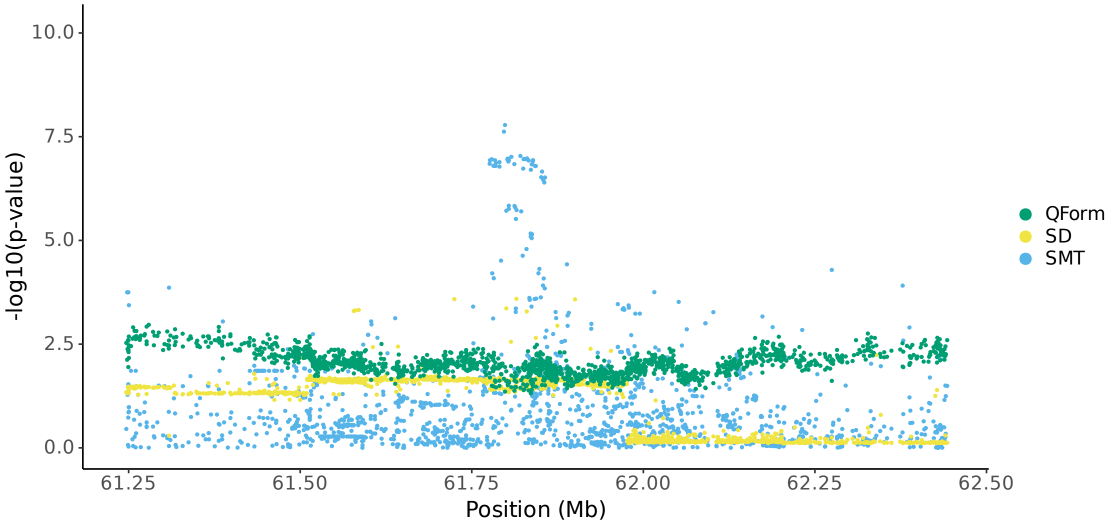


**(D)** Bar plot of -log$_{10}$(P) for association results before and after modified GC at lead marker chr11:61843278. Shown are the three individual sub-tests, including single marker test (SMT), stable distillation (SD) and quadratic form (Qform), as well as the three tests combined. Grey bars show -log$_{10}$(P) for SD, QForm and "combined" before modified GC by the slope and intercept of Q-Q plots, black bars show results after modified GC, and the final black bar at right shows the final combined -log$_{10}$(P) used for all final results, which also accounts for the different number of independent tests performed by SMT and LOCATER. **(E)** Local Manhattan plot of HDL2 cholesterol on chr11:61243278-62443278, showing modified GCed -log$_{10}$(P) for the 3 LOCATER sub-tests.

#### Figure S13F-G

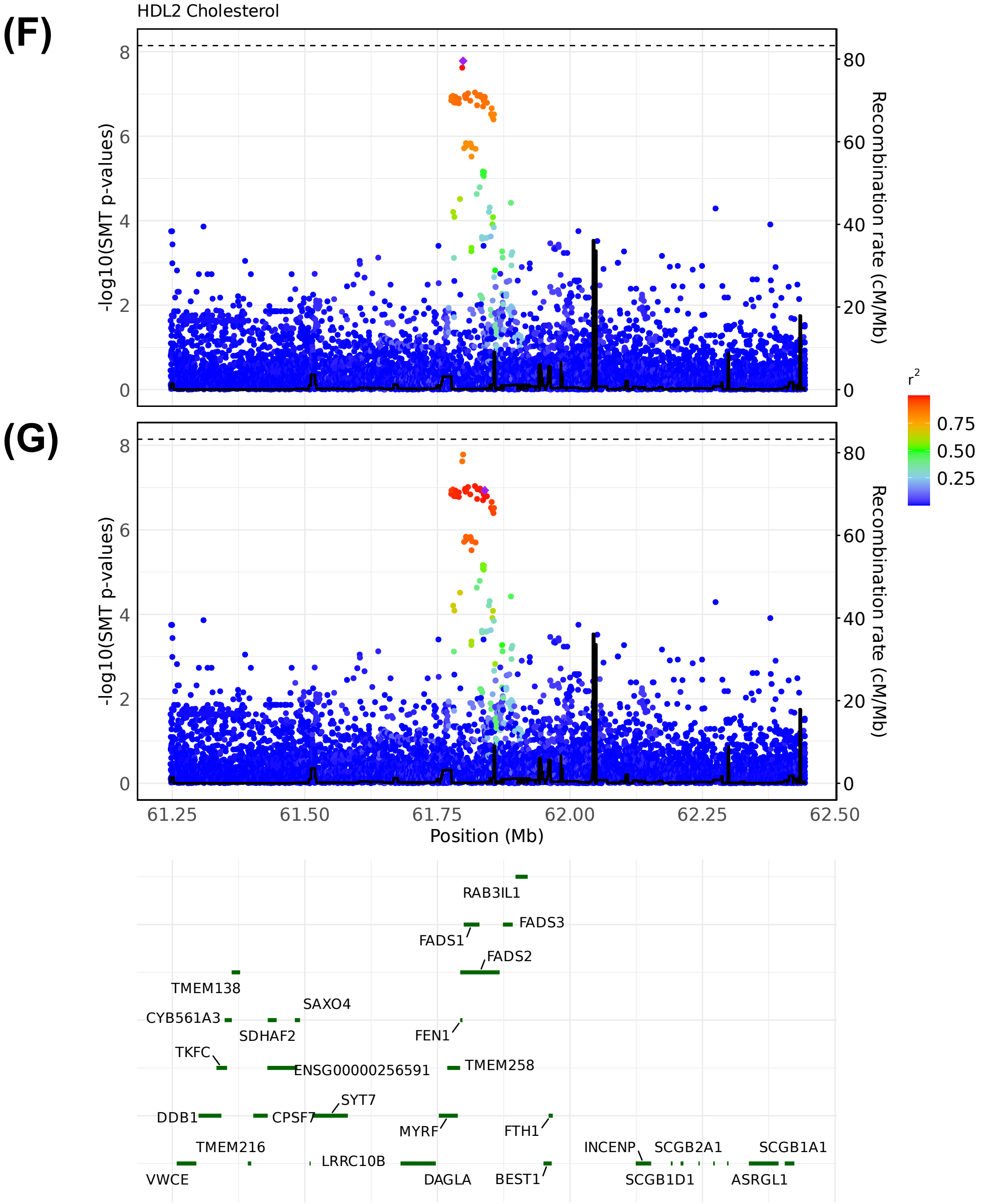


LocusZoom plots of SMT results for HDL2 cholesterol on chr11:61243278-62443278. Variants are colored based on their $r^2$ with the focal marker (purple diamond), where LD is calculated in the studied samples. The black line shows the recombination rate in Finns (See Methods). Gene annotations are from GENCODE v45. **(F)** LocusZoom plot based on SMT lead marker chr11:61798436. **(G)** LocusZoom plot based on GWAS catalog lead marker chr11:61839211.

#### Figure S13H-I

**(H)**

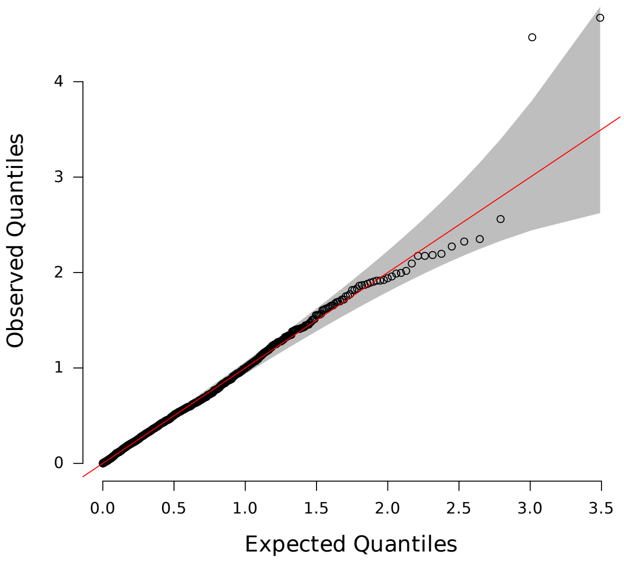


**(H)** Q-Q inflation plot of -log10(p-values) from all "sprigs" at the lead marker chr11:61843278, where "sprigs" are defined as the smallest possible inferred clades. The gray area corresponds to the 95% confidence interval, and the red line denotes x=y. **(I)** Histogram of phenotype values after projecting out the genotype vector of the LOCATER lead marker (chr11:61843278), thus removing signal that can be accounted for by the SMT sub-test. Connected dots show the phenotype value of individuals assigned to significant sprigs.

**(I)**

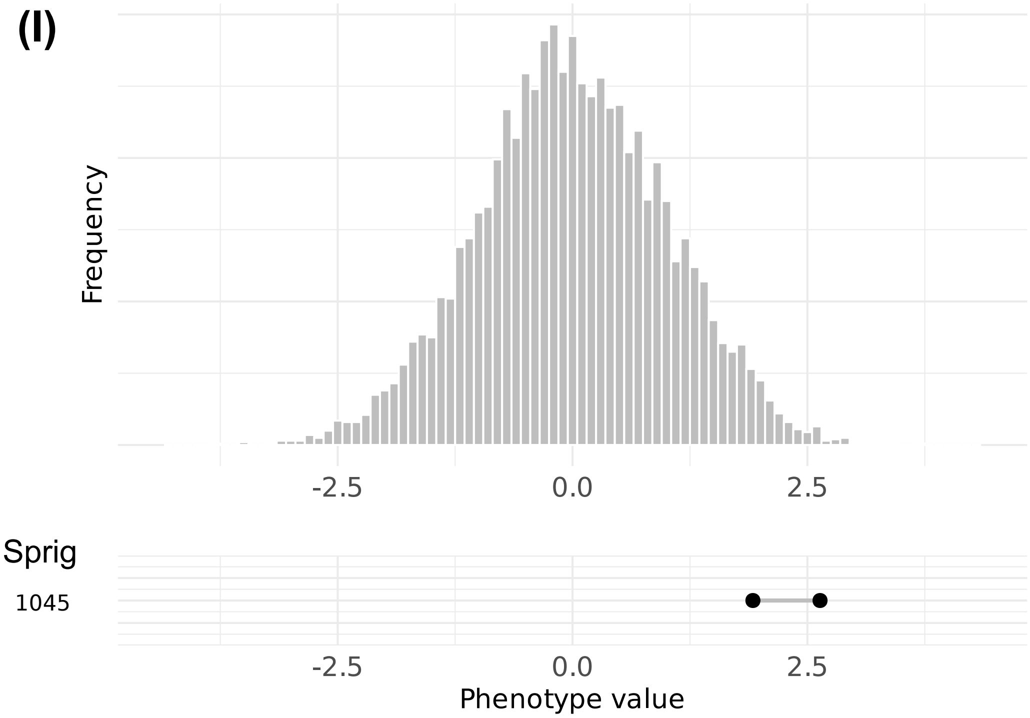

Figure S13J

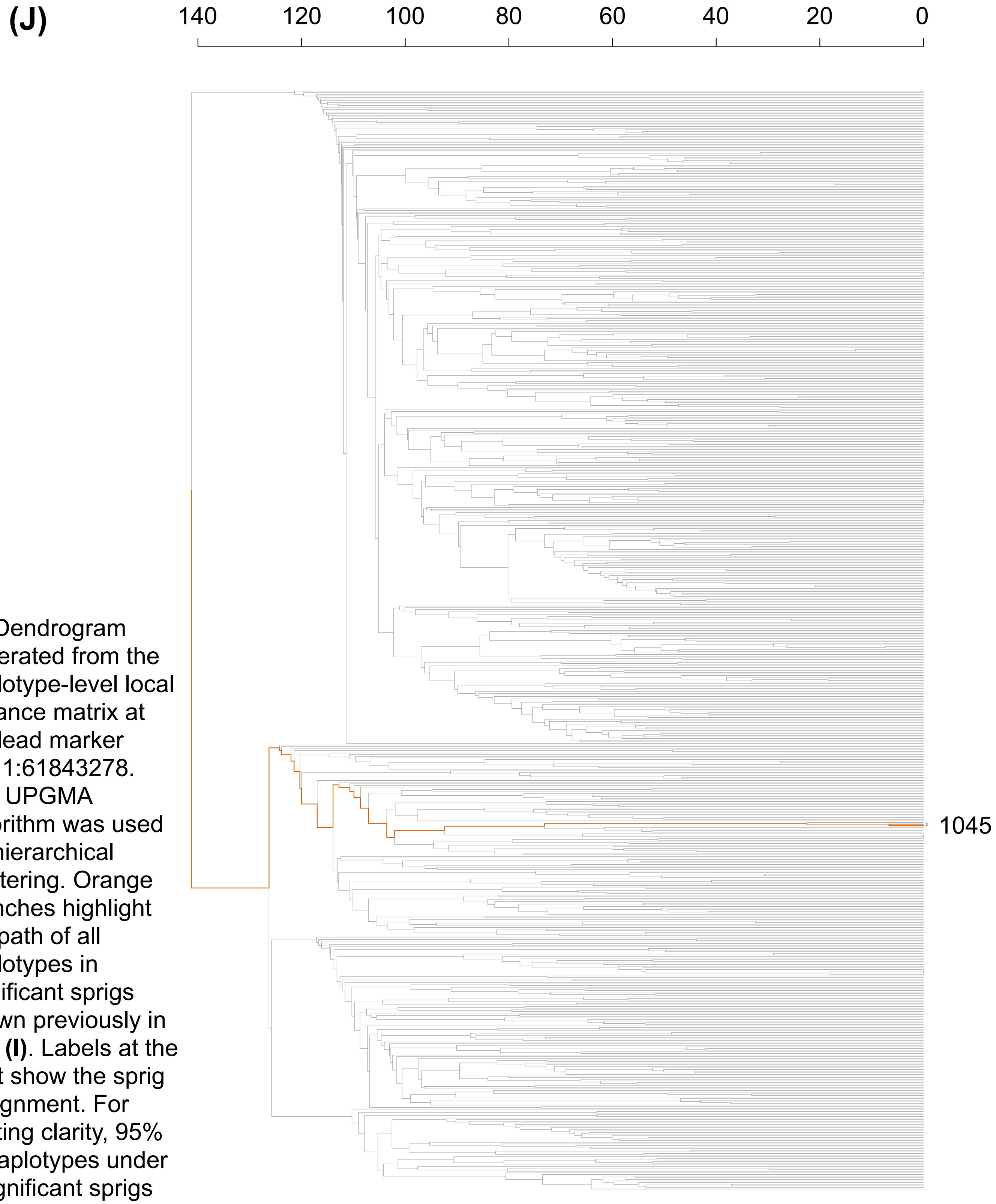


**(J)** Dendrogram generated from the haplotype-level local distance matrix at the lead marker chr11:61843278. The UPGMA algorithm was used for hierarchical clustering. Orange branches highlight the path of all haplotypes in significant sprigs shown previously in part **(I)**. Labels at the right show the sprig assignment. For plotting clarity, 95% of haplotypes under insignificant sprigs were pruned.

#### Figure S13K-L

**(K)**

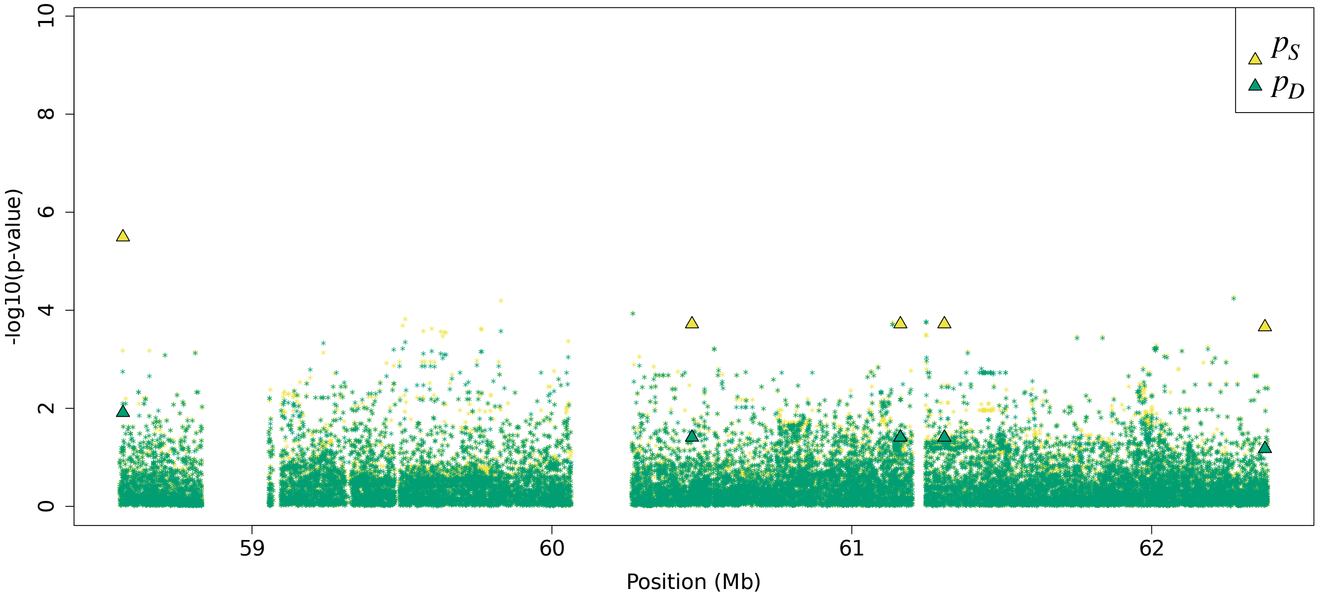


**(L)**

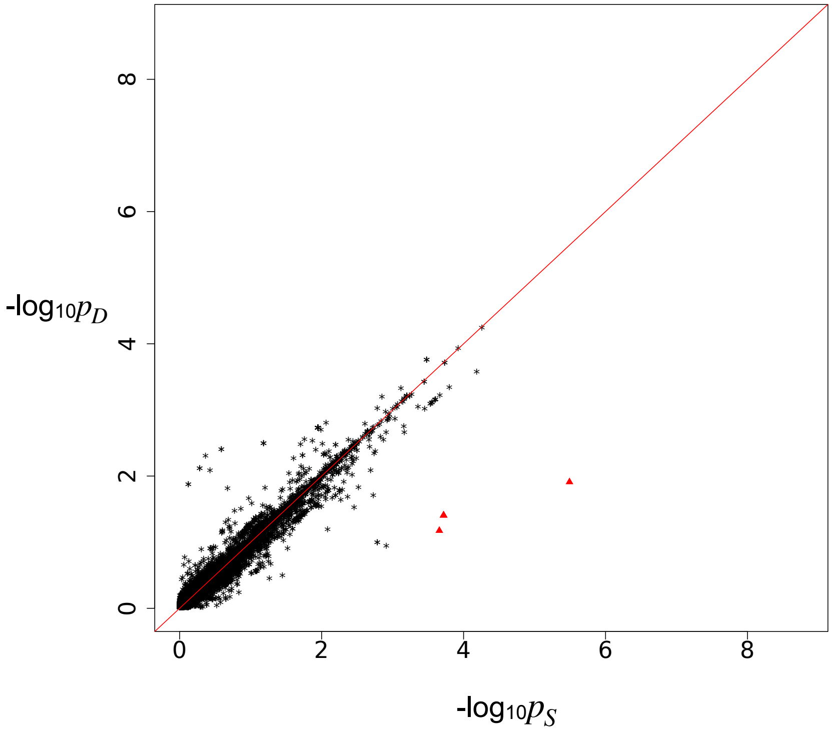


Residual association signals at the *FADS2* locus after accounting for signal from the LOCATER lead marker. $p_S$ is defined as the SMT p-value based on a residualized phenotype orthogonal against the LOCATER lead marker; $p_D$ is the SMT p-value based on a residualized phenotype orthogonal against the LOCATER lead marker and also the SD signal. The difference between $p_S$ and $p_D$ thus shows the contribution of genomic variants to the SD signal. Triangles: genomic variants with $p_S < 10^{-3}$ and $p_D > 10 * p_S$. **(K)** Local Manhattan plot of $p_S$ and $p_D$. **(L)** Scatter plot of $p_S$ and $p_D$.

Figure S13M

**(M)**

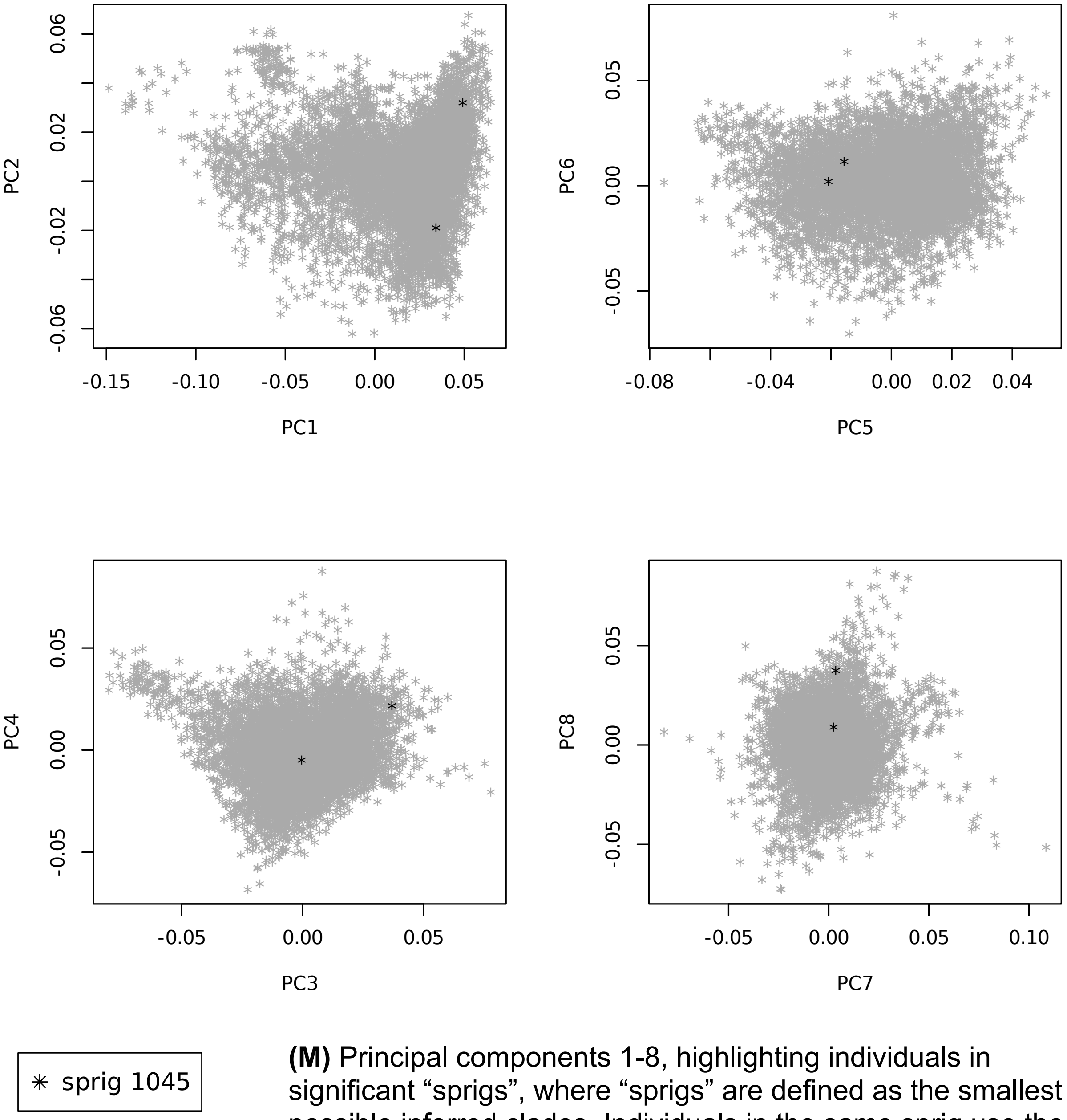


**(M)** Principal components 1-8, highlighting individuals in significant “sprigs”, where “sprigs” are defined as the smallest possible inferred clades. Individuals in the same sprig use the same color and marker.

### Figure S14 HDL3-C at *MIR99AHG*
### Figure S14 A-C

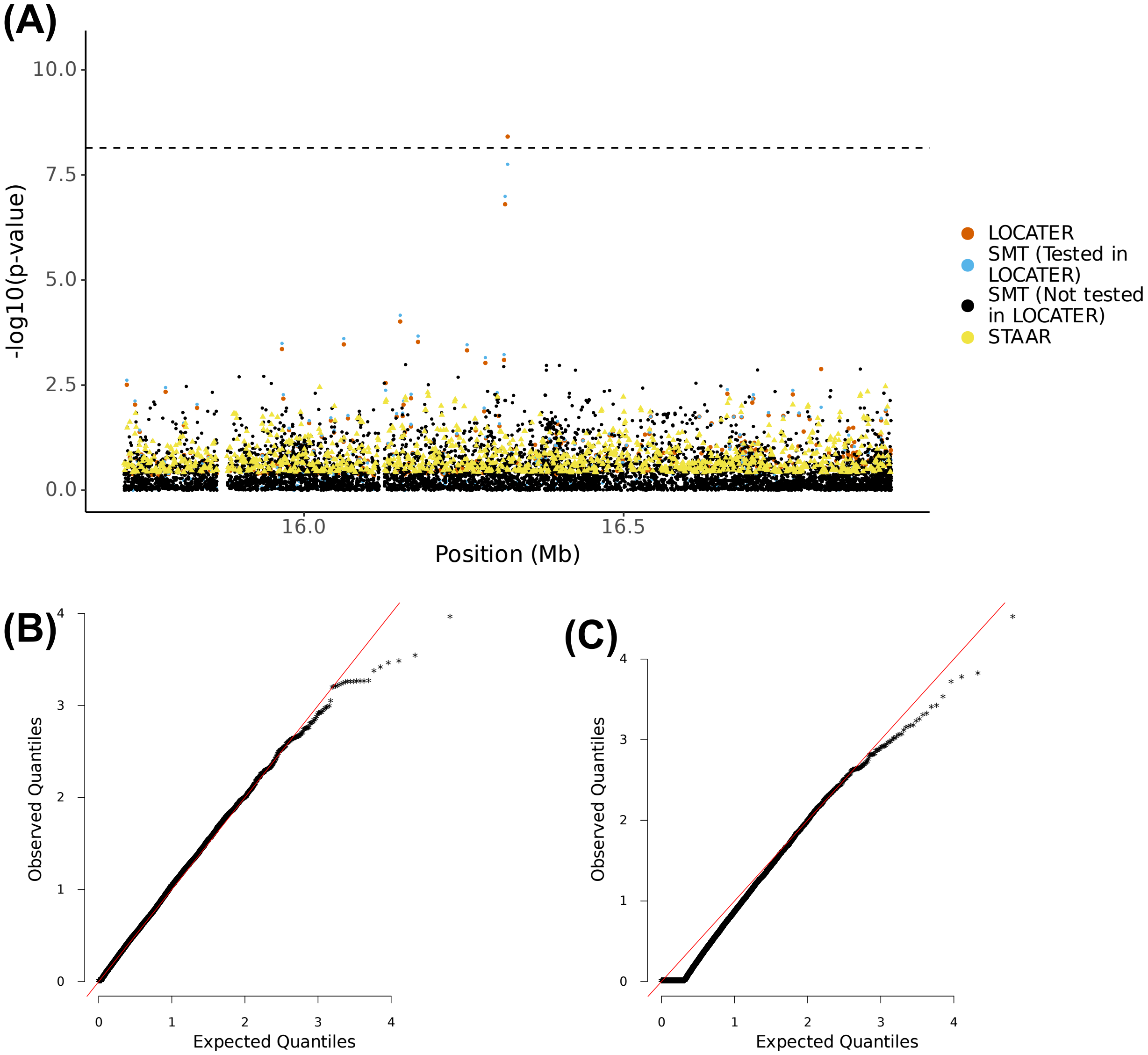


**(A)** Local Manhattan plot of the association signal for HDL3 cholesterol on chr21:15718536-16918536, including results for single marker test (SMT; blue and black), LOCATER (orange) and STAAR (yellow). Note that LOCATER results are only shown for variants with an SMT p-value less than $1x10^{-3}$, since for computational efficiency only these variants were tested by LOCATER (see Methods). SMT results from variants tested by LOCATER are shown in blue, and those from variants not tested by LOCATER are shown in black. The black dashed line corresponds to the genome-wide significance threshold for SMT, standardized LOCATER, and standardized STAAR. **(B)** and **(C)** are Q-Q inflation plots after modified GC of the LOCATER sub-tests stable distillation (SD) and quadratic form (QForm), respectively, for HDL3 cholesterol. The red line on the diagonal corresponds to x=y.

Figure S14D-E

**(D)**

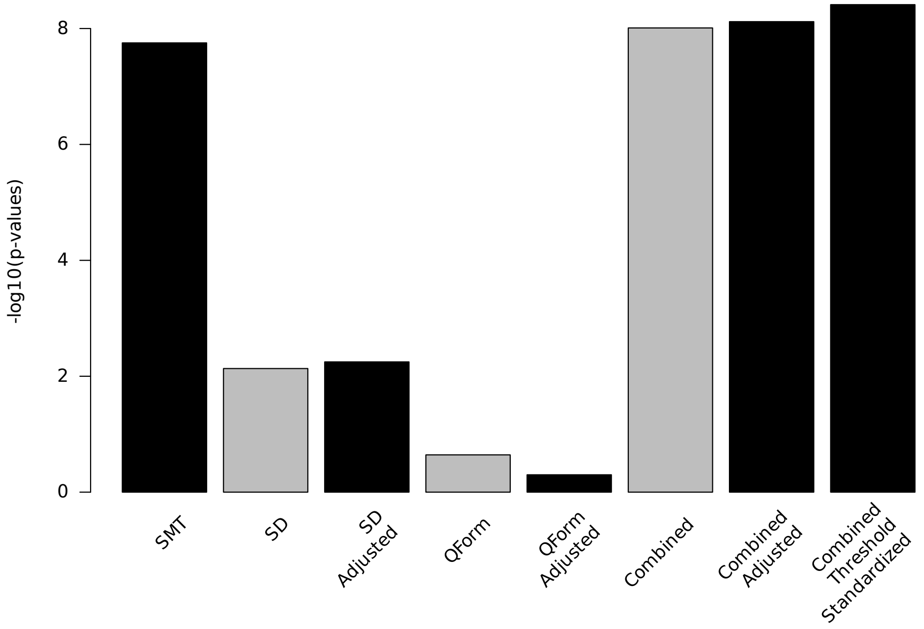


**(E)**

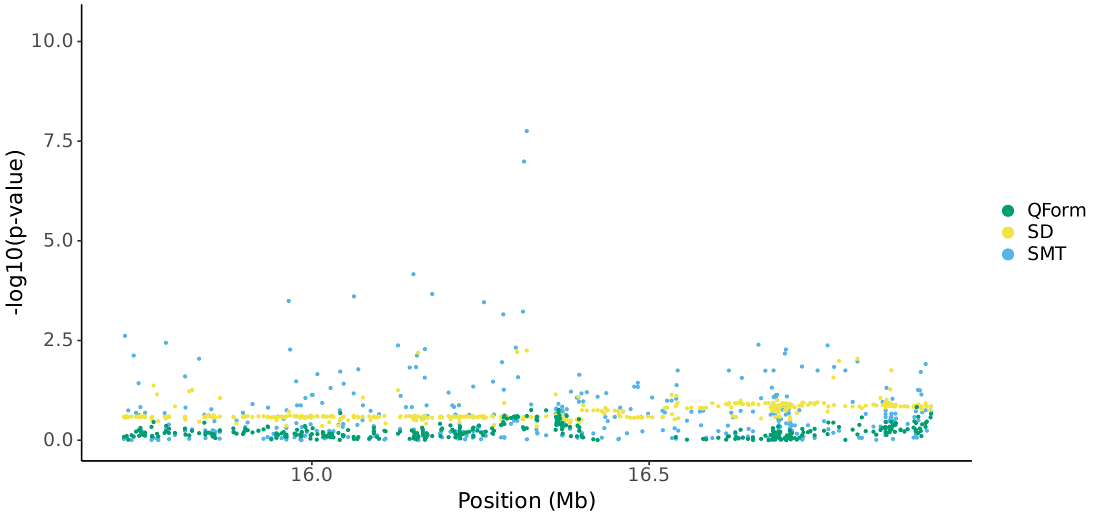


**(D)** Bar plot of -$\log_{10}$(P) for association results before and after modified GC at lead marker chr21:16318536. Shown are the three individual sub-tests, including single marker test (SMT), stable distillation (SD) and quadratic form (Qform), as well as the three tests combined. Grey bars show -$\log_{10}$(P) for SD, QForm and "combined" before modified GC by the slope and intercept of Q-Q plots, black bars show results after modified GC, and the final black bar at right shows the final combined -$\log_{10}$(P) used for all final results, which also accounts for the different number of independent tests performed by SMT and LOCATER. **(E)** Local Manhattan plot of HDL3 cholesterol on chr21:15718536-16918536, showing modified GCed -$\log_{10}$(P) for the 3 LOCATER sub-tests.

### Figure S14F-G

**(F)**

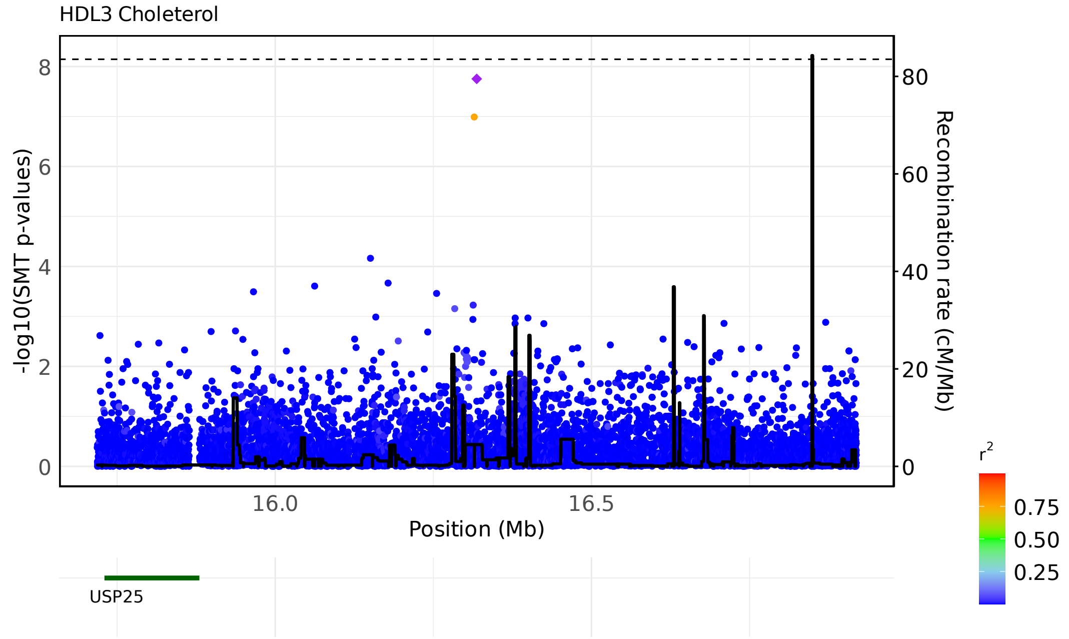


**(G)**

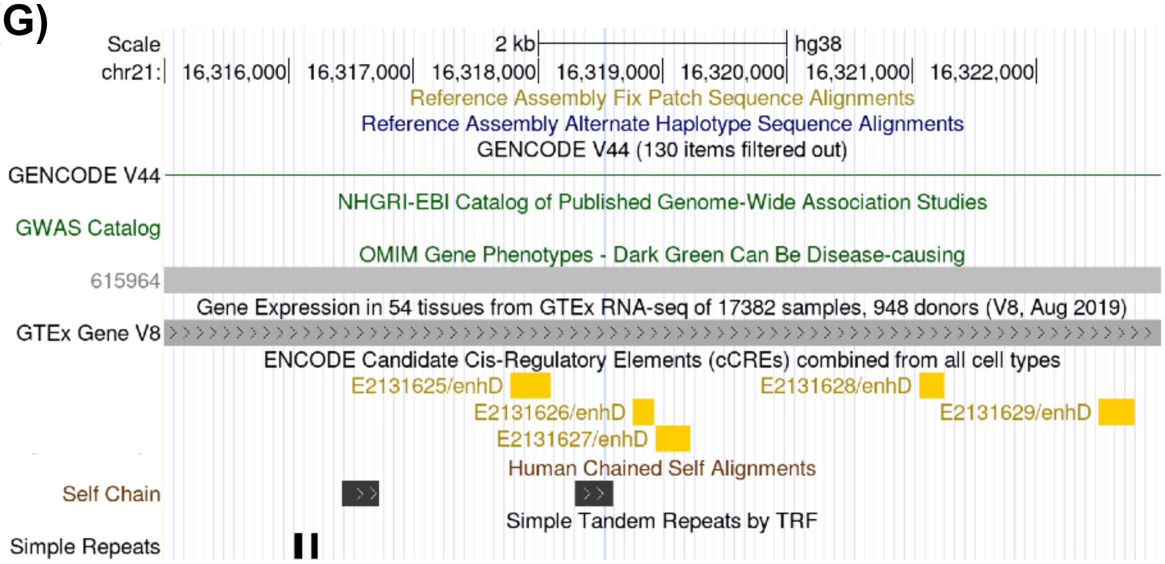


**(F)** LocusZoom plots of SMT results for HDL3 cholesterol on chr21:15718536-16918536. Variants are colored based on their r² with the focal marker (purple diamond), where LD is calculated in the studied samples. The black line shows the recombination rate in Finns (See Methods). Gene annotations are from GENCODE v45. This plot is based on lead marker chr21:16318536. **(G)** Screenshot of the UCSC genome browser, highlighting lead marker chr21:16318536.

#### Figure S14H-I

**(H)**

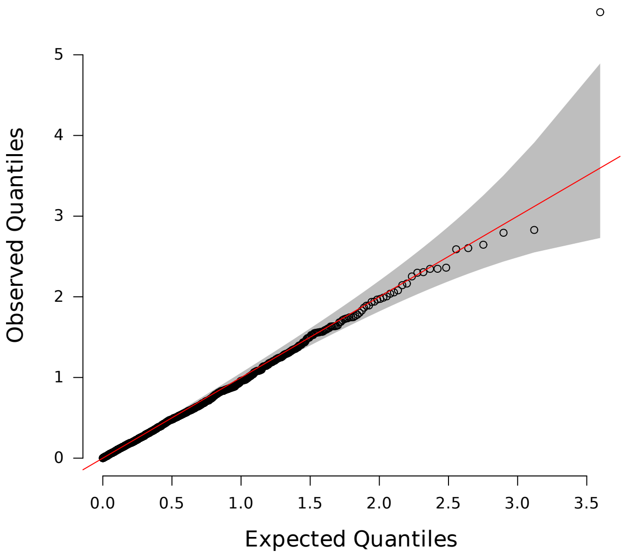


**(H)** Q-Q inflation plot of -log10(p-values) from all "sprigs" at the lead marker chr21:16318536, where "sprigs" are defined as the smallest possible inferred clades. The gray area corresponds to the 95% confidence interval, and the red line denotes x=y. **(I)** Histogram of phenotype values after projecting out the genotype vector of the LOCATER lead marker (chr21:16318536), thus removing signal that can be accounted for by the SMT sub-test. Connected dots show the phenotype value of individuals assigned to significant sprigs.

**(I)**

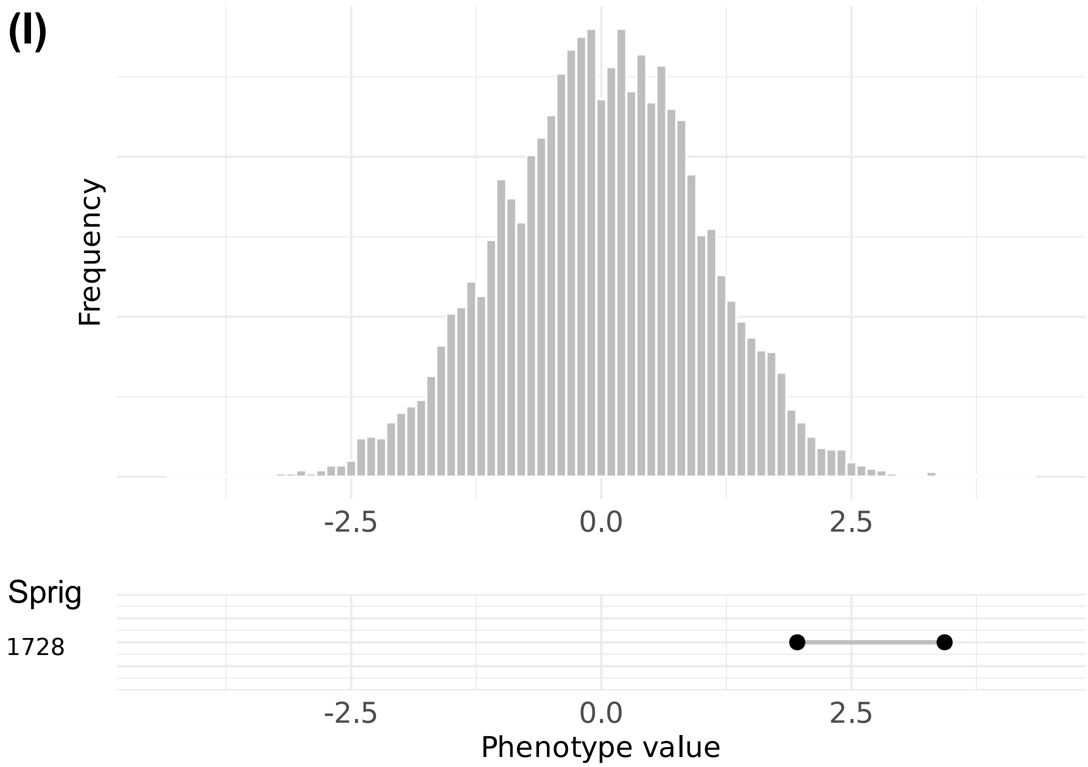

Figure S14J

**(J)**

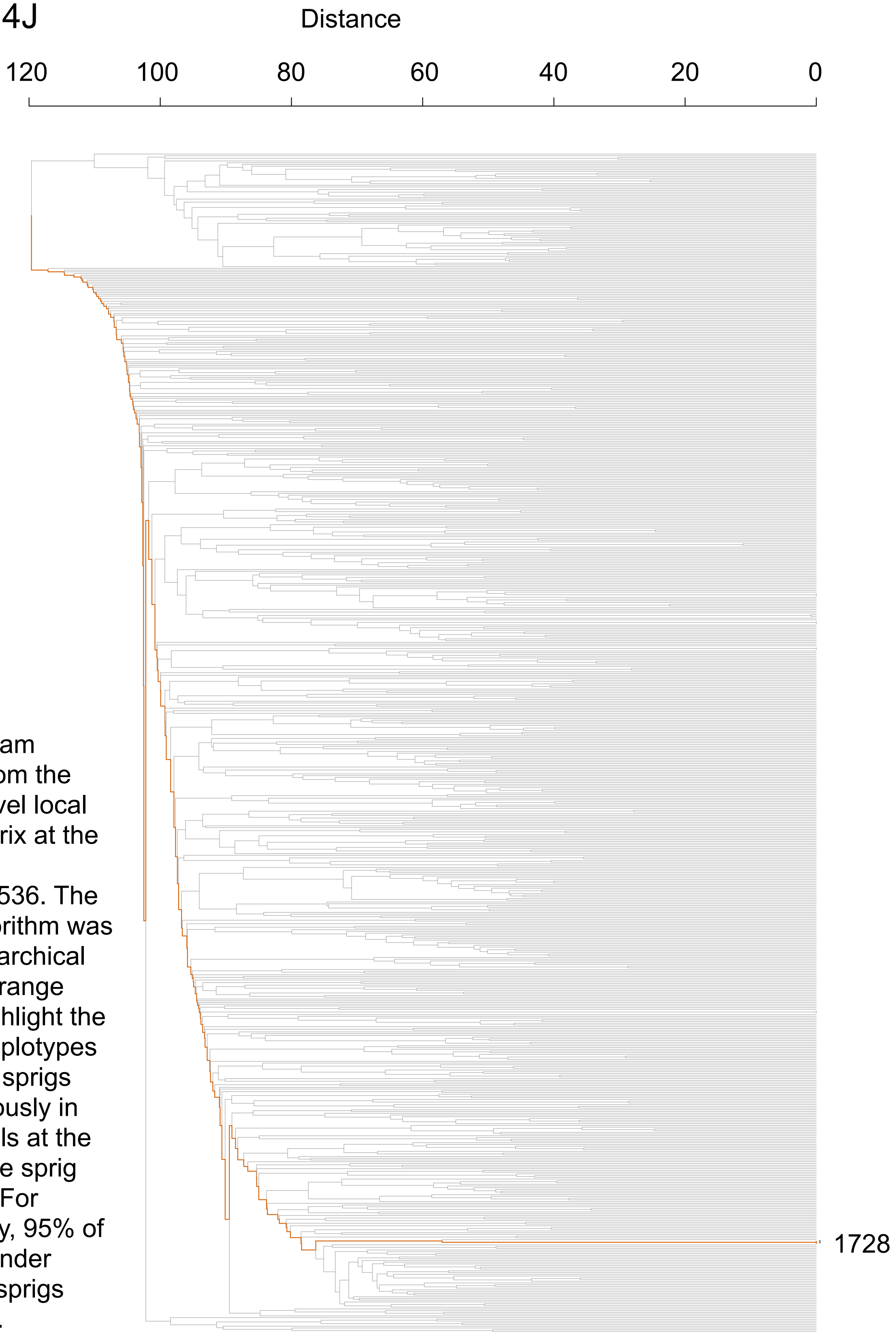


**(J)** Dendrogram generated from the haplotype-level local distance matrix at the lead marker chr21:16318536. The UPGMA algorithm was used for hierarchical clustering. Orange branches highlight the path of all haplotypes in significant sprigs shown previously in part **(I)**. Labels at the right show the sprig assignment. For plotting clarity, 95% of haplotypes under insignificant sprigs were pruned.

#### Figure S14K-L

**(K)**

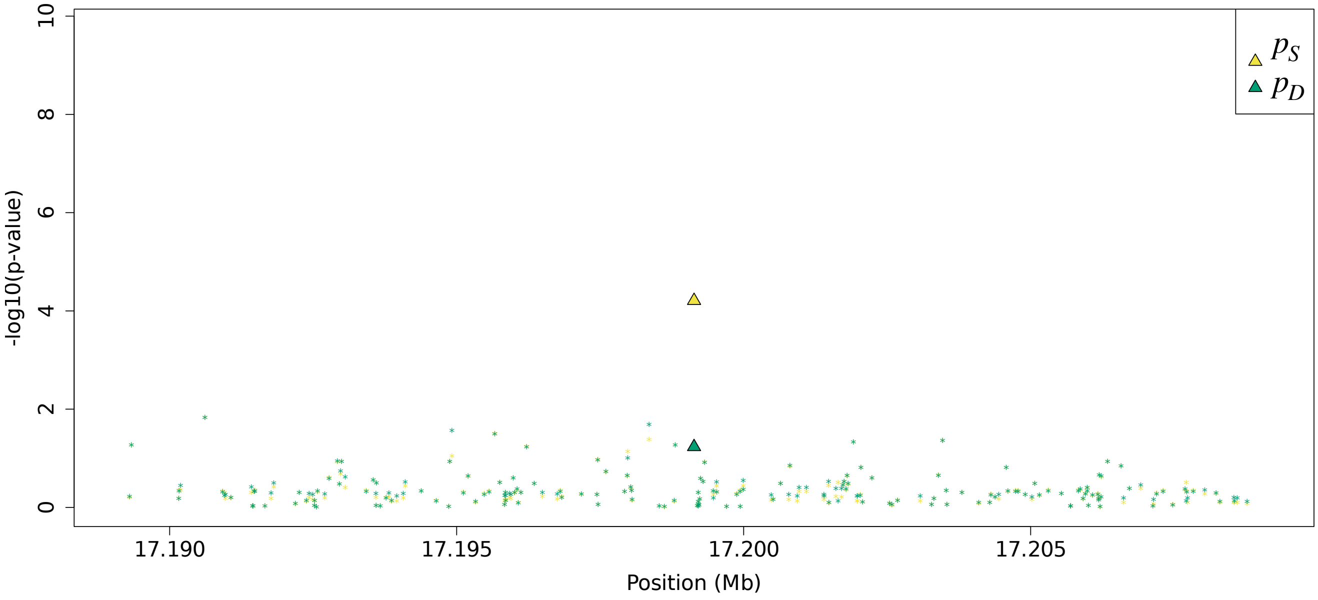


**(L)**

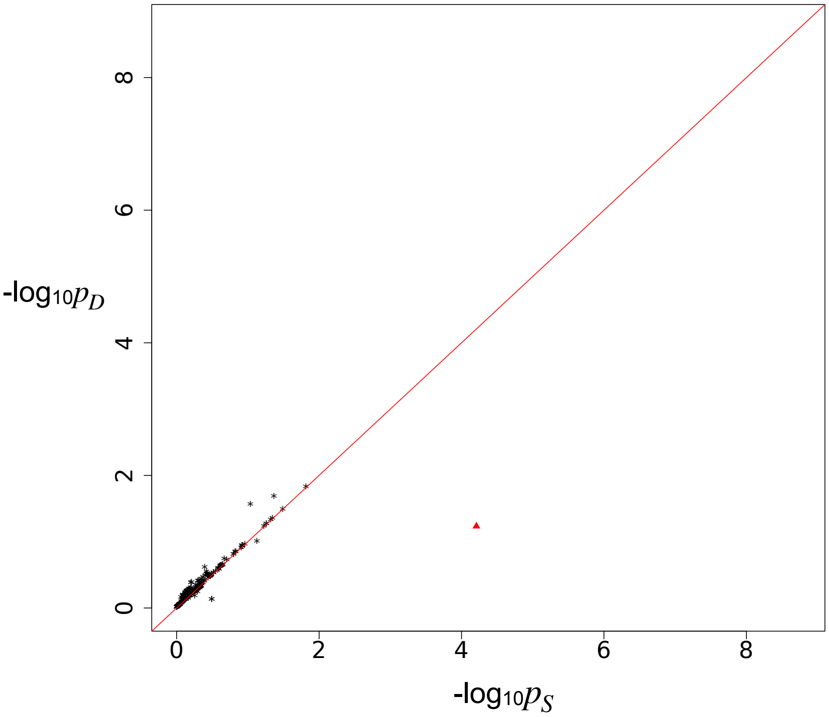


Residual association signals at the *MIR99AHG* locus after accounting for signal from the LOCATER lead marker. $p_S$ is defined as the SMT p-value based on a residualized phenotype orthogonal against the LOCATER lead marker; $p_D$ is the SMT p-value based on a residualized phenotype orthogonal against the LOCATER lead marker and also the SD signal. The difference between $p_S$ and $p_D$ thus shows the contribution of genomic variants to the SD signal. Triangles: genomic variants with $p_S < 10^{-3}$ and $p_D > 10 * p_S$. **(K)** Local Manhattan plot of $p_S$ and $p_D$. **(L)** Scatter plot of $p_S$ and $p_D$.

Figure S14M

**(M)**

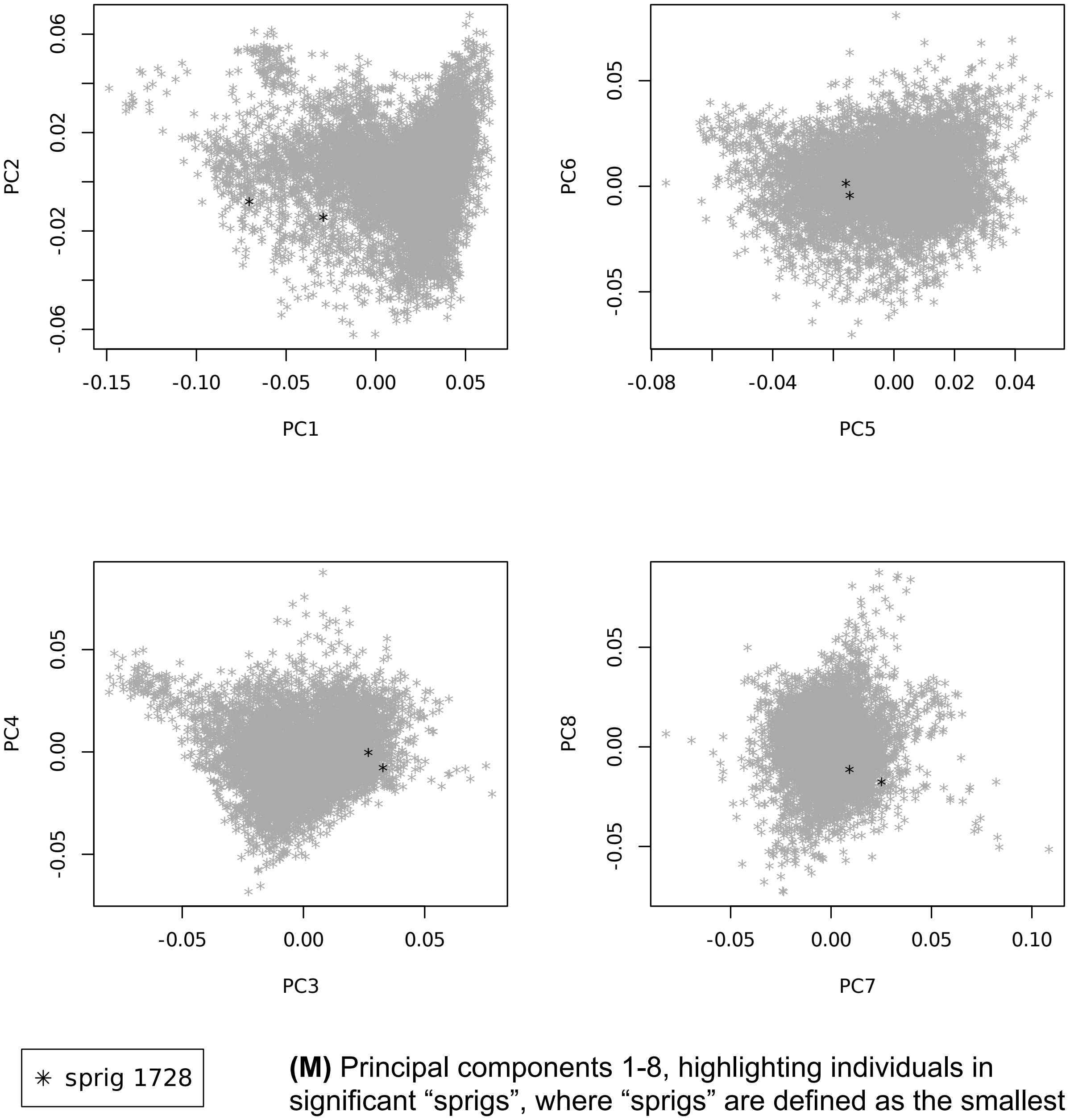


**(M)** Principal components 1-8, highlighting individuals in significant “sprigs”, where “sprigs” are defined as the smallest possible inferred clades. Individuals in the same sprig use the same color and marker.
